# Cortical and Subcortical Neuroanatomical Signatures of Schizotypy in 3,004 Individuals Assessed in a Worldwide ENIGMA Study

**DOI:** 10.1101/2021.04.29.21255609

**Authors:** Matthias Kirschner, Benazir Hodzic-Santor, Mathilde Antoniades, Igor Nenadic, Tilo Kircher, Axel Krug, Tina Meller, Dominik Grotegerd, Alex Fornito, Aurina Arnatkeviciute, Mark A Bellgrove, Jeggan Tiego, Udo Dannlowski, Katharina Koch, Carina Hülsmann, Harald Kugel, Verena Enneking, Melissa Klug, Elisabeth J. Leehr, Joscha Böhnlein, Marius Gruber, David Mehler, Pamela DeRosse, Ashley Moyett, Bernhard T. Baune, Melissa Green, Yann Quidé, Christos Pantelis, Raymond Chan, Yi Wang, Ulrich Ettinger, Martin Debbané, Melodie Derome, Christian Gaser, Bianca Besteher, Kelly Diederen, Tom J Spencer, Paul Fletcher, Wulf Rössler, Lukasz Smigielski, Veena Kumari, Preethi Premkumar, Haeme R. P. Park, Kristina Wiebels, Imke Lemmers-Jansen, James Gilleen, Paul Allen, Petya Kozhuharova, Jan-Bernard Marsman, Irina Lebedeva, Alexander Tomyshev, Anna Mukhorina, Stefan Kaiser, Anne-Kathrin Fett, Iris Sommer, Sanne Schuite-Koops, Casey Paquola, Sara Larivière, Boris Bernhardt, Alain Dagher, Phillip Grant, Theo G. M. van Erp, Jessica A. Turner, Paul M. Thompson, André Aleman, Gemma Modinos

## Abstract

Neuroanatomical abnormalities have been reported along a continuum from at-risk stages, including high schizotypy, to early and chronic psychosis. However, a comprehensive neuroanatomical mapping of schizotypy remains to be established. The authors conducted the first large-scale meta-analyses of cortical and subcortical morphometric patterns of schizotypy in healthy individuals, and compared these patterns with neuroanatomical abnormalities observed in major psychiatric disorders. The sample comprised 3,004 unmedicated healthy individuals (12-68 years, 46.5% male) from 29 cohorts of the worldwide ENIGMA Schizotypy working group. Cortical and subcortical effect size maps with schizotypy scores were generated using standardized methods. Pattern similarities were assessed between the schizotypy-related cortical and subcortical maps and effect size maps from comparisons of schizophrenia (SZ), bipolar disorder (BD) and major depression (MDD) patients with controls. Thicker right medial orbitofrontal/ventromedial prefrontal cortex (mOFC/vmPFC) was associated with higher schizotypy scores (r=.07, p_FDR_=.02). The cortical thickness profile in schizotypy was positively correlated with cortical abnormalities in SZ (r=.33, p_spin_=.01), but not BD (r=.19, p_spin_=.16) or MDD (r=-.22, p_spin_=.10). The schizotypy-related subcortical volume pattern was negatively correlated with subcortical abnormalities in SZ (rho=-.65, p_spin_=.01), BD (rho=-.63, p_spin_=.01), and MDD (rho=-.69, p_spin_=.004). Comprehensive mapping of schizotypy-related brain morphometry in the general population revealed a significant relationship between higher schizotypy and thicker mOFC/vmPFC, in the absence of confounding effects due to antipsychotic medication or disease chronicity. The cortical pattern similarity between schizotypy and schizophrenia yields new insights into a dimensional neurobiological continuity across the extended psychosis phenotype.

## Introduction

Schizophrenia and related psychotic disorders are increasingly recognized as clinical manifestations of quantitative characteristics that are continuously distributed in the general population. In this regard, schizotypy has been linked to trait-like vulnerabilities along a continuum from health to (psychotic) illness [1–3]. Since the early descriptions from Rado and Meehl [4–6], different continuous models of schizotypy and its relationship to schizophrenia have been proposed [7]. A fully-dimensional model assumes that schizotypal traits are not inherently associated with illness but sit squarely within the realm of health, and that clinical manifestations (such as schizophrenia and related psychoses) result from multifactorial breakdown-processes within some (but not all) highly schizotypal individuals in the general population [7–9].

Schizotypy is multi-faceted in nature [10–12], with clinical conditions thought to reflect qualitatively different schizotypy dimensions which interact with general psychopathology [13]. Specifically, schizotypy dimensions closely resemble those of schizophrenia and reflect distinct patterns of positive, negative, and disorganized symptoms [12, 14]. Psychometric measures of schizotypy have been used to assess individuals at clinical-high risk (CHR) for psychosis, and were predictive of conversion to psychosis in both CHR samples and in the general population [1, 15]. Moreover, individuals with high schizotypy exhibit genetic, neurobiological, cognitive, and behavioral characteristics similar (albeit attenuated) to patients with schizophrenia spectrum disorders [16–23]. Thus, in a neurodevelopmental and multifactorial view of a psychosis [24, 25], high schizotypy represents a liability that in concert with other environmental and biological risk factors increases the risk for developing psychosis [9, 26]. Of note, this is not contradictory to a fully dimensional model of schizotypy conceptualizing schizotypal traits as both healthy variations as well as predisposing to psychosis [16]. From a psychosis continuum perspective, healthy individuals with high schizotypy could therefore be placed on the left side, followed by CHR individuals, with schizophrenia spectrum disorders towards the right-most end of the continuum [1].

Converging evidence from neuroimaging research also supports a dimensional view of psychosis. Cortical neuroanatomical abnormalities have been reported in individuals with treatment resistant schizophrenia [27], chronic schizophrenia [28], first-episode psychosis [29], CHR [30, 31], schizotypal personality disorders [32], and individuals with non-clinical psychotic symptoms [33]. Recently, the Schizophrenia Working Group within the ENIGMA (Enhancing Neuro Imaging Genetics through Meta Analysis) consortium provided meta-analytic evidence for robust abnormalities in subcortical volumes [34] and cortical thickness (CT) in schizophrenia [28], while also indicating that these abnormalities are influenced by illness severity and antipsychotic medication [28, 34].

In this context, the study of schizotypy offers a unique opportunity to identify neuroanatomical signatures related to psychosis vulnerability without the common confounds of antipsychotic treatment or disease chronicity [32]. Over the last decade, there has been increased focus on the neuroanatomy of schizotypy [19, 35–43]. The majority of these single studies examined morphometric brain correlates related to total schizotypy scores (including all schizotypy dimensions) [33, 36, 39, 43–45]. Others applied combined approaches using both total scores and schizotypy dimensions [35, 46], or multivariate statistics [42], and reported evidence for shared cross-dimensional neuroanatomical abnormalities of total schizotypy as well as patterns related to distinct dimensions [23, 35, 41, 42]. However, while these studies consistently reported morphometric abnormalities associated with high levels of schizotypy, most studies included relatively small samples and the directionality of the findings was largely inconclusive (e.g., larger or smaller thickness/volumes). Furthermore, almost all studies focused on CT or cortical grey matter volume, rendering the schizotypy literature on subcortical volume and surface area (SA) relatively scarce.

To address these issues, the Schizotypy Working Group within the ENIGMA consortium brought together schizotypy researchers worldwide towards the first large-scale meta-analysis of regional CT, SA and subcortical volumes in schizotypy. We report the first comprehensive neuroanatomical mapping of overall schizotypy using standardized methods in 29 datasets worldwide. To provide meta-analytical evidence that reflects the large body of single neuroanatomical studies in schizotypy presented above, we focused on total schizotypy scores. In line with a dimensional view of psychosis, our main models capitalized on partial correlation effect sizes with continuous measures of schizotypy. Based on recent work in schizophrenia [28, 47, 48], we hypothesized stronger effect sizes for schizotypy-related CT effects, compared to SA and subcortical volumes. Our second aim was to examine the shared morphometric characteristics of schizotypy with previously reported structural abnormalities in schizophrenia, bipolar disorder and major depression. To this end, we correlated subcortical and cortical effect size maps derived from the present meta-analysis with recently published effect size maps from the ENIGMA consortia of these three major psychiatric disorders. We hypothesized that schizotypy-related morphometric patterns would be most similar to morphometric patterns of schizophrenia, relative to bipolar disorder and major depression.

## Methods

### Study Sample

#### Cortical Thickness and Surface Area

Twenty-nine cross-sectional study samples totaling 3,004 unmedicated healthy individuals with varying levels of schizotypy (below) passed Quality Control (QC) and contributed to the cortical meta-analysis. Sample-size average of mean (range) age across samples for this meta-analysis was 30.1 (12–68) years and samples were on average 46.5% male (27%-100%) (Table S1 and Figure S1).

#### Subcortical Volumes

From the same 29 cross-sectional study samples, data from 2,990 healthy, unmedicated individuals passed QC and contributed to the subcortical meta-analysis. Sample-size average of mean (range) age across samples was 30.1 (12.1–67.8) years and samples were on average 46.5% male (26.6%-100%) (Table S3 and Figure S1).

All participants included in this meta-analysis had no current or past history of any psychiatric or neurological disorder. Each study sample was collected with participants’ written informed consent approved by local institutional review boards.

#### Assessment of Schizotypy

Across all 29 centers, schizotypy was assessed with well-validated instruments, including the Chapman scales [49–51], the Community Assessment of Psychotic Experiences (CAPE) [52], the Schizotypal Personality Questionnaire (SPQ) [53], the Oxford-Liverpool Inventory of Feelings and Experiences (O-LIFE) [54], and the Rust Inventory of Schizotypal Cognitions (RISC) [55]. Overall, 18 sites used the SPQ or brief version (SPQ-B), six sites used the CAPE, three sites used the O-LIFE, one site used the Chapman scale, and one site used the RISC. Because our hypotheses involved the association between neuroanatomy with overall schizotypal traits, only total schizotypy scores were included. Associations based on different schizotypy dimensions are to be reported elsewhere [56].

#### Image Acquisition and Processing

Following published ENIGMA pipelines [28, 34], all sites processed T1-weighted structural scans using FreeSurfer [57, 58] (http://surfer.nmr.mgh.harvard.edu) and extracted CT and SA for 70 Desikan-Killiany (DK) atlas regions [59] (34 regions per hemisphere; 1 left and right hemisphere mean thickness or total SA) (Tables S6-S8). Simultaneously, subcortical volumes of 16 brain structures including left and right lateral ventricle, thalamus, caudate, putamen, pallidum, accumbens, hippocampus and amygdala, and intracranial volume (ICV) were extracted (Tables S9-S10). Number of scanners, vendor, strength, sequence, acquisition parameters, and FreeSurfer versions are provided in Table S5. QC followed standard ENIGMA protocols at each site before analysis. For subcortical data, all regions of interest (ROIs) with a volume deviating from the mean by more than 1.5 times from the interquartile range were identified and only included after additional visual inspection. For cortical data, ENIGMA’s quality assurance protocol was performed (http://enigma.usc.edu/protocols/imaging-protocols) including visual inspection of the cortical segmentation and region-by-region removal of values from incorrect segmentations.

### Statistical Meta-Analyses

#### Cortical Measures

Continuous models were fitted to examine the relationship between schizotypy and CT (or SA) in each sample. Partial correlation analysis (*pcor*.*test*, R version 3.6.0, R Foundation for Statistical Computing, Vienna, Austria) was used to assess the association between the CT (or SA) of left and right DK atlas regions with total schizotypy scores including age, sex, and global mean CT (or total SA) as covariates (continuous model 1) and secondarily excluding global mean CT (or total SA) (continuous model 2, Table S7, S9). For multisite studies (n=3), binary dummy covariates were included to account for differences that may emerge across sites. To account for potential confounding effects of smoking on brain morphometry [60, 61], we were able to use data on smoking status that was available for a subsample of participants (n=1,303). Within this subsample, the continuous model 1 including age, sex and global mean CT was repeated with and without smoking as additional covariate to assess whether smoking status would affect the relationship between schizotypy and cortical thickness (Tables S14 and S15).

#### Subcortical Volumes

Similar to the cortical analyses, continuous models were applied to examine the relationship between schizotypy scores and subcortical volume for each ROI in each sample. To this end, partial correlation analysis was used to test correlations between the left and right subcortical volumes with total schizotypy scores. The main model included age, sex, and ICV as covariates. For multisite studies (n=3), binary dummy covariates were included to account for differences that may emerge across sites. As for the cortical analysis, partial correlation analysis was repeated in a subsample of n=1,303 participants including smoking status as covariate and are reported in Tables S16 and S17.

#### Meta-Analyses

For cortical and subcortical measures, Pearson’s *r* effect sizes from the partial correlations using schizotypy scores as a continuous predictor were meta-analyzed in separate random effects models to account for between study differences (*rma* function, metafor package for R 3.6.0) [62]. The false discovery rate (FDR) procedure (p_FDR_<0.05) was used to control for multiple comparisons [63, 64]. Meta-analyses were adjusted for sample sizes across different sites and results were weighted for sample sizes. Possible confounding effects of schizotypy questionnaire type, differences in average schizotypy severity between sites, FreeSurfer version and scanner field strength were examined using moderator analyses (Table S21-S24).

#### Cortical and Subcortical Pattern Similarity between Schizotypy and Major Psychiatric Disorders

To answer our second question on how overall CT and subcortical volumes in schizotypy relate to the neuroanatomical patterns observed in schizophrenia, bipolar disorder and major depression, we correlated the schizotypy-related effect size maps of correlation coefficient r for CT and SA (age, sex corrected), as well as for subcortical volumes (age, sex, ICV corrected) with the Cohen’s d maps derived from recently published meta-analyses of case-control studies by the ENIGMA schizophrenia (SZ) [28, 34], bipolar disorder (BD) [65, 66], and depression (MDD) working groups [67, 68]. This approach followed previous studies correlating cortical effects size maps between different disorders [69, 70] or disorders and regional network features of the brain [71]. Specifically, we applied a recently developed approach from the ENIGMA Epilepsy Working Group using Pearson correlation to investigate spatial pattern similarity of cortical effect size maps [71]. Statistical significance of all cortical pattern correlations was assessed using spin permutation tests correcting for spatial autocorrelation [72, 73], following recent work from the ENIGMA Epilepsy Group [71]. In this framework, null models are generated by projecting the spatial coordinates of cortical data onto the surface spheres, applying randomly sampled rotations (10,000 repetitions), and reassigning cortical data (here effect size values) [72]. The original correlation coefficients are then compared against the empirical distribution of spatially permuted correlation coefficients.

Spatial pattern similarity between subcortical maps was examined using Spearman rank correlations to account for potential outlier effects in small sample sizes [74]. Statistical significance testing followed a similar approach as the spin permutation from the cortical analysis with the exception that subcortical labels were randomly shuffled as opposed to being projected onto spheres [71]. We hypothesized that the schizotypy-related cortical or subcortical effect size maps would be positively correlated with the corresponding SZ effect size maps, suggesting neuroanatomical continuity of the schizophrenia spectrum. We additionally hypothesized that the strength of cortical and subcortical relationships would follow a psychosis-to-affective disorder axis (SZ>BD>MDD). To this end, pairwise comparisons of the correlations between schizotypy-SZ with schizotypy-BD and schizotypy-MDD were performed using one-tailed Steiger’s test (*cocor* package, R 3.6.0) [75].

## Results

### Cortical Thickness and Schizotypy

Meta-analysis of the continuous relationship between CT and schizotypy (n=3,004) revealed positive correlations between higher schizotypy and greater thickness of the medial orbitofrontal cortex / ventromedial prefrontal cortex (mOFC/vmPFC) (left: r=.057, p_unc_=.004, 95% CI [.02 - .09]; right: r=.067, p_unc_<.001, 95% CI [.03, .10]) as well the frontal pole (left: r=.046, p_unc_=.02, 95% CI [.01, .08]; right: r=.050, p_unc_=.007, 95% CI [.01, .09]) (Figure 1A, Table S6). The positive association between greater right mOFC/vmPFC thickness and higher schizotypy remained significant after FDR correction (r=.067, p_FDR_=.02) (Figure 2A/C). Moderator analyses did not reveal any significant effects of type of schizotypy questionnaire, FreeSurfer version or scanner field strength on the associations between cortical thickness and schizotypy (all p_FDR_>.05) (Table S21-23).

**Figure 1.**
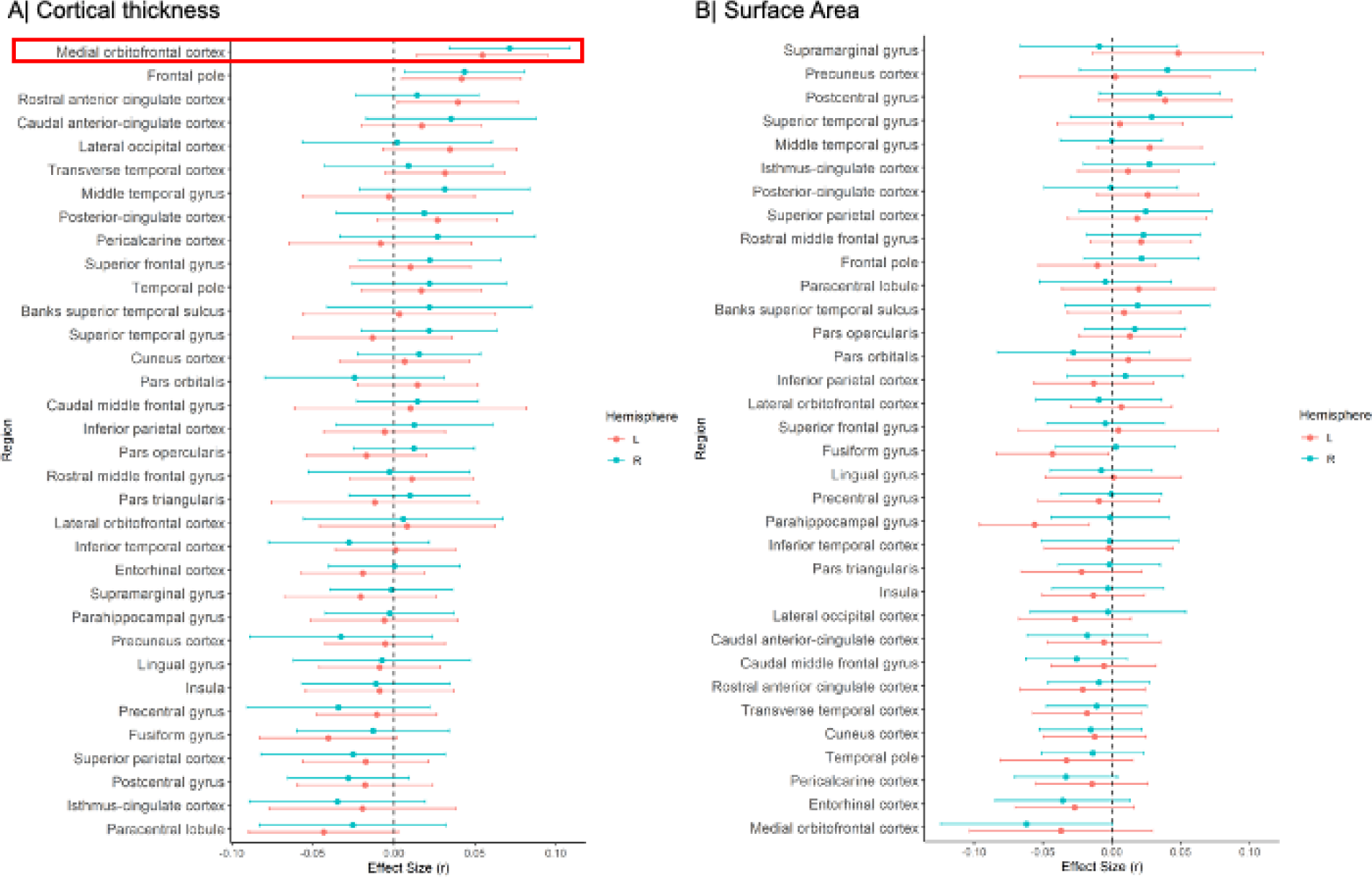
Effect sizes of partial correlation (r) between cortical thickness, surface area and schizotypy. (A) Cortical thickness, (B) Surface area. Effect sizes for all regions depicted were corrected for age, sex and global cortical thickness or total surface area, respectively. Red rectangle highlights effects surviving false discovery rate (FDR) correction (p_FDR_<0.05).

**Figure 2.**
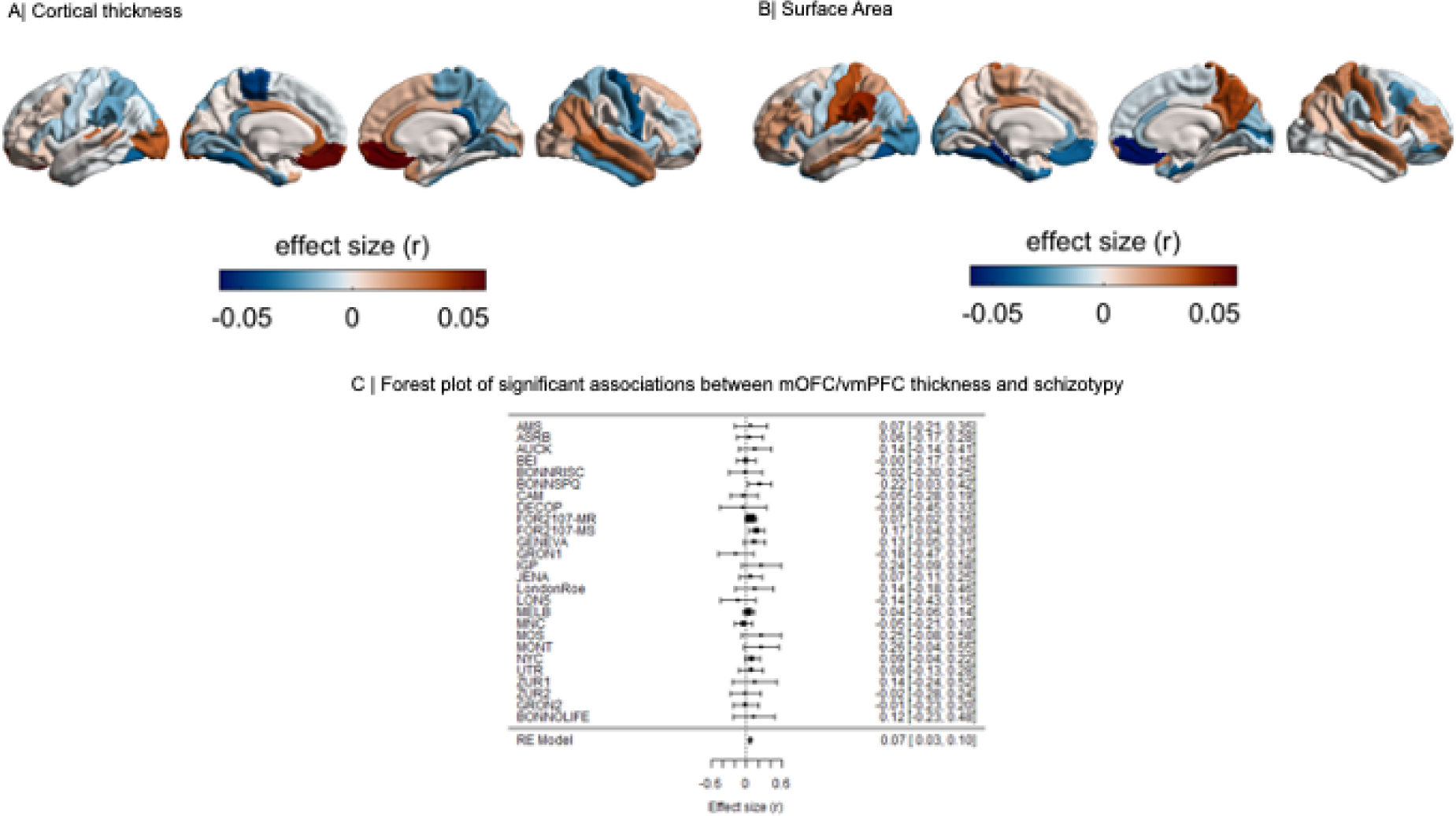
Cortical maps of regional effect sizes for associations with schizotypy. (A) Cortical thickness, (B) Surface area. (C) Forest plot of the significant association between mOFC/vmPFC thickness and schizotypy, after false discovery rate (FDR) correction (p_FDR_<0.05).

### Surface Area and Schizotypy

Higher schizotypy was associated with lower SA in the left parahippocampal gyrus (r=-.056, p_unc_=.006, 95% CI [-.096, -.016]) and left fusiform cortex (r=-.043, p_unc_=.04, 95% CI [-.084, - .002]) (Table S8), but these effects did not survive correction for multiple comparisons (p_FDR_<.05) (Figure 1B, Figure 2B).

### Subcortical Volume and Schizotypy

When examining the continuous relationship between subcortical volumes of 16 brain structures and schizotypy scores (n=2,990), only non-significant correlations were found, ranging from negative correlations between higher schizotypy and lower volume in the right pallidum (r=-.032, p_unc_=.08, 95% CI [-.07, .004]), to positive correlations between higher schizotypy and greater volume in the right accumbens (r=.020, p_unc_=.38, 95% CI [-.02, .06]), right amygdala (r=.019, p_unc_=.44, 95% CI [-.03, .07]), and right hippocampus (r=.022, p_unc_=.23, 95% CI [-.01, .06]) (Figure 3). All subcortical results from the continuous analysis are summarized in Table S13.

**Figure 3.**
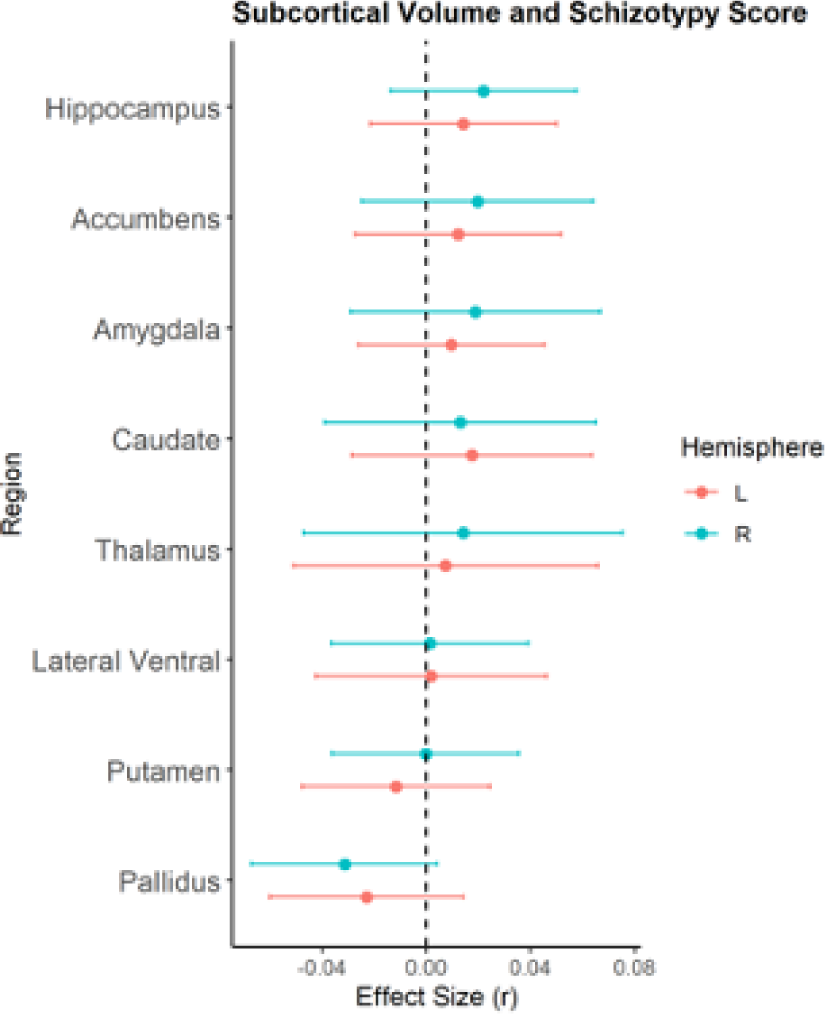
Effect sizes of partial correlation (r) between subcortical volumes and schizotypy. Effect sizes for all subcortical volumes depicted were corrected for age, sex and intracranial volume (ICV).

*Cortical and Subcortical Pattern Similarity Between Schizotypy and Major Psychiatric Disorders* Correlations of cortical maps revealed significant positive associations between increasing levels of schizotypy-related morphometry (partial correlation r) and Cohen’s d maps from ENIGMA SZ (r=.285, p_spin_=.024), but not ENIGMA BD (r =.166, p_spin_=.205) or ENIGMA MDD (r=-.27, p_spin_=.073) (Figure 4). Pairwise comparisons of these correlations revealed that the relationship between schizotypy and SZ cortical patterns was significantly stronger than the correlations between schizotypy and MDD (Z=4.063, p-value<.0001, 95% CI [0.3 - 0.8]) and at trend level to the correlation between schizotypy and BD (Z=1.462, p=.072, 95% CI [-0.04 - 0.8]). These findings indicate that the schizotypy-related CT pattern is more closely related to cortical brain alterations in SZ compared to BD and MDD.

**Figure 4.**
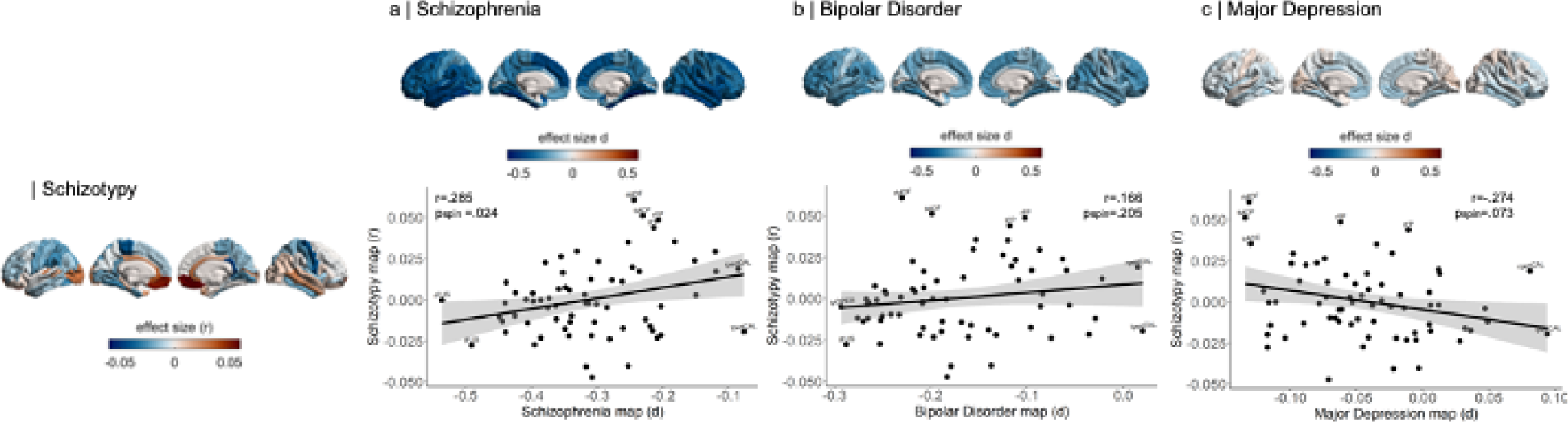
Pattern similarity between cortical thickness effects in schizotypy and major psychiatric disorders. Abbreviations of the cortical regions are adopted from the brainGraph package and shown for regions with the most positive and negative effect sizes (r): FUS fusiform gyrus, ITG inferior temporal gyrus, iCC isthmus cingulate cortex, MOF medial orbitofrontal cortex, paraC paracentral, lobule, pOPER pars opercularis of inferior frontal gyrus, periCAL pericalcarine cortex, rACC rostral anterior cingulate cortex, FP frontal pole, INS insula. L left, R right.

Repeating the cortical pattern similarity analyses with SA effect size maps of schizotypy and all three psychiatric disorder maps revealed that the schizotypy related SA pattern was not correlated with the SA patterns of either SZ (r= -.009 p_spin_=.95), BP (r=-.221 p_spin_=.14) or MDD (r=-.05 p_spin_=.70). These findings suggest that the observed cortical pattern similarity between schizotypy and schizophrenia is specific for cortical thickness rather than surface area.

In terms of subcortical volumes, effect sizes for schizotypy-related patterns (partial correlation *r*) were significantly negatively associated with the profile of Cohen’s *d* values for subcortical volume abnormalities in ENIGMA SZ (rho=-.690, p_spin_=.006), ENIGMA BD (rho=-.672, p_spin_=.009), and ENIGMA MDD (rho=-.692, p_spin_=.004) (Figure 5). In other words, subcortical volumes in schizotypy showed an opposite pattern to subcortical abnormalities in SZ, BD or MDD, with similar magnitudes for all three correlations. The largest differences in effect sizes between subcortical profiles of schizotypy and all three psychiatric disorders were observed in the hippocampus. In particular, patients with SZ but also BD and MDD showed smaller hippocampal volume compared to controls, while in healthy individuals higher schizotypy was (weakly) positively correlated with greater hippocampus volume (Figure 5). In addition, for SZ the opposite effect was observed for pallidum volume, displaying larger volume in patients compared to controls, and a (weak) negative correlation between smaller volume and higher schizotypy in healthy individuals (Figure 5).

**Figure 5.**
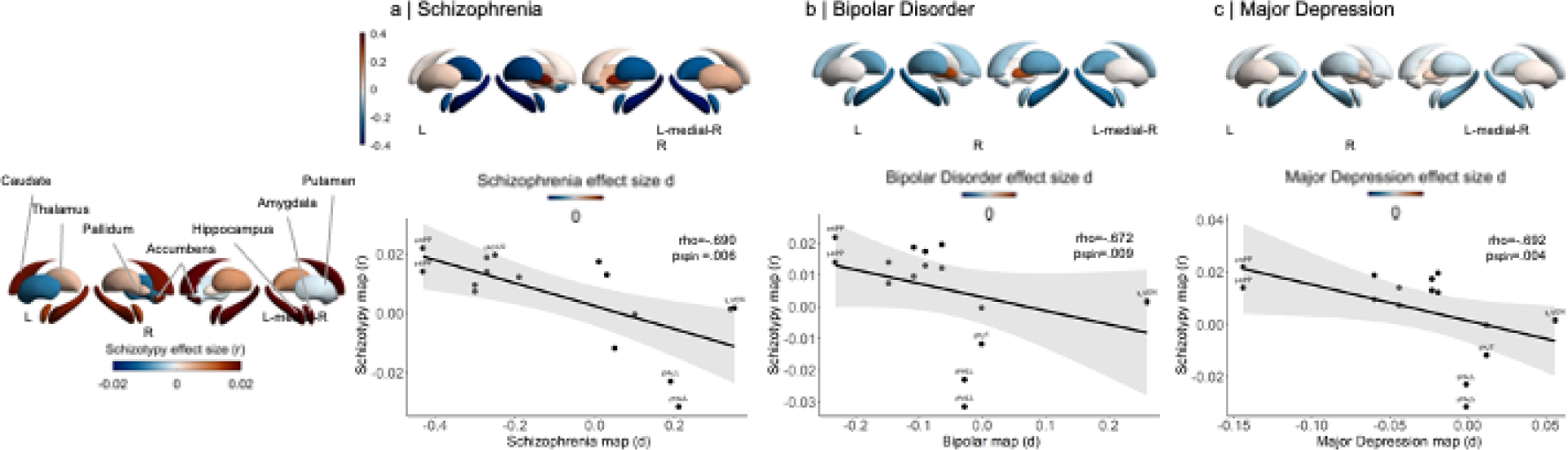
Pattern similarity between subcortical volume effects in schizotypy and major psychiatric disorders. Abbreviations of subcortical regions are adopted from the brainGraph package and are shown for regions with the most positive and negative effect sizes (r): HIPP hippocampus, PALL pallidum, PUT putamen, LVEN lateral ventricle, l left, r right.

## Discussion

Leveraging neuroimaging data from 29 international sites, the present work is the first large-scale meta-analysis of cortical and subcortical measures in schizotypy, including over 3,000 individuals. Our main finding was that greater CT in the mOFC/vmPFC was positively associated with higher schizotypy. Subcortical volume and SA analyses showed only subtle, non-significant correlations with schizotypy scores. These findings were, due to our healthy sample, free of potential influences of illness chronicity or antipsychotic medication on neuroanatomy. Moreover, we found a positive association between the schizotypy-related CT pattern and the pattern of CT abnormalities observed in SZ, but not with that of BD or MDD, supporting the notion of neurobiological continuity across the extended psychosis phenotype. In contrast, patterns of schizotypy-related subcortical effects showed an unexpected negative association with patterns of subcortical volume abnormalities more generally, across the three psychiatric disorders.

The directionality of the observed association for CT, reflecting thicker mOFC/vmPFC with higher schizotypy, is inverse to findings of prefrontal cortical thinning commonly observed in medicated patients with schizophrenia [28, 47] and first-episode psychosis [27, 43]. However, our findings are in line with reports of greater OFC thickness in drug-naïve first-episode schizophrenia patients [76, 77]. Prior schizotypy studies reported mixed findings, including greater prefrontal [38] or spared OFC thickness [36], and lower frontal grey matter volume [37, 42, 46]. Findings from studies in individuals at CHR for psychosis have likewise been inconsistent, with evidence for thicker OFC [78, 79], no differences [80] or thinner cortex [81] compared to healthy controls. This heterogeneity among these single studies may be partly explained by the use of different methods or limited power. On the other hand, there is growing evidence that variations of prefrontal CT are related to different at-risk phenotypes and the onset of early and late neurodevelopmental disturbances [25, 82]. Different anatomical trajectories have been reported in schizotypy [35], between different at-risk states [83] and particularly in CHR individuals who subsequently develop psychosis [82, 84]. Transition to psychosis has been associated with a steeper cortical thinning of heteromodal cortices, including the mOFC/vmPFC, compared to CHR individuals who do not convert to psychosis and healthy controls [85–88]. Thus, structural characteristics of the mOFC/vmPFC may vary based on the degree of risk and illness chronicity, from relatively thicker mOFC/vmPFC in healthy individuals with high schizotypy, to normal-to-elevated in CHR with lesser transition risk, to thinner in CHR individuals who convert to psychosis, to further thinning in chronic schizophrenia.

The observed relationship between high schizotypy and greater mOFC/vmPFC thickness shares neuroanatomical similarities to other neurodevelopmental disorders such as autism spectrum disorders (ASD) (Cohen’s d=0.15, p_FDR_=.0001 [89]) and 22q deletion syndrome (Cohen’s d=0.61, p_FDR_<.0001 [69]). Such convergence aligns with observations of increased phenotypic expression of schizotypy in ASD [90, 91] and 22q deletion syndrome [92]. Together, these findings support the notion that high schizotypy may describe a predisposing trait of genetically and clinically overlapping phenotypes with schizophrenia-spectrum disorders and ASD [16, 93, 94]. Our cross-disorder whole-brain cortical mapping revealed a strong link between morphometric signatures of schizotypy with schizophrenia, along a SZ-BD-MDD axis. These findings partly resemble previous genetic [95] and neuroanatomical correlations [70, 96] observed between schizophrenia, bipolar disorder and major depression [97]. Interestingly, the relationship between the cortical patterns of schizotypy and schizophrenia revealed regional discordant and concordant effect sizes. Specifically, in schizotypy, the associations between higher schizotypy and lower CT were only observed in those regions showing the strongest negative effect sizes in schizophrenia (e.g. thinning), such as the left and right fusiform gyrus or inferiotemporal gyrus. In other words, schizotypy and schizophrenia showed somewhat concordant effect sizes in these regions. In contrast, discordant effect sizes of thinner CT in schizophrenia and greater CT in high schizotypy were observed in other regions such as the rostral anterior cingulate, frontal pole and mOFC/vmPFC. Here, higher schizotypy was positively associated with greater CT, while patients with schizophrenia showed thinner cortex compared to controls (negative effect size). This neuroanatomical pattern of spatially distributed thinner and thicker cortex might reflect that in schizotypy samples, which comprise healthy individuals that will most likely not develop a full-blown psychotic disorder, effects may be subtle. Although speculative, the relative greater cortical thickness could reflect either compensatory mechanisms or a pre-existing condition before the onset of accelerated thinning in those individuals that develop psychosis.

Whereas the cortex-wide similarity between schizotypy and schizophrenia provides support for a specific relationship, schizotypy-related subcortical volumes showed a negative association with subcortical abnormalities in all three disorders (SZ, BD, MDD). Combined with the overall subtle links between schizotypy and subcortical volume, these inverse correlations may suggest that schizotypal traits are not directly linked to subcortical alterations of major psychiatric disorders in general, which may be better explained by other risk/disease factors (e.g., medication, disease course, common mental distress) [34, 66, 68], or that spared subcortical volume changes might be a protective factor in those individuals with high schizotypy and high resilience [16, 98, 99]. In line with the latter, first-degree relatives of bipolar disorder patients showed higher than normal ICV (compared to controls), suggesting putative survivor effects in unaffected individuals with high liability for psychiatric disorders [100]. While an inverse subcortical pattern in schizotypy was not anticipated, further investigation and replication of these findings will be of interest.

Overall, our meta-analyses suggest that schizotypal traits in the general population are predominantly embedded in CT effects, while less associated with variations of subcortical volume or SA. The nature of preserved or greater cortical morphometry in schizotypy has been intensively debated, although the underlying cellular and molecular mechanisms remain unclear. Greater prefrontal CT might reflect abnormal or delayed cortical development [101–103], due to insufficient synaptic pruning [104] or altered cortical myelination [105, 106]. Greater CT as a result of microstructural and cellular perturbations may therefore be associated with higher vulnerability, while schizophrenia may only emerge in the presence of additional environmental, biological or genetic factors [25, 82, 107]. In line with this notion, several previous studies in CHR, genetic and clinical samples have shown that measures of schizotypy can be predictive for developing psychosis [1], although the predictive value of schizotypy might be sensitive to vulnerable periods (e.g. adolescence) and potentially increase with accumulation and interaction of other psychosis risk factors [26]. Alternatively, it has been proposed that neuroanatomical signatures such as preserved/greater prefrontal CT could reflect protective mechanisms for developing clinical symptoms of psychosis/schizophrenia in high schizotypy individuals with absence of other risk factors [16, 99]. This would align with a fully-dimensional model of schizotypy [7, 8], which predicts inverse associations in schizotypy samples consisting of healthy participants compared to samples of patients with schizophrenia, supported by findings that frontal capacity is positively associated with schizotypy [99]. Large-scale longitudinal studies from neurodevelopment in childhood to early adulthood could help to further differentiate neuroanatomical trajectories of high schizotypy and their contribution to either psychosis in those individuals with concomitant risk factor [26] or to normative variations in otherwise healthy individuals [16].

## Limitations

In interpreting the current findings, it is relevant to note that rather small effect sizes are typical for these types of large-scale neuroimaging meta-analysis and comparable effect size magnitudes have been reported in other ENIGMA studies of clinical populations [67, 89] and populations at higher risk for mental illnesses [100, 108]. Given that schizotypal traits were derived from healthy individuals in the general population, relationships between brain morphometry and schizotypy were expected to be even more subtle compared to clinical populations. The observed effect sizes can be positioned at the lower end of cortical effects in psychiatric disorders (<SZ and BD [28, 65] but are comparable to those reported for MDD and ASD [67, 89]) as well as population neuroscience studies of associations between brain structure and polygenic risk for psychosis or psychotic symptoms [109, 110]. Furthermore, no moderating effects of the type of schizotypy questionnaire, FreeSurfer version, or scanner field strength were found supporting the strength of the observed findings. This first meta-analysis aimed at reflecting the large body of research on neuroanatomical patterns of total schizotypy within the psychosis continuum. However, the use of total schizotypy scores might have obscured some neuroanatomical differences related to distinct schizotypy dimensions (e.g. positive, negative and disorganized) [35, 111]. The ENIGMA Schizotypy Working Group is actively working on pooling data to map neuroanatomical patterns of separate schizotypy dimensions, which will be reported elsewhere [56].

To advance our understanding of the neurobiology of psychosis risk beyond MRI measurements associated with overall schizotypy levels, there will be continued effort by the ENIGMA Schizotypy Working Group to (i) include more datasets, (ii) incorporate genetic as well as multimodal neuroimaging data, and (iii) identify associations between MRI-derived measures and cell-type specific gene expression [97].

## Conclusion

In summary, this is the first meta-analysis of neuroimaging data to comprehensively map the morphometric signature of schizotypy in healthy individuals. The results suggest a profile of CT abnormalities involving thicker prefrontal cortex related to more severe schizotypy. The CT pattern related to schizotypy was most closely linked to CT abnormalities in schizophrenia, thus providing neuroanatomical support for dimensional continuity across the extended psychosis phenotype.

## Data Availability

Data are available from the authors upon request with permission from the chairs and members of the ENIGMA Schizotypy Working Group.

## Funding and Acknowledgments

Core funding for ENIGMA was provided by the NIH Big Data to Knowledge (BD2K) program under consortium grant U54 EB020403 (PI: Paul M Thompson). GM is supported by a Sir Henry Dale Fellowship jointly funded by the Wellcome Trust and the Royal Society (grant number 202397/Z/16/Z). Acknowledgments and funding details for the various participating data contributors are listed in at the end of the Supplement.

## Disclosures

PMT received partial grant support from Biogen, Inc. (Boston, USA) for work unrelated to this manuscript. SK received speaker honoraria from Janssen, Takeda, Lundbeck and Roche. Royalties for cognitive test and training software from Schuhfried. In the past 3 years, CP served on an advisory board for Lundbeck, Australia Pty Ltd. He has received honoraria for talks presented at educational meetings organized by Lundbeck. No other disclosures were reported.

## Role of the Funder/Sponsor

The funders had no role in the design and conduct of the study; collection, management, analysis, and interpretation of the data; preparation, review, or approval of the manuscript; and decision to submit the manuscript for publication.

## Author Contributions

### Author contributions

Kirschner, Hodzic-Santor and Modinos had full access to all of the data in the study and take responsibility for the integrity of the data and the accuracy of the data analysis. Kirschner and Hodzic-Santor contributed equally to this work.

### Study concept and design

Kirschner, van Erp, Turner, Thompson, Aleman and Modinos.

### Acquisition, analysis, interpretation of data, and funding (in alphabetical order)

Aleman, Allen, Antoniades, Arnatkeviciute, Baune, Bellgrove, Bernhardt, Besteher, Böhnlein, Chan, Dagher, Dannlowski, Debbané, Derome, DeRosse, Diederen, Enneking, Ettinger, Fett, Fletcher, Fornito, Gaser, Gilleen, Grant, Green, Grotegerd, Gruber, Hülsmann, Hodzic-Santor, Kaiser, Kircher, Kirschner, Klug, Koch, Koops, Kozhuharova, Krug, Kugel, Kumari, Larivière, Lebedeva, Leehr, Lemmers-Jansen, Marsman, Mehler, Meller, Modinos, Moyett, Mukhorina, Nenadic, Pantelis, Paquola, Park, Premkumar, Quidé, Rössler, Sommer, Smigielski, Spencer, Tiego, Thompson, Tomyshev Turner, van Erp, Wang, Wiebels.

### Statistical analysis

Kirschner, Hodzic-Santor, Antoniades, Marsman.

### Drafting of the manuscript

Kirschner, Hodzic-Santor and Modinos.

### Critical revision of the manuscript for important intellectual content (in alphabetical order)

Aleman, Allen, Antoniades, Arnatkeviciute, Baune, Bellgrove, Bernhardt, Besteher, Böhnlein, Chan, Dagher, Dannlowski, Debbané, Derome, DeRosse, Diederen, Enneking, Ettinger, Fett, Fletcher, Fornito, Gaser, Gilleen, Grant, Green, Grotegerd, Gruber, Hülsmann, Hodzic-Santor, Kaiser, Kircher, Kirschner, Klug, Koch, Koops, Kozhuharova, Krug, Kugel, Kumari, Larivière, Lebedeva, Leehr, Lemmers-Jansen, Marsman, Mehler, Meller, Modinos, Moyett, Mukhorina, Nenadic, Pantelis, Paquola, Park, Premkumar, Quidé, Rössler, Sommer, Smigielski, Spencer, Tiego, Thompson, Tomyshev Turner, van Erp, Wang, Wiebels. All authors approved the contents of the manuscript.

## Notes

### Competing Interest Statement

The authors have declared no competing interest.

### Author Declarations

Each study sample was collected with participants written informed consent approved by local Institutional Review Boards (IRB). The details of the IRB/oversight body that provided approval or exemption for the research described are given below. Ethical approval was given by the following boards/committees: Institute of Psychiatry Research Ethics Committee at King's College London, Institutional Review Board of University of Muenster, Institutional Review Board of Chinese Academy of Sciences, Institutional Review Board of Monash University Institutional Review Board of Philipps-University Marburg, Institutional Review Board of University of Bonn, Institutional Review Board of University of London, Institutional Review Board of University of Geneva, Institutional Review Board of University of Auckland, Ethics Committee of the Mental Health Research Center, Moscow, Institutional Review Board of McGill University, Institutional Review Board of University of Roehampton, Institutional Review Board of Jena University Hospital, Institutional Review Board of Donald and Barbara Zucker School of Medicine at Hofstra University/Northwell, Institutional Review Board of University of Amsterdam, Ethic Commission Zurich, Institutional Review Board of Brunel University London, Institutional Review Board of University of Groningen, Human Research Ethics Committee at University of Melbourne.

