## Supplement for "Cortical and Subcortical Neuroanatomical Signatures of Schizotypy in 3,004 Individuals Assessed in a Worldwide ENIGMA Study"

---

### Index

#### Supplementary Figures


- Figure S1: Location of Enigma Schizotypy Working Group Members


#### Site Information and Demographics


- Table S1: Scanner Details
- Table S2: Demographics Cortical Meta-Analysis
- Table S3: Demographics Subcortical Meta-Analysis


#### Supplementary Methods

#### Brain Region Mean Values


- Table S4: Mean Cortical Thickness and Surface Area
- Table S5: Mean Subcortical Volume


#### Cortical Thickness Models


- Table S6: Cortical Thickness Continuous Model
- Table S7: Cortical Thickness Continuous Model - No Thickness Covariate


#### Cortical Surface Models


- Table S8: Cortical Surface Area Continuous Model
- Table S9: Cortical Surface Area Continuous Model - No Surface Area Covariate


#### Moderator Analysis - Coritcal Thickness Continuous Models


- Table S10: Schizotypy Questionaire Moderator
- Table S11: Scanner Number Moderator
- Table S12: FreeSurfer Version Moderator


#### Subcortical Models


- Table S13: Subcortical Correlation Model


#### Smoking Status - Cortical Thickness


- Table S14: Effect of Schizotypy in Subgroup with Smoking Data
- Table S15: Effect of Schizotypy Controlling for Smoking Status


#### Smoking Status - Subcortical Volume


- Table S16: Effect of Schizotypy in Subgroup with Smoking Data
- Table S17: Effect of Schizotypy Controlling for Smoking Status


#### Measures of Schizotypy


- Table S18: Schizotypy Scales


#### Funding and Acknowledgments

#### Supplementary Figures

##### Figure S1: Location of Enigma Schizotypy Working Group Members

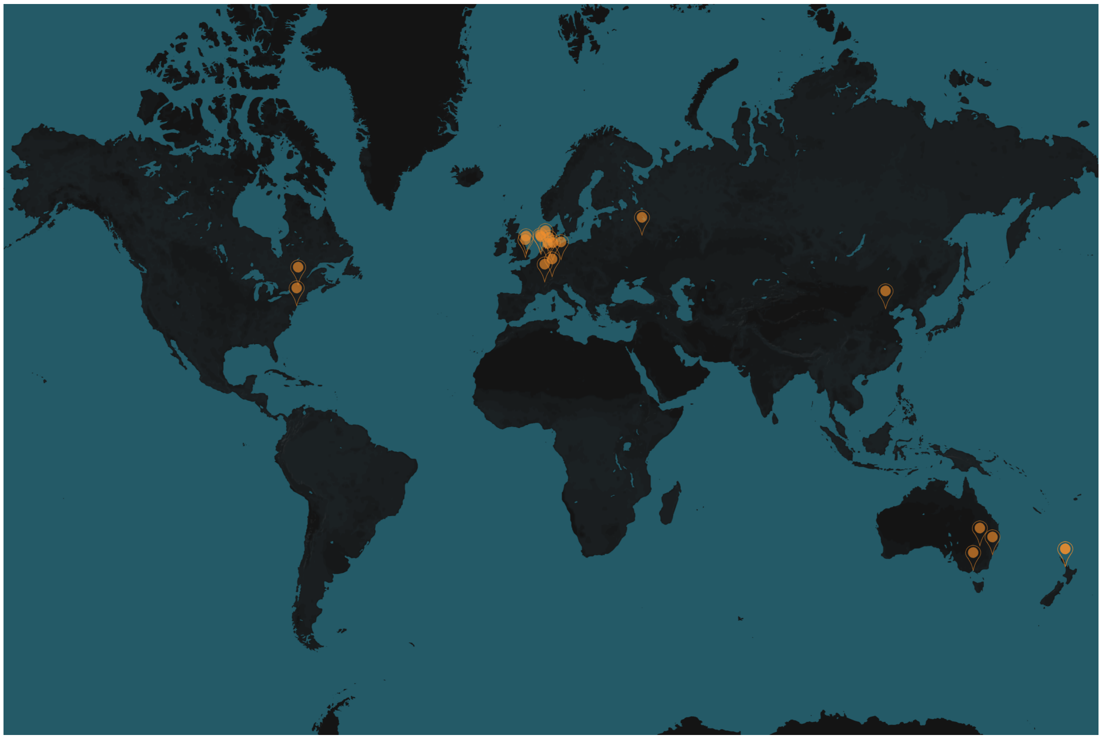

Location of Enigma Schizotypy Working Group Members

#### Site Information and Demographics

##### Table S1: Scanner Details

| Sample Abbreviation | Sample Name | Number of Scanners | Scanner Type | Imaging protocols | Slice Orientation | FreeSurfer Version | Operating System |
|---|---|---|---|---|---|---|---|
| AMS | Amsterdam | 1 | 3T Phillips Achieva | 3D T1-weighted images (TR = 8.2, TE = 3.8, FA = 8°, FOV 240 × 188 mm, voxel size 1 × 1 × 1, 220 slices) | sequential ascending | 6.0.1 | Virtual linux environmen (singularity on HPC) |
| ASRB | Sydney 1 | 5 | 1.5T Siemens Avanto | 3D MPRAGE; TR 1980ms, TE 4.3ms, field-of-view 250 x 250 mm2, data acquisition matrix 256 x 256, 176 contiguous 1mm slices; voxel size 0.98 x 0.98 x 1.0 mm3, flip angle 15° | sagittal | 5.1 | Mac OS X 10.9.5 |
| AUCK | Aukland | 1 | 3T Siemens | 3D MPRAGE; FOV = 256 mm2; matrix size = 256x 256 mm; number of slices = 176; slice thickness = 1 mm; voxel-size = 1 x1 x1 mm3; TE/TR/flip angle = 2.07 ms/1900 ms/9deg; GRAPPA acceleration factor = 2; TA = 4.26 min. | interleaved | 5.3.0 | Linux |
| BEIJ | Beijing | 2 | 3T Siemens | site 1: MPRAGE; TR 2530ms, TE 2.34ms, FOV 256mm, matrix 256x256, 192 slices, slice thickness 1mm, flip angle 7; site 2: MPRAGE; TR 2530ms, TE 2.34ms, FOV 256mm, matrix 256x256, 172 slices, slice thickness 1mm, flip angle 7; | sagittal | 6.0.0 | Linux |
| BONNOLIFE | Bonn O-Life | 1 | 3T Siemens Trio | TR = 1570 ms; TE = 3.42 ms; inversion time (TI) = 800 ms; flip angle = 15°; FoV = 256 mm; matrix size = 256 x 256; 160 slices; slice thickness = 1 mm; voxel size = 1 x 1 x 1 | sagittal | 6.0.0 | Virtual linux environmen (singularity on HPC) |

| Sample Abbreviation | Sample Name | Number of Scanners | Scanner Type | Imaging protocols | Slice Orientation | FreeSurfer Version | Operating System |
|---|---|---|---|---|---|---|---|
| BONNRISC | Bonn RISC | 2 | 1.5 T GE Signa Advantage, 3T Siemens Magnetom Verio | GE Signa Advantage: 3D-SPGR voxel resolution: 1 x 1 x 1.5 mm; repetition time (TR): 18 ms; inversion time (TI): 450 ms; echo time (TE): 5.1 ms; bandwidth: 15.63 kHz | sagittal | 6.0.0 | Virtual linux environmen (singularity on HPC) |
| BONNSPQ | Bonn SPQ | 1 | 3T Siemens Magnetom Verio | 3D MPRAGE sequence, repetition time of TR = 2400 ms, echo time TE = 3.06 ms, flip angle = 9 degrees with 160 slices, slice thickness = 1.0 mm, voxel size = 1.0 × 1.0 × 1.0 mm, field of view FOV = 256 mm | sagittal | 6.0.0 | Virtual linux environmen (singularity on HPC) |
| CAM | Cambridge | 1 | 3T Siemens Trio | MPRAGE; TR/TE 2.98/2300 ms, 1 × 1 voxels, slice thickness 1 mm, flip angle 9°, FOV 24 × 25.6 mm, 176 slices | saggital | 6.0.0 | Unix |
| DECOP | London 4 | 1 | 3T GE | T1-weighted image (196 slices; isotropic voxels of 1.2 mm; TR 7.312 ms; TE 3.016 ms;flip angle: 11°; FOV 270 mm) | sagittal | 5.3 | Red Hat Enterprise Linux Serve release 5.1 (Tikanga) |
| FOR2107-MR | Marburg | 1 | 3T Siemens Magnetom TiroTim syngo | 3D T1-weighted magnetization prepared rapid acquisition gradient echo (MPRAGE); TR=1900ms, TE=2.26ms, TI=900ms, FA=9°, voxel size=1.0x1.0x1.0mm³, Acquisition Direction Sagittal, 176 slices, slice gap 0.5mm. | sagittal | 5.3 | Red Hat Enterprise Linux Serve release 5.1 (Tikanga) |
| FOR2107-MS | Muenster | 1 | 3T Siemens PRISMA | Site Muenster: 3D T1-weighted magnetization prepared rapid acquisition gradient echo (MPRAGE); TR=2130ms, TE=2.28ms, TI=900ms, FA=8°, voxel size=1.0x1.0x1.0mm³, Acquisition Direction Sagittal, 192 slices, no slice gap. | descending (anterior to posterior) | 6.0.0 | Mac OS SIERRA v 10.12.6 |
| GENEVA | Geneva | 2 | 3T Siemens Trio | T1-Weighted images (192 slices, TR=2500ms, TE=3ms, flip angle=( degree, slice hickness=1.1mm, FOV=22cm, Acq matrix 256x256) | descending | 6.0.1 | Virtual linux environmen (singularity on HPC) |
| GRON1 | Groningen 1 | 1 | 3T Phillips | a T1-weighted image was obtained (TR/TE?=?9/3.5?ms) using fast-field echo and turbo-field echo: 170 axial slices; FOV (rl, ap, fh)?=?232?×?170?×?256 mm; flip angle?=?8°, voxel size?=?1?×?1?×?1 mm, slice thickness?=?1 mm. | descending | 6.0.1 | Virtual linux environmen (singularity on HPC) |
| GRON2 | Groningen 2 | 1 | 3T Phillips | anatomic images were obtained using a sagittal 3-dimensional T1-weighted sequence (176 slices, repetition time 9 ms, echo time 3.5 ms, field of view 256 mm, voxel size 1 × 1 × 1 mm, slice thickness 1.0 mm) using an 8 channel head coil. | sagittal | 5.3 | Mac OS X 10.9.5 |
| IGP | Sydney 2 | 1 | 3T Phillips Achieva TX | 3D MPRAGE; TR 8.9ms, TE 4.1ms, field of view 240mm, matrix 268 x 268, 200 slices, slice thickness 0.9mm, no gap | sagittal | 6.0.0 | Linux |

| Sample Abbreviation | Sample Name | Number of Scanners | Scanner Type | Imaging protocols | Slice Orientation | FreeSurfer Version | Operating System |
|---|---|---|---|---|---|---|---|
| JENA1 | Jena | 1 | 3T Siemens Tim Trio | 3D T1-weighted MPRAGE, TR: 2300ms, TE: 3.03 ms, flip angle: 9°, 192 slices, field of view 256mm, voxel resolution 1x1x1mm, acquisition time: 5:21min | sequential (top down) | 6.0.0 | Unix |
| LOND1a | London 1a | 1 | 3T GE Discovery MR750 | 3D T1-weighted inversion recovery prepared gradient echo sequence (voxel size: 1.05 × 1.05 × 1.2 mm, field of view: 270 mm, 196 slices, TR: 7.3 ms, TE: 3.0 ms, inversion time: 400 ms, flip angle = 11°), based on the well-validated ADNI 2/ADNI GO protocols (see http://adni.loni.usc.edu/methods/documents/mri-protocols/ (http://adni.loni.usc.edu/methods/documents/mri-protocols/)). | interleaved | 6.0.0 | Unix |
| LOND1b | London 1b | 1 | 3T Phillips Intera | T1-weighted 3D fast-field echo (FFE) sequence [repetition time (TR)=25 ms, echo time (TE)=4.6 ms, field of view (FOV)=260 mm, matrix=256x256, 160 contiguous axial slices of 1-mm thickness, voxel size= 1x1x1 mm]. | axial | 6.0.0 | Unix |
| LOND2 | London 2 | 2 | 1.5 T GE, 3T GE | 1.5 T GE: 3D, TR=11.1 ms, TE=4.9 ms, inversion time=300 ms, acquisition matrix=256 x 160, 150 locations, slice thickness=1.1 mm, in-plane resolution=1.094 mm, flip angle=18°; 3 T GE: TR: 7.3 ms, TE: 3.0 ms, inversion time: 400 ms, voxel size: 1.05 x1.05 x1.2mm, field of view: 270mm, 196 slices, flip angle = 11° | interleaved | 6.0.0 | Mac |
| LOND5 | London 5 | 1 | 3T Siemens | MPRAGE; TR 1900ms; 1mm x 1mm 1mm voxel size; in plane resolution of 256 x 256 x 176 slices, flip angle 11, slice thickness 1mm, TE = 2.07ms | sequential (top down) | 6.0.0 | Unix |
| LONDRoe | London 3 | 1 | 3T Siemens | MPRAGE; TR 2000ms; 1mm x 1mm 1mm. 256 x 256 x 176 slices, flip angle 11, slice thickness 1mm, TE = 2.07ms | interleaved | 6.0.0 | Linux Ubun Bionic |
| MELB | Montreal | 1 | 3T Siemens | 3D magnetic-prepared rapid gradient echo sequence. A total of 192 slices were acquired for each participant's T1-weighted images using an ascending acquisition with the following parameters: TR of 2300 ms, TE of 2.07 ms, flip angle of 9°, FOV of 256mm, and voxel size of 1mm3. | sagittal | 5.3.0 | Linux |
| MNC | Melbourne | 1 | 3T scanner Gyroscan Intera, Philips Medical Systems | 3D fast gradient echo sequence (turbo field echo), TR = 7.4 msec, TE = 3.4 msec, Flip Angle = 9°, two signal averages, inversion prepulse every 814.5 msec, acquired over a FOV of 256 (feet-head [FH]) × 204 (anterior-posterior [AP]) × 160 (right-left [RL]) mm, phase encoding in AP and RL direction, reconstructed to cubic voxels of .5 × .5 × .5 mm | sagittal | 5.3 | Red Hat Enterprise Linux Serve release 5.1 (Tikanga) |
| MOSC | Moscow | 1 | 3 T Phillips Ingenia | 3D T1-weighted TFE; TR=7.9 ms, TE=3.5 ms; flip angle 8; number of slices=170; voxel size=0.98 x 0.98 x 1.0, no gap | axial | 5.3 | LinuxCentC 6.10 |

| Sample Abbreviation | Sample Name | Number of Scanners | Scanner Type | Imaging protocols | Slice Orientation | FreeSurfer Version | Operating System |
|---|---|---|---|---|---|---|---|
| MTL | Muenster Neuroimaging Cohort | 1 | 3T Siemens Trio | 3D T1-weighted images were acquired with a gradient echo T1-weighted sequence, voxel size 1x1x1 mm3. TR=1900ms; TE=4.9ms; FA=25; matrix 176x256x256 | saggital | 6.0.0 | Unix |
| NYC | New York | 1 | 3T GE | 3D-SPGR images using a 1mm thick slice acquisition with image parameters: TR = 7.5 ms, TE = 3 ms, matrix = 256x256, FOV = 240 mm, 216 contiguous images. | interleaved | 6.0.0 | Linux |
| UTR | Utretch | 1 | 3T Phillips Achieva | T1-weighted images were acquired with a 3D T1 turbo field-echo sequence. The acquisition protocol parameters were: 160 slices; repetition time = 9.96 ms; echo time = 4.59 ms; flip angle 8°; 1 mm slice thickness with no inter-slice gap; matrix = 256 × 256 and field of view 224 mm, achieving a voxel size of 0.875 × 0.875 × 1 mm. Scan time was 8 min and 50 s | saggital | 5.1.0 | Unix |
| ZUR1 | Zurich 1 | 1 | 3T Phillips | 3D T1-weighted images were acquired with an ultrafast gradient echo T1-weighted sequence (TR=8.4ms, TE=3.8ms, flip angle=8°) in 160 sagittal plan slices (1mm slice thickness, no slice gap) of 240×240mm2 resulting in 1x1x1mm3 voxels. | sagittal | 6.0.0 | Unix |
| ZUR2 | Zurich 2 | 1 | 3T Phillips Achieva | 3D T1-weighted images; 160 slices; TR, 8.2 ms; TE, 3.8 ms; flip angle, 8°; spatial resolution, 1 × 1 × 1 mm3; FOV = 160 × 240 mm2 | sagittal | 6.0.0 | OS X |

Table S2: Demographics Cortical Meta-Analysis

| Site | N | Scale | Female/Male | Mean Age | Age S.D. | Age Range | Mean SPT score | Score S.D. | Score range |
|---|---|---|---|---|---|---|---|---|---|
| AMS | 49 | CAPE | 23/26 | 21.36 | 3.08 | 16.18-32.24 | 64.33 | 10.19 | 46-86 |
| ASRB | 191 | SPQ | 91/100 | 39.69 | 13.77 | 18-65 | 10.24 | 9.11 | 0-43 |
| AUK | 49 | OLIFE | 32/17 | 23.33 | 4.5 | 18-38 | 42.71 | 17.85 | 10-81 |
| BEI | 146 | SPQ | 77/69 | 19.57 | 1.08 | 16-24 | 23.19 | 12.67 | 2-53 |
| BONNOLIFE | 31 | OLIFE | 13/18 | 23.61 | 4.52 | 19-43 | 11.48 | 8.04 | 1-24 |
| BONNRISC | 51 | RISC | 18/33 | 27.2 | 5.36 | 19-40 | 22.37 | 7.71 | 2-34 |
| BONNSPQ | 92 | SPQ | 46/46 | 28.16 | 8.01 | 19-50 | 8.4 | 7.19 | 0-35 |
| CAM | 89 | SPQ | 48/41 | 24.07 | 5.3 | 1-41 | 16.17 | 10.13 | 0-43 |
| DECOP | 26 | CAPE | 09/17 | 38.62 | 7.34 | 28-54 | 66.73 | 12.78 | 50-101 |
| FOR2107-MR | 446 | SPQB | 279/167 | 34.67 | 12.85 | 18-65 | 3.49 | 3.02 | 0-16 |
| FOR2107-MS | 228 | SPQB | 146/82 | 28.42 | 10.41 | 18-65 | 3.04 | 2.96 | 0-16 |
| GENEVA | 118 | SPQ | 58/60 | 17.02 | 4.22 | 12.06-33.76 | 19.6 | 12.54 | 1-57 |
| GRON1 | 42 | SPQ | 16/26 | 22.67 | 2.26 | 18-27 | 33.64 | 5.22 | 25-44 |
| GRON2 | 83 | CAPE-Adj | 47/36 | 32.43 | 9.53 | 19-55 | 0.96 | 1.29 | 0-4.12 |
| IGPS | 63 | SPQ | 28/35 | 36.43 | 11.05 | 20.15-59.03 | 12.21 | 11.12 | 0-46 |

| Site | N | Scale | Female/Male | Mean Age | Age S.D. | Age Range | Mean SPT score | Score S.D. | Score range |
|---|---|---|---|---|---|---|---|---|---|
| JENA | 115 | SPQ | 60/55 | 29.37 | 9.65 | 20-60 | 10.76 | 8.21 | 0-49 |
| LOND1a | 46 | OLIFE | 22/24 | 27.46 | 6.84 | 18-44 | 26.52 | 15.48 | 4-58 |
| LOND1b | 40 | CAPE | 19/21 | 20.8 | 3.76 | 18-39 | 1.44 | 0.34 | 1-2.15 |
| LOND5 | 44 | SPQ | 32/12 | 20.09 | 1.7 | 18-27 | 26.55 | 20.54 | 0-63 |
| LOND3 | 40 | SPQ | 16/24 | 30.97 | 10.87 | 19-51 | 7.86 | 5.12 | 0-19 |
| MELB | 392 | CAPE | 224/168 | 23.38 | 5.17 | 18-50 | 25.43 | 4 | 20-38 |
| MTL | 40 | CHAP | 27/13 | 19.82 | 1.14 | 18.1-22.8 | 24.73 | 14.41 | 4-55 |
| MOS | 33 | SPQ | 16/17 | 23.83 | 3.35 | 18.1-30.1 | 15 | 11.84 | 1-45 |
| MNC | 173 | SPQB | 92/81 | 43.31 | 11.85 | 18-64 | 3.34 | 3.08 | 0-21 |
| NYC | 225 | CAPE-Adj | 110/115 | 35.92 | 12.85 | 18.47-68.04 | 1.14 | 0.16 | 1-1.84 |
| UTR | 97 | SPQ | 62/35 | 41.26 | 14.26 | 18-65 | 17.62 | 14.29 | 0-57 |
| ZUR1 | 27 | SPQ | 10/17 | 29.15 | 10.35 | 19-48 | 42.56 | 7.06 | 34-60 |
| ZUR2 | 62 | SPQ | 0/62 | 27.74 | 4.74 | 20-38 | 18.5 | 10.9 | 2-46 |
| Overall | **3038** | | **560.01/247.25** | **29.89** | **4.3** | **12.06-67.78** | | | |

Table S3: Demographics Subcortical Meta-Analysis

| Site | N | Scale | Female/Male | Mean Age | Age S.D. | Age Range | Mean SPT score | Score S.D. | Score range |
|---|---|---|---|---|---|---|---|---|---|
| AMS | 49 | CAPE | 23/26 | 21.36 | 3.08 | 16.18-32.24 | 64.33 | 10.19 | 46-86 |
| ASRB | 191 | SPQ | 100/91 | 39.69 | 13.77 | 18-65 | 10.24 | 9.11 | 0-43 |
| AUK | 49 | OLIFE | 32/17 | 23.33 | 4.5 | 18-38 | 42.71 | 17.85 | 10-81 |
| BEI | 146 | SPQ | 77/69 | 19.57 | 1.08 | 16-24 | 23.19 | 12.67 | 2-53 |
| BONNOLIFE | 31 | OLIFE | 13/18 | 23.61 | 4.52 | 19-43 | 11.48 | 8.04 | 1-24 |
| BONNRISC | 55 | RISC | 18/37 | 26.73 | 5.5 | 18-40 | 22.35 | 7.59 | 2-34 |
| BONNSPQ | 118 | SPQ | 58/60 | 27.18 | 7.92 | 18-50 | 8.29 | 7.04 | 0-35 |
| CAM | 89 | SPQ | 48/41 | 24.29 | 4.7 | 18-41 | 16.17 | 10.13 | 0-43 |
| DECOP | 26 | CAPE | 9/17 | 38.62 | 7.34 | 28-54 | 66.73 | 12.78 | 50-101 |
| FOR2107-MR | 446 | SPQB | 279/167 | 34.67 | 12.85 | 18-65 | 3.49 | 3.02 | 0-16 |
| FOR2107-MS | 228 | SPQB | 146/82 | 28.42 | 10.41 | 18-65 | 3.04 | 2.96 | 0-16 |
| GENEVA | 118 | SPQ | 58/60 | 17.02 | 4.22 | 12.06-33.76 | 19.6 | 12.54 | 1-57 |
| GRON1 | 42 | SPQ | 16/26 | 22.67 | 2.26 | 18-27 | 33.64 | 5.22 | 25-44 |
| GRON2 | 38 | CAPE-Adj | 19/19 | 36.61 | 9.42 | 20-53 | 1.74 | 1.29 | 0-4.05 |
| IGPS | 63 | SPQ | 28/35 | 36.43 | 11.05 | 20.15-59.03 | 12.21 | 11.12 | 0-46 |
| JENA | 115 | SPQ | 60/55 | 29.37 | 9.65 | 20-60 | 10.76 | 8.21 | 0-49 |

| Site | N | Scale | Female/Male | Mean Age | Age S.D. | Age Range | Mean SPT score | Score S.D. | Score range |
|---|---|---|---|---|---|---|---|---|---|
| LOND1a | 46 | OLIFE | 22/24 | 27.46 | 6.84 | 18-44 | 26.52 | 15.48 | 4-58 |
| LOND1b | 40 | CAPE | 19/21 | 20.8 | 3.76 | 18-39 | 1.44 | 0.34 | 1-2.15 |
| LOND3 | 40 | SPQ | 16/24 | 30.97 | 10.87 | 19-51 | 7.86 | 5.12 | 0-19 |
| LOND5 | 30 | SPQ | 22/8 | 19.73 | 1.31 | 18-23 | 27.03 | 20.55 | 0-57 |
| MELB | 387 | CAPE | 221/166 | 23.38 | 5.15 | 18-50 | 25.38 | 3.92 | 20-38 |
| MTL | 40 | CHAP | 27/13 | 19.92 | 1.21 | 18-23 | 24.73 | 14.41 | 4-55 |
| MOS | 33 | SPQ | 16/17 | 23.83 | 3.35 | 18.1-30.1 | 15 | 11.84 | 1-45 |
| MNC | 173 | SPQB | 92/81 | 43.31 | 11.85 | 18-64 | 3.34 | 3.08 | 0-21 |
| NYC | 225 | CAPE-Adj | 110/115 | 35.92 | 12.85 | 18.47-68.04 | 1.14 | 0.16 | 1-1.84 |
| UTR | 97 | SPQ | 62/35 | 41.26 | 14.26 | 18-65 | 17.62 | 14.29 | 0-57 |
| ZUR1 | 27 | SPQ | 10/17 | 29.15 | 10.35 | 19-48 | 42.56 | 7.06 | 34-60 |
| ZUR2 | 62 | SPQ | 0/62 | 27.74 | 4.74 | 20-38 | 18.5 | 10.9 | 2-46 |
| Overall | **3004** | | **561.09/246.91** | **29.9** | **4.31** | **12.06-67.78** | | | |

### Supplementary Methods

### Brain Region Mean Values

Table S4: Mean Cortical Thickness and Surface Area

| | Thickness | | | | | Surface | | | | |
|---|---|---|---|---|---|---|---|---|---|---|
| **Region** | **N** | **Mean** | **S.D.** | **Min** | **Max** | **N** | **Mean** | **S.D.** | **Min** | **Max** |
| L_bankssts | 2788 | 2.54 | 0.09 | 1.93 | 2.66 | 2824 | 1053.87 | 50.74 | 495 | 1299 |
| L_caudalanteriorcingulate | 2943 | 2.75 | 0.12 | 1.97 | 2.71 | 2979 | 662.19 | 31.27 | 312 | 744 |
| L_caudalmiddlefrontal | 2945 | 2.63 | 0.08 | 2 | 2.68 | 2981 | 2362.07 | 90.72 | 1252 | 2957 |
| L_cuneus | 2815 | 1.89 | 0.09 | 1.28 | 1.75 | 2851 | 1534.11 | 79.68 | 700 | 1734 |
| L_entorhinal | 2781 | 3.3 | 0.19 | 1.87 | 3.46 | 2816 | 443.49 | 36.78 | 207 | 586 |
| L_fusiform | 2911 | 2.74 | 0.13 | 1.97 | 2.67 | 2947 | 3301.29 | 179.12 | 1811 | 3997 |
| L_inferiorparietal | 2856 | 2.51 | 0.08 | 1.91 | 2.4 | 2892 | 4694.9 | 186.41 | 2377 | 5829 |
| L_inferiortemporal | 2882 | 2.82 | 0.17 | 2 | 2.65 | 2918 | 3480.87 | 243.74 | 1581 | 4170 |
| L_isthmuscingulate | 2957 | 2.53 | 0.11 | 1.79 | 2.55 | 2993 | 1028.75 | 38.47 | 587 | 1258 |
| L_lateraloccipital | 2915 | 2.22 | 0.11 | 1.5 | 1.95 | 2951 | 5041 | 319.83 | 3310 | 5992 |
| L_lateralorbitofrontal | 2931 | 2.7 | 0.09 | 2.1 | 2.81 | 2967 | 2737.12 | 119.92 | 1446 | 3118 |

| | Thickness | | | | | Surface | | | | |
|---|---|---|---|---|---|---|---|---|---|---|
| **Region** | **N** | **Mean** | **S.D.** | **Min** | **Max** | **N** | **Mean** | **S.D.** | **Min** | **Max** |
| L_lingual | 2894 | 2.04 | 0.09 | 1.41 | 1.92 | 2930 | 3141.08 | 163.91 | 1571 | 3487 |
| L_medialorbitofrontal | 2915 | 2.48 | 0.11 | 1.86 | 2.45 | 2951 | 1896.48 | 107.96 | 1064 | 2248 |
| L_middletemporal | 2780 | 2.93 | 0.14 | 2.11 | 2.94 | 2816 | 3229.17 | 195.65 | 1725 | 3904 |
| L_parahippocampal | 2942 | 2.83 | 0.12 | 1.62 | 2.95 | 2977 | 708.66 | 36.88 | 432 | 828 |
| L_paracentral | 2954 | 2.45 | 0.1 | 1.81 | 2.26 | 2990 | 1370.63 | 54.69 | 806 | 1586 |
| L_parsopercularis | 2931 | 2.67 | 0.09 | 1.63 | 2.65 | 2967 | 1694.64 | 59.98 | 1032 | 1970 |
| L_parsorbitalis | 2935 | 2.8 | 0.11 | 1.89 | 2.71 | 2971 | 677.84 | 49.49 | 396 | 781 |
| L_parstriangularis | 2920 | 2.54 | 0.09 | 1.33 | 2.57 | 2956 | 1342.39 | 64.32 | 691 | 1718 |
| L_pericalcarine | 2868 | 1.63 | 0.11 | 1.09 | 1.58 | 2904 | 1428.78 | 77.26 | 776 | 1726 |
| L_postcentral | 2889 | 2.13 | 0.06 | 1.72 | 2.03 | 2925 | 4229.85 | 161.74 | 2762 | 4920 |
| L_posteriorcingulate | 2956 | 2.59 | 0.1 | 1.92 | 2.52 | 2990 | 1209.35 | 41.41 | 741 | 1469 |
| L_precentral | 2910 | 2.63 | 0.1 | 1.95 | 2.57 | 2945 | 4913.82 | 215.13 | 3315 | 5822 |
| L_precuneus | 2942 | 2.44 | 0.08 | 1.88 | 2.37 | 2978 | 3881.81 | 139.88 | 2551 | 4795 |
| L_rostralanteriorcingulate | 2929 | 2.94 | 0.12 | 1.96 | 2.94 | 2965 | 862.72 | 30.94 | 344 | 1129 |
| L_rostralmiddlefrontal | 2936 | 2.44 | 0.1 | 1.89 | 2.41 | 2972 | 5902.13 | 294.36 | 3692 | 6981 |
| L_superiorfrontal | 2936 | 2.82 | 0.1 | 2.14 | 2.67 | 2971 | 7446.57 | 313.1 | 4887 | 8644 |
| L_superiorparietal | 2895 | 2.24 | 0.07 | 1.65 | 2.18 | 2931 | 5524.96 | 194.19 | 3462 | 6325 |
| L_superiortemporal | 2767 | 2.87 | 0.11 | 2.25 | 3.04 | 2804 | 3920.64 | 182.28 | 2326 | 4589 |
| L_supramarginal | 2802 | 2.62 | 0.09 | 1.98 | 2.59 | 2838 | 4056.47 | 227.54 | 2192 | 4949 |
| L_frontalpole | 2962 | 2.85 | 0.14 | 1.66 | 3.01 | 2998 | 233.95 | 28.24 | 110 | 266 |
| L_temporalpole | 2911 | 3.58 | 0.22 | 1.77 | 3.8 | 2946 | 481.69 | 21.75 | 253 | 577 |
| L_transversetemporal | 2962 | 2.43 | 0.13 | 1.37 | 2.37 | 2998 | 466.68 | 21.39 | 273 | 587 |
| L_insula | 2889 | 3.1 | 0.11 | 2.31 | 2.99 | 2925 | 2328.64 | 139.5 | 1462 | 2466 |
| R_bankssts | 2893 | 2.66 | 0.1 | 1.71 | 2.71 | 2929 | 957.45 | 54.15 | 542 | 1132 |
| R_caudalanteriorcingulate | 2954 | 2.6 | 0.11 | 1.81 | 2.71 | 2989 | 770.34 | 40.67 | 329 | 978 |
| R_caudalmiddlefrontal | 2938 | 2.59 | 0.09 | 1.96 | 2.68 | 2974 | 2205.54 | 111.71 | 1188 | 2674 |
| R_cuneus | 2840 | 1.92 | 0.09 | 1.3 | 1.85 | 2876 | 1586.95 | 82.89 | 560 | 1931 |
| R_entorhinal | 2702 | 3.43 | 0.19 | 2.1 | 3.5 | 2736 | 388 | 32.51 | 114 | 477 |
| R_fusiform | 2918 | 2.76 | 0.15 | 1.82 | 2.71 | 2954 | 3227.61 | 166.49 | 1883 | 3809 |
| R_inferiorparietal | 2854 | 2.55 | 0.09 | 1.95 | 2.56 | 2891 | 5557.38 | 241.94 | 3172 | 6593 |
| R_inferiortemporal | 2892 | 2.84 | 0.18 | 1.65 | 2.79 | 2928 | 3325.58 | 221.94 | 1412 | 4042 |
| R_isthmuscingulate | 2946 | 2.51 | 0.09 | 1.86 | 2.6 | 2981 | 937.19 | 29.27 | 530 | 1146 |
| R_lateraloccipital | 2909 | 2.28 | 0.12 | 1.52 | 2.13 | 2945 | 4946.39 | 333.68 | 2909 | 5814 |

| | Thickness | | | | | Surface | | | | |
|---|---|---|---|---|---|---|---|---|---|---|
| Region | N | Mean | S.D. | Min | Max | N | Mean | S.D. | Min | Max |
| R_lateralorbitofrontal | 2907 | 2.66 | 0.1 | 1.94 | 2.58 | 2943 | 2701.92 | 138.52 | 1519 | 3259 |
| R_lingual | 2895 | 2.08 | 0.09 | 1.55 | 2.01 | 2931 | 3194.39 | 165.49 | 1816 | 3708 |
| R_medialorbitofrontal | 2895 | 2.47 | 0.12 | 1.83 | 2.49 | 2930 | 1916.35 | 100.58 | 1090 | 2242 |
| R_middletemporal | 2864 | 2.96 | 0.13 | 2.06 | 2.95 | 2902 | 3560.95 | 195.57 | 2133 | 4085 |
| R_parahippocampal | 2944 | 2.79 | 0.12 | 1.91 | 2.92 | 2980 | 685.3 | 41.3 | 341 | 797 |
| R_paracentral | 2951 | 2.47 | 0.11 | 1.85 | 2.29 | 2987 | 1545.79 | 63.41 | 987 | 1723 |
| R_parsopercularis | 2908 | 2.66 | 0.08 | 2.03 | 2.64 | 2944 | 1423.51 | 55.76 | 829 | 1690 |
| R_parsorbitalis | 2933 | 2.77 | 0.1 | 1.65 | 2.79 | 2969 | 828.11 | 47.05 | 510 | 1008 |
| R_parstriangularis | 2899 | 2.52 | 0.09 | 1.85 | 2.5 | 2935 | 1544.28 | 75.87 | 895 | 1800 |
| R_pericalcarine | 2850 | 1.63 | 0.11 | 1.12 | 1.56 | 2886 | 1564.36 | 80.51 | 692 | 1855 |
| R_postcentral | 2904 | 2.1 | 0.06 | 1.69 | 2.14 | 2941 | 4068.26 | 152.31 | 2493 | 4747 |
| R_posteriorcingulate | 2957 | 2.56 | 0.09 | 1.95 | 2.59 | 2991 | 1232.3 | 46.22 | 702 | 1457 |
| R_precentral | 2908 | 2.58 | 0.09 | 1.94 | 2.54 | 2944 | 4950.88 | 209.46 | 3359 | 5707 |
| R_precuneus | 2941 | 2.45 | 0.08 | 1.88 | 2.42 | 2977 | 4059.22 | 153.04 | 2584 | 4856 |
| R_rostralanteriorcingulate | 2904 | 2.91 | 0.12 | 1.94 | 3.05 | 2940 | 672.22 | 49.95 | 273 | 797 |
| R_rostralmiddlefrontal | 2914 | 2.37 | 0.11 | 1.88 | 2.26 | 2950 | 6094.3 | 304.84 | 3918 | 7580 |
| R_superiorfrontal | 2935 | 2.78 | 0.1 | 2.13 | 2.73 | 2971 | 7218.26 | 305.46 | 4387 | 7915 |
| R_superiorparietal | 2911 | 2.23 | 0.07 | 1.71 | 2.17 | 2947 | 5501.62 | 204.91 | 3317 | 6671 |
| R_superiortemporal | 2842 | 2.9 | 0.1 | 2.19 | 2.96 | 2878 | 3705.84 | 162.91 | 2382 | 4175 |
| R_supramarginal | 2829 | 2.63 | 0.09 | 1.98 | 2.57 | 2865 | 3770.22 | 144.57 | 2139 | 4604 |
| R_frontalpole | 2953 | 2.8 | 0.14 | 1.57 | 2.95 | 2988 | 301.61 | 21.78 | 140 | 367 |
| R_temporalpole | 2848 | 3.67 | 0.24 | 1.75 | 4 | 2884 | 442.52 | 27.47 | 204 | 538 |
| R_transversetemporal | 2965 | 2.46 | 0.15 | 1.42 | 2.46 | 3001 | 350.23 | 16.54 | 167 | 425 |
| R_insula | 2856 | 3.08 | 0.13 | 2.31 | 3.03 | 2892 | 2331.06 | 106.23 | 1449 | 2653 |
| LThickness | 2967 | 2.54 | 0.08 | 2.08 | 2.36 | 3003 | 12.71 | 48.22 | 2.08 | 2.36 |
| RThickness | 2967 | 2.54 | 0.08 | 2.04 | 2.4 | 3003 | 13.31 | 46.87 | 2.04 | 2.4 |
| LSurfArea | 2967 | 87267.07 | 3148.57 | 59365.8 | 97806.3 | 3003 | 87333.3 | 3265.94 | 59365.8 | 97806.3 |
| RSurfArea | 2967 | 87544.61 | 3106.6 | 59252 | 96576.2 | 3003 | 87608.73 | 3224.42 | 59252 | 96576.2 |

Table S5: Mean Subcortical Volume

| Region | N | Mean | S.D. | Min | Max |
|---|---|---|---|---|---|
| LLatVent | 2997 | 7456.26 | 1053.33 | 1369.30 | 10534.10 |

| Region | N | Mean | S.D. | Min | Max |
|---|---|---|---|---|---|
| RLatVent | 2997 | 6978.64 | 1023.95 | 1264.90 | 12906.20 |
| Lthal | 2973 | 7953.10 | 482.56 | 4685.70 | 8519.10 |
| Lcaud | 2988 | 3761.75 | 197.85 | 2036.90 | 4058.80 |
| Lput | 2941 | 5478.50 | 458.59 | 2175.20 | 4843.50 |
| Lpal | 2769 | 1748.65 | 299.01 | 419.10 | 982.60 |
| Lhippo | 2977 | 4291.68 | 238.80 | 2098.30 | 4625.80 |
| Lamyg | 2979 | 1673.98 | 100.81 | 1044.40 | 1778.70 |
| Laccumb | 2972 | 592.75 | 169.58 | 141.20 | 409.60 |
| Rthal | 2993 | 7357.46 | 572.20 | 2729.20 | 4381.60 |
| Rcaud | 2989 | 3796.49 | 269.10 | 1447.00 | 2487.30 |
| Rput | 2972 | 5309.39 | 564.15 | 1125.50 | 1922.70 |
| Rpal | 2977 | 1814.57 | 670.70 | 723.00 | 2003.20 |
| Rhippo | 2985 | 4407.02 | 226.90 | 2484.00 | 4867.30 |
| Ramyg | 2967 | 1789.44 | 284.99 | 1074.00 | 2024.30 |
| Raccumb | 2970 | 610.73 | 65.95 | 260.70 | 637.50 |
| Mvent | 2996 | 7212.76 | 1027.45 | 1471.70 | 11361.40 |
| Mthal | 2970 | 7655.74 | 459.63 | 4568.80 | 6598.60 |
| Mcaud | 2981 | 3777.99 | 141.06 | 2185.80 | 4088.90 |
| Mput | 2927 | 5391.41 | 485.77 | 1994.60 | 3327.75 |
| Mpal | 2764 | 1792.46 | 350.06 | 833.05 | 1930.80 |
| Mhippo | 2973 | 4349.02 | 217.64 | 2627.85 | 4689.60 |
| Mamyg | 2957 | 1730.82 | 175.00 | 1113.55 | 1908.60 |
| Maccumb | 2951 | 601.66 | 103.30 | 231.25 | 507.95 |
| ICV | 2999 | 1533873.22 | 68257.29 | 769523.00 | 1710000.00 |

### Cortical Thickness Models

##### Note - Measures of heterogeneity

$Tau^2$: Represents the absolute value of the true variance of the effect sizes, between-study variance

The $I^2$ statistic: Represents the proportion (in percentage) of total variation in the estimates that can be attributed to heterogeneity among the effects rather than sampling variability. The $I^2$ may be interpreted according to the following ranges $I^2$ = 0-40%: low heterogeneity, $I^2$ = 30-60%: moderate heterogeneity, $I^2$ = 50-90%: substantial heterogeneity, $I^2$ = 75-100%: considerable heterogeneity

The $H^2$ statistic: Represents the ratio of the total amount of variability in the effect size estimates to the amount of sampling variability

Table S6: Cortical Thickness Continuous Model

| Region | N | Effect Size (r) | SE | Lower CI | Upper CI | $tau^2$ | $I^2$ | $H^2$ | pvalue | FDRp |
|---|---|---|---|---|---|---|---|---|---|---|
| L_bankssts_thickavg | 2746 | 0.019 | 0.031 | -0.041 | 0.079 | 0.012 | 54.381 | 2.192 | 0.5310 | 0.981 |
| L_caudalanteriorcingulate_thickavg | 2884 | 0.019 | 0.019 | -0.018 | 0.055 | 0.000 | 0.000 | 1.000 | 0.3090 | 0.910 |
| L_caudalmiddlefrontal_thickavg | 2886 | 0.007 | 0.034 | -0.060 | 0.073 | 0.018 | 65.196 | 2.873 | 0.8400 | 0.981 |
| L_cuneus_thickavg | 2777 | 0.002 | 0.023 | -0.044 | 0.048 | 0.003 | 24.046 | 1.317 | 0.9360 | 0.981 |
| L_entorhinal_thickavg | 2755 | -0.019 | 0.019 | -0.057 | 0.018 | 0.000 | 0.000 | 1.000 | 0.3080 | 0.910 |
| L_fusiform_thickavg | 2870 | -0.038 | 0.020 | -0.078 | 0.002 | 0.001 | 9.521 | 1.105 | 0.0620 | 0.555 |
| L_inferiorparietal_thickavg | 2828 | -0.004 | 0.019 | -0.041 | 0.033 | 0.000 | 0.000 | 1.000 | 0.8360 | 0.981 |
| L_inferiortemporal_thickavg | 2843 | -0.003 | 0.019 | -0.039 | 0.034 | 0.000 | 0.000 | 1.000 | 0.8840 | 0.981 |
| L_isthmuscingulate_thickavg | 2898 | -0.023 | 0.028 | -0.077 | 0.032 | 0.009 | 47.412 | 1.902 | 0.4140 | 0.981 |
| L_lateraloccipital_thickavg | 2866 | 0.035 | 0.023 | -0.010 | 0.080 | 0.003 | 23.738 | 1.311 | 0.1240 | 0.844 |
| L_lateralorbitofrontal_thickavg | 2873 | 0.006 | 0.024 | -0.042 | 0.054 | 0.005 | 31.913 | 1.469 | 0.8020 | 0.981 |
| L_lingual_thickavg | 2842 | -0.013 | 0.019 | -0.050 | 0.024 | 0.000 | 1.786 | 1.018 | 0.4870 | 0.981 |
| L_medialorbitofrontal_thickavg | 2862 | 0.057 | 0.020 | 0.018 | 0.096 | 0.001 | 8.255 | 1.090 | 0.0040 | 0.120 |
| L_middletemporal_thickavg | 2756 | -0.002 | 0.030 | -0.061 | 0.057 | 0.011 | 52.553 | 2.108 | 0.9430 | 0.981 |
| L_parahippocampal_thickavg | 2888 | 0.012 | 0.028 | -0.042 | 0.066 | 0.008 | 45.968 | 1.851 | 0.6580 | 0.981 |
| L_paracentral_thickavg | 2897 | -0.046 | 0.022 | -0.089 | -0.003 | 0.002 | 19.606 | 1.244 | 0.0350 | 0.477 |
| L_parsopercularis_thickavg | 2875 | -0.012 | 0.019 | -0.048 | 0.025 | 0.000 | 0.000 | 1.000 | 0.5280 | 0.981 |
| L_parsorbitalis_thickavg | 2885 | 0.010 | 0.019 | -0.026 | 0.046 | 0.000 | 0.000 | 1.000 | 0.5910 | 0.981 |
| L_parstriangularis_thickavg | 2873 | -0.012 | 0.032 | -0.075 | 0.051 | 0.015 | 60.455 | 2.529 | 0.7160 | 0.981 |
| L_pericalcarine_thickavg | 2811 | -0.019 | 0.026 | -0.070 | 0.032 | 0.006 | 37.466 | 1.599 | 0.4700 | 0.981 |
| L_postcentral_thickavg | 2838 | -0.004 | 0.022 | -0.047 | 0.039 | 0.002 | 17.479 | 1.212 | 0.8610 | 0.981 |
| L_posteriorcingulate_thickavg | 2896 | 0.027 | 0.018 | -0.009 | 0.064 | 0.000 | 0.000 | 1.000 | 0.1400 | 0.844 |
| L_precentral_thickavg | 2855 | -0.010 | 0.019 | -0.046 | 0.027 | 0.000 | 0.000 | 1.000 | 0.5920 | 0.981 |
| L_precuneus_thickavg | 2887 | 0.001 | 0.022 | -0.042 | 0.044 | 0.002 | 18.010 | 1.220 | 0.9570 | 0.981 |
| L_rostralanteriorcingulate_thickavg | 2875 | 0.036 | 0.019 | -0.001 | 0.072 | 0.000 | 0.000 | 1.000 | 0.0550 | 0.555 |
| L_rostralmiddlefrontal_thickavg | 2886 | 0.004 | 0.020 | -0.036 | 0.043 | 0.001 | 9.134 | 1.101 | 0.8460 | 0.981 |
| L_superiorfrontal_thickavg | 2879 | -0.003 | 0.019 | -0.039 | 0.033 | 0.000 | 0.000 | 1.000 | 0.8720 | 0.981 |
| L_superiorparietal_thickavg | 2851 | -0.019 | 0.020 | -0.058 | 0.019 | 0.001 | 4.898 | 1.052 | 0.3200 | 0.910 |
| L_superiortemporal_thickavg | 2733 | 0.001 | 0.027 | -0.053 | 0.054 | 0.008 | 42.226 | 1.731 | 0.9790 | 0.981 |
| L_supramarginal_thickavg | 2775 | -0.022 | 0.021 | -0.063 | 0.020 | 0.001 | 12.048 | 1.137 | 0.3060 | 0.910 |
| L_frontalpole_thickavg | 2902 | 0.046 | 0.020 | 0.007 | 0.084 | 0.001 | 7.506 | 1.081 | 0.0210 | 0.382 |
| L_temporalpole_thickavg | 2864 | 0.019 | 0.019 | -0.018 | 0.055 | 0.000 | 0.000 | 1.000 | 0.3140 | 0.910 |
| L_transversetemporal_thickavg | 2903 | 0.035 | 0.018 | -0.001 | 0.071 | 0.000 | 0.000 | 1.000 | 0.0600 | 0.555 |

| Region | N | Effect Size (r) | SE | Lower CI | Upper CI | $tau^2$ | $I^2$ | $H^2$ | pvalue | FDRp |
|---|---|---|---|---|---|---|---|---|---|---|
| L_insula_thickavg | 2829 | -0.002 | 0.022 | -0.045 | 0.041 | 0.002 | 18.872 | 1.233 | 0.9280 | 0.981 |
| R_bankssts_thickavg | 2840 | 0.034 | 0.031 | -0.027 | 0.096 | 0.014 | 58.338 | 2.400 | 0.2740 | 0.910 |
| R_caudalanteriorcingulate_thickavg | 2897 | 0.025 | 0.026 | -0.026 | 0.077 | 0.007 | 40.506 | 1.681 | 0.3320 | 0.919 |
| R_caudalmiddlefrontal_thickavg | 2884 | 0.003 | 0.019 | -0.033 | 0.039 | 0.000 | 0.000 | 1.000 | 0.8690 | 0.981 |
| R_cuneus_thickavg | 2793 | 0.004 | 0.019 | -0.033 | 0.041 | 0.000 | 0.000 | 1.000 | 0.8420 | 0.981 |
| R_entorhinal_thickavg | 2695 | 0.007 | 0.020 | -0.032 | 0.046 | 0.001 | 4.544 | 1.048 | 0.7350 | 0.981 |
| R_fusiform_thickavg | 2877 | -0.001 | 0.024 | -0.047 | 0.045 | 0.004 | 27.295 | 1.375 | 0.9730 | 0.981 |
| R_inferiorparietal_thickavg | 2828 | 0.024 | 0.028 | -0.031 | 0.079 | 0.009 | 46.709 | 1.876 | 0.4000 | 0.981 |
| R_inferiortemporal_thickavg | 2855 | -0.026 | 0.025 | -0.076 | 0.023 | 0.006 | 35.851 | 1.559 | 0.2980 | 0.910 |
| R_isthmuscingulate_thickavg | 2890 | -0.039 | 0.026 | -0.091 | 0.013 | 0.007 | 41.837 | 1.719 | 0.1370 | 0.844 |
| R_lateraloccipital_thickavg | 2868 | 0.009 | 0.030 | -0.050 | 0.069 | 0.012 | 55.610 | 2.253 | 0.7570 | 0.981 |
| R_lateralorbitofrontal_thickavg | 2853 | 0.005 | 0.031 | -0.055 | 0.065 | 0.013 | 55.888 | 2.267 | 0.8810 | 0.981 |
| R_lingual_thickavg | 2843 | -0.019 | 0.028 | -0.075 | 0.036 | 0.009 | 48.214 | 1.931 | 0.4960 | 0.981 |
| R_medialorbitofrontal_thickavg | 2847 | 0.067 | 0.019 | 0.031 | 0.103 | 0.000 | 0.000 | 1.000 | 0.0001 | 0.017 |
| R_middletemporal_thickavg | 2832 | 0.031 | 0.031 | -0.030 | 0.092 | 0.013 | 57.434 | 2.349 | 0.3170 | 0.910 |
| R_parahippocampal_thickavg | 2897 | -0.003 | 0.023 | -0.048 | 0.042 | 0.003 | 24.683 | 1.328 | 0.9100 | 0.981 |
| R_paracentral_thickavg | 2894 | -0.020 | 0.027 | -0.072 | 0.032 | 0.007 | 42.450 | 1.738 | 0.4490 | 0.981 |
| R_parsopercularis_thickavg | 2859 | 0.006 | 0.019 | -0.031 | 0.042 | 0.000 | 0.000 | 1.000 | 0.7670 | 0.981 |
| R_parsorbitalis_thickavg | 2882 | -0.013 | 0.029 | -0.070 | 0.044 | 0.010 | 51.293 | 2.053 | 0.6580 | 0.981 |
| R_parstriangularis_thickavg | 2853 | 0.008 | 0.019 | -0.029 | 0.044 | 0.000 | 0.000 | 1.000 | 0.6810 | 0.981 |
| R_pericalcarine_thickavg | 2795 | 0.019 | 0.028 | -0.035 | 0.074 | 0.008 | 45.088 | 1.821 | 0.4870 | 0.981 |
| R_postcentral_thickavg | 2852 | -0.017 | 0.019 | -0.053 | 0.020 | 0.000 | 0.000 | 1.000 | 0.3760 | 0.962 |
| R_posteriorcingulate_thickavg | 2900 | 0.013 | 0.027 | -0.040 | 0.065 | 0.007 | 43.111 | 1.758 | 0.6360 | 0.981 |
| R_precentral_thickavg | 2859 | -0.038 | 0.027 | -0.092 | 0.016 | 0.008 | 45.034 | 1.819 | 0.1690 | 0.910 |
| R_precuneus_thickavg | 2886 | -0.020 | 0.030 | -0.079 | 0.039 | 0.012 | 54.430 | 2.194 | 0.5040 | 0.981 |
| R_rostralanteriorcingulate_thickavg | 2853 | 0.025 | 0.021 | -0.017 | 0.067 | 0.002 | 15.669 | 1.186 | 0.2420 | 0.910 |
| R_rostralmiddlefrontal_thickavg | 2867 | -0.008 | 0.024 | -0.054 | 0.039 | 0.004 | 27.734 | 1.384 | 0.7460 | 0.981 |
| R_superiorfrontal_thickavg | 2881 | 0.013 | 0.021 | -0.029 | 0.055 | 0.002 | 15.955 | 1.190 | 0.5550 | 0.981 |
| R_superiorparietal_thickavg | 2866 | -0.016 | 0.028 | -0.071 | 0.040 | 0.009 | 48.525 | 1.943 | 0.5780 | 0.981 |
| R_superiortemporal_thickavg | 2802 | 0.024 | 0.021 | -0.017 | 0.065 | 0.001 | 11.363 | 1.128 | 0.2560 | 0.910 |
| R_supramarginal_thickavg | 2794 | 0.003 | 0.025 | -0.046 | 0.051 | 0.005 | 32.058 | 1.472 | 0.9150 | 0.981 |
| R_frontalpole_thickavg | 2894 | 0.050 | 0.018 | 0.014 | 0.086 | 0.000 | 0.000 | 1.000 | 0.0070 | 0.145 |
| R_temporalpole_thickavg | 2820 | 0.022 | 0.022 | -0.021 | 0.065 | 0.002 | 17.066 | 1.206 | 0.3100 | 0.910 |
| R_transversetemporal_thickavg | 2905 | 0.012 | 0.024 | -0.034 | 0.058 | 0.004 | 27.716 | 1.383 | 0.6070 | 0.981 |

| Region | N | Effect Size (r) | SE | Lower CI | Upper CI | $tau^2$ | $I^2$ | $H^2$ | pvalue | FDRp |
|---|---|---|---|---|---|---|---|---|---|---|
| R_insula_thickavg | 2797 | -0.013 | 0.023 | -0.058 | 0.031 | 0.003 | 20.723 | 1.261 | 0.5540 | 0.981 |
| LThickness | 2907 | -0.016 | 0.019 | -0.053 | 0.020 | 0.000 | 0.000 | 1.000 | 0.3830 | 0.962 |
| RThickness | 2907 | 0.010 | 0.019 | -0.026 | 0.046 | 0.000 | 0.000 | 1.000 | 0.5890 | 0.981 |

Table S7: Cortical Thickness Continuous Model - No Thickness Covariate

| Region | N | Effect Size (r) | SE | Lower CI | Upper CI | $tau^2$ | $I^2$ | $H^2$ | pvalue | FDRp |
|---|---|---|---|---|---|---|---|---|---|---|
| L_bankssts_thickavg | 2746 | 0.016 | 0.029 | -0.040 | 0.073 | 0.010 | 48.214 | 1.931 | 0.567 | 0.969 |
| L_caudalanteriorcingulate_thickavg | 2884 | 0.017 | 0.019 | -0.019 | 0.054 | 0.000 | 0.000 | 1.000 | 0.356 | 0.969 |
| L_caudalmiddlefrontal_thickavg | 2886 | -0.004 | 0.031 | -0.064 | 0.057 | 0.013 | 57.288 | 2.341 | 0.907 | 0.969 |
| L_cuneus_thickavg | 2777 | -0.004 | 0.026 | -0.055 | 0.047 | 0.006 | 36.154 | 1.566 | 0.878 | 0.969 |
| L_entorhinal_thickavg | 2755 | -0.022 | 0.019 | -0.059 | 0.016 | 0.000 | 0.000 | 1.000 | 0.257 | 0.969 |
| L_fusiform_thickavg | 2870 | -0.028 | 0.023 | -0.073 | 0.018 | 0.004 | 26.235 | 1.356 | 0.237 | 0.969 |
| L_inferiorparietal_thickavg | 2828 | -0.012 | 0.019 | -0.049 | 0.025 | 0.000 | 0.000 | 1.000 | 0.524 | 0.969 |
| L_inferiortemporal_thickavg | 2843 | -0.010 | 0.019 | -0.047 | 0.026 | 0.000 | 0.000 | 1.000 | 0.577 | 0.969 |
| L_isthmuscingulate_thickavg | 2898 | -0.022 | 0.027 | -0.075 | 0.032 | 0.008 | 45.032 | 1.819 | 0.422 | 0.969 |
| L_lateraloccipital_thickavg | 2866 | 0.030 | 0.025 | -0.019 | 0.079 | 0.005 | 35.385 | 1.548 | 0.236 | 0.969 |
| L_lateralorbitofrontal_thickavg | 2873 | 0.004 | 0.027 | -0.048 | 0.056 | 0.007 | 41.370 | 1.706 | 0.878 | 0.969 |
| L_lingual_thickavg | 2842 | -0.018 | 0.019 | -0.055 | 0.020 | 0.000 | 2.970 | 1.031 | 0.357 | 0.969 |
| L_medialorbitofrontal_thickavg | 2862 | 0.051 | 0.022 | 0.008 | 0.095 | 0.002 | 19.860 | 1.248 | 0.021 | 0.246 |
| L_middletemporal_thickavg | 2756 | -0.013 | 0.019 | -0.050 | 0.024 | 0.000 | 0.000 | 1.000 | 0.493 | 0.969 |
| L_parahippocampal_thickavg | 2888 | 0.012 | 0.028 | -0.043 | 0.067 | 0.009 | 48.631 | 1.947 | 0.669 | 0.969 |
| L_paracentral_thickavg | 2897 | -0.040 | 0.021 | -0.082 | 0.001 | 0.002 | 15.386 | 1.182 | 0.057 | 0.514 |
| L_parsopercularis_thickavg | 2875 | -0.005 | 0.024 | -0.052 | 0.042 | 0.004 | 30.249 | 1.434 | 0.838 | 0.969 |
| L_parsorbitalis_thickavg | 2885 | 0.003 | 0.019 | -0.033 | 0.040 | 0.000 | 0.000 | 1.000 | 0.868 | 0.969 |
| L_parstriangularis_thickavg | 2873 | -0.014 | 0.034 | -0.080 | 0.052 | 0.018 | 64.460 | 2.814 | 0.685 | 0.969 |
| L_pericalcarine_thickavg | 2811 | -0.020 | 0.025 | -0.068 | 0.029 | 0.005 | 30.743 | 1.444 | 0.426 | 0.969 |
| L_postcentral_thickavg | 2838 | -0.017 | 0.027 | -0.070 | 0.036 | 0.008 | 43.400 | 1.767 | 0.521 | 0.969 |
| L_posteriorcingulate_thickavg | 2896 | 0.023 | 0.019 | -0.013 | 0.060 | 0.000 | 0.000 | 1.000 | 0.208 | 0.969 |
| L_precentral_thickavg | 2855 | -0.022 | 0.022 | -0.064 | 0.020 | 0.002 | 16.485 | 1.197 | 0.307 | 0.969 |
| L_precuneus_thickavg | 2887 | -0.003 | 0.023 | -0.047 | 0.042 | 0.003 | 22.911 | 1.297 | 0.899 | 0.969 |
| L_rostralanteriorcingulate_thickavg | 2875 | 0.036 | 0.019 | -0.001 | 0.072 | 0.000 | 0.000 | 1.000 | 0.056 | 0.514 |
| L_rostralmiddlefrontal_thickavg | 2886 | -0.012 | 0.024 | -0.059 | 0.035 | 0.004 | 29.607 | 1.421 | 0.620 | 0.969 |
| L_superiorfrontal_thickavg | 2879 | -0.010 | 0.019 | -0.046 | 0.027 | 0.000 | 0.000 | 1.000 | 0.597 | 0.969 |

| Region | N | Effect Size (r) | SE | Lower CI | Upper CI | $tau^2$ | $I^2$ | $H^2$ | pvalue | FDRp |
|---|---|---|---|---|---|---|---|---|---|---|
| L_superiorparietal_thickavg | 2851 | -0.023 | 0.020 | -0.063 | 0.016 | 0.001 | 8.610 | 1.094 | 0.252 | 0.969 |
| L_superiortemporal_thickavg | 2733 | -0.002 | 0.028 | -0.056 | 0.052 | 0.008 | 43.695 | 1.776 | 0.945 | 0.991 |
| L_supramarginal_thickavg | 2775 | -0.027 | 0.019 | -0.064 | 0.010 | 0.000 | 0.000 | 1.000 | 0.149 | 0.969 |
| L_frontalpole_thickavg | 2902 | 0.044 | 0.018 | 0.008 | 0.080 | 0.000 | 0.000 | 1.000 | 0.017 | 0.233 |
| L_temporalpole_thickavg | 2864 | 0.020 | 0.019 | -0.017 | 0.056 | 0.000 | 0.000 | 1.000 | 0.290 | 0.969 |
| L_transversetemporal_thickavg | 2903 | 0.035 | 0.021 | -0.007 | 0.077 | 0.002 | 16.767 | 1.201 | 0.101 | 0.779 |
| L_insula_thickavg | 2829 | 0.000 | 0.025 | -0.048 | 0.049 | 0.005 | 32.748 | 1.487 | 0.997 | 0.997 |
| R_bankssts_thickavg | 2840 | 0.026 | 0.027 | -0.026 | 0.079 | 0.008 | 43.210 | 1.761 | 0.327 | 0.969 |
| R_caudalanteriorcingulate_thickavg | 2897 | 0.024 | 0.029 | -0.033 | 0.081 | 0.011 | 51.777 | 2.074 | 0.412 | 0.969 |
| R_caudalmiddlefrontal_thickavg | 2884 | -0.002 | 0.022 | -0.046 | 0.042 | 0.003 | 21.706 | 1.277 | 0.927 | 0.981 |
| R_cuneus_thickavg | 2793 | -0.012 | 0.024 | -0.059 | 0.035 | 0.004 | 28.547 | 1.400 | 0.618 | 0.969 |
| R_entorhinal_thickavg | 2695 | 0.003 | 0.021 | -0.038 | 0.044 | 0.001 | 9.055 | 1.100 | 0.893 | 0.969 |
| R_fusiform_thickavg | 2877 | 0.000 | 0.024 | -0.046 | 0.046 | 0.004 | 27.405 | 1.378 | 0.991 | 0.997 |
| R_inferiorparietal_thickavg | 2828 | 0.008 | 0.028 | -0.047 | 0.062 | 0.008 | 45.382 | 1.831 | 0.784 | 0.969 |
| R_inferiortemporal_thickavg | 2855 | -0.020 | 0.029 | -0.077 | 0.037 | 0.010 | 51.045 | 2.043 | 0.495 | 0.969 |
| R_isthmuscingulate_thickavg | 2890 | -0.047 | 0.019 | -0.084 | -0.011 | 0.000 | 0.000 | 1.000 | 0.011 | 0.164 |
| R_lateraloccipital_thickavg | 2868 | 0.006 | 0.028 | -0.048 | 0.060 | 0.008 | 45.453 | 1.833 | 0.830 | 0.969 |
| R_lateralorbitofrontal_thickavg | 2853 | 0.007 | 0.032 | -0.056 | 0.069 | 0.014 | 58.990 | 2.438 | 0.835 | 0.969 |
| R_lingual_thickavg | 2843 | -0.023 | 0.027 | -0.075 | 0.029 | 0.007 | 41.524 | 1.710 | 0.386 | 0.969 |
| R_medialorbitofrontal_thickavg | 2847 | 0.061 | 0.019 | 0.025 | 0.097 | 0.000 | 0.000 | 1.000 | 0.001 | 0.037 |
| R_middletemporal_thickavg | 2832 | 0.023 | 0.029 | -0.033 | 0.078 | 0.010 | 48.635 | 1.947 | 0.430 | 0.969 |
| R_parahippocampal_thickavg | 2897 | -0.005 | 0.024 | -0.052 | 0.043 | 0.005 | 32.269 | 1.476 | 0.848 | 0.969 |
| R_paracentral_thickavg | 2894 | -0.019 | 0.029 | -0.076 | 0.038 | 0.010 | 51.473 | 2.061 | 0.515 | 0.969 |
| R_parsopercularis_thickavg | 2859 | 0.004 | 0.019 | -0.032 | 0.041 | 0.000 | 0.000 | 1.000 | 0.813 | 0.969 |
| R_parsorbitalis_thickavg | 2882 | -0.010 | 0.030 | -0.068 | 0.049 | 0.012 | 53.737 | 2.162 | 0.745 | 0.969 |
| R_parstriangularis_thickavg | 2853 | 0.001 | 0.019 | -0.036 | 0.038 | 0.000 | 0.000 | 1.000 | 0.958 | 0.995 |
| R_pericalcarine_thickavg | 2795 | 0.019 | 0.032 | -0.044 | 0.082 | 0.015 | 59.607 | 2.476 | 0.558 | 0.969 |
| R_postcentral_thickavg | 2852 | -0.024 | 0.024 | -0.071 | 0.024 | 0.004 | 29.856 | 1.426 | 0.326 | 0.969 |
| R_posteriorcingulate_thickavg | 2900 | 0.013 | 0.018 | -0.024 | 0.049 | 0.000 | 0.000 | 1.000 | 0.493 | 0.969 |
| R_precentral_thickavg | 2859 | -0.041 | 0.028 | -0.096 | 0.015 | 0.010 | 49.213 | 1.969 | 0.153 | 0.969 |
| R_precuneus_thickavg | 2886 | -0.014 | 0.031 | -0.074 | 0.046 | 0.013 | 56.638 | 2.306 | 0.653 | 0.969 |
| R_rostralanteriorcingulate_thickavg | 2853 | 0.030 | 0.024 | -0.017 | 0.076 | 0.004 | 27.177 | 1.373 | 0.209 | 0.969 |
| R_rostralmiddlefrontal_thickavg | 2867 | -0.002 | 0.019 | -0.039 | 0.034 | 0.000 | 0.000 | 1.000 | 0.903 | 0.969 |
| R_superiorfrontal_thickavg | 2881 | -0.001 | 0.024 | -0.047 | 0.045 | 0.004 | 27.735 | 1.384 | 0.974 | 0.997 |

| Region | N | Effect Size (r) | SE | Lower CI | Upper CI | $tau^2$ | $I^2$ | $H^2$ | pvalue | FDRp |
|---|---|---|---|---|---|---|---|---|---|---|
| R_superiorparietal_thickavg | 2866 | -0.016 | 0.028 | -0.071 | 0.039 | 0.009 | 48.389 | 1.938 | 0.571 | 0.969 |
| R_superiortemporal_thickavg | 2802 | 0.011 | 0.023 | -0.034 | 0.056 | 0.003 | 22.902 | 1.297 | 0.644 | 0.969 |
| R_supramarginal_thickavg | 2794 | 0.000 | 0.019 | -0.037 | 0.037 | 0.000 | 0.000 | 1.000 | 0.990 | 0.997 |
| R_frontalpole_thickavg | 2894 | 0.049 | 0.018 | 0.012 | 0.085 | 0.000 | 0.000 | 1.000 | 0.008 | 0.152 |
| R_temporalpole_thickavg | 2820 | 0.017 | 0.024 | -0.029 | 0.063 | 0.004 | 25.975 | 1.351 | 0.466 | 0.969 |
| R_transversetemporal_thickavg | 2905 | 0.011 | 0.023 | -0.035 | 0.057 | 0.004 | 27.639 | 1.382 | 0.637 | 0.969 |
| R_insula_thickavg | 2797 | -0.014 | 0.026 | -0.064 | 0.036 | 0.006 | 35.994 | 1.562 | 0.582 | 0.969 |
| LThickness | 2907 | -0.014 | 0.024 | -0.061 | 0.033 | 0.004 | 30.355 | 1.436 | 0.550 | 0.969 |
| RThickness | 2907 | -0.010 | 0.027 | -0.063 | 0.042 | 0.008 | 43.677 | 1.775 | 0.699 | 0.969 |

### Cortical Surface Models

##### Note - Measures of heterogeneity

$Tau^2$: Represents the absolute value of the true variance of the effect sizes, between-study variance

The $I^2$ statistic: Represents the proportion (in percentage) of total variation in the estimates that can be attributed to heterogeneity among the effects rather than sampling variability. The $I^2$ may be interpreted according to the following ranges $I^2$ = 0-40%: low heterogeneity, $I^2$ = 30-60%: moderate heterogeneity, $I^2$ = 50-90%: substantial heterogeneity, $I^2$ = 75-100%: considerable heterogeneity

The $H^2$ statistic: Represents the ratio of the total amount of variability in the effect size estimates to the amount of sampling variability

##### Table S8: Cortical Surface Area Continuous Model

| Region | N | Effect Size (r) | SE | Lower CI | Upper CI | $tau^2$ | $I^2$ | $H^2$ | pvalue | FDRp |
|---|---|---|---|---|---|---|---|---|---|---|
| L_bankssts_surfavg | 2707 | 0.009 | 0.021 | -0.032 | 0.050 | 0.001 | 8.943 | 1.098 | 0.672 | 0.905 |
| L_caudalanteriorcingulate_surfavg | 2845 | -0.006 | 0.021 | -0.047 | 0.035 | 0.001 | 12.394 | 1.141 | 0.780 | 0.980 |
| L_caudalmiddlefrontal_surfavg | 2848 | -0.006 | 0.019 | -0.044 | 0.032 | 0.000 | 4.217 | 1.044 | 0.761 | 0.967 |
| L_cuneus_surfavg | 2738 | -0.013 | 0.019 | -0.050 | 0.025 | 0.000 | 0.000 | 1.000 | 0.509 | 0.859 |
| L_entorhinal_surfavg | 2716 | -0.027 | 0.022 | -0.070 | 0.016 | 0.002 | 15.371 | 1.182 | 0.217 | 0.856 |
| L_fusiform_surfavg | 2831 | -0.043 | 0.021 | -0.084 | -0.002 | 0.001 | 11.497 | 1.130 | 0.038 | 0.827 |
| L_inferiorparietal_surfavg | 2790 | -0.013 | 0.022 | -0.056 | 0.030 | 0.002 | 17.472 | 1.212 | 0.552 | 0.882 |
| L_inferiortemporal_surfavg | 2804 | -0.002 | 0.024 | -0.049 | 0.044 | 0.004 | 27.412 | 1.378 | 0.922 | 0.981 |
| L_isthmuscingulate_surfavg | 2859 | 0.012 | 0.019 | -0.025 | 0.048 | 0.000 | 0.000 | 1.000 | 0.529 | 0.879 |
| L_lateraloccipital_surfavg | 2827 | -0.027 | 0.021 | -0.067 | 0.014 | 0.001 | 10.670 | 1.119 | 0.195 | 0.856 |
| L_lateralorbitofrontal_surfavg | 2834 | 0.007 | 0.019 | -0.030 | 0.043 | 0.000 | 0.000 | 1.000 | 0.716 | 0.932 |
| L_lingual_surfavg | 2803 | 0.001 | 0.025 | -0.048 | 0.050 | 0.005 | 33.945 | 1.514 | 0.964 | 0.981 |
| L_medialorbitofrontal_surfavg | 2823 | -0.037 | 0.034 | -0.103 | 0.029 | 0.018 | 64.449 | 2.813 | 0.274 | 0.856 |

| Region | N | Effect Size (r) | SE | Lower CI | Upper CI | $tau^2$ | $I^2$ | $H^2$ | pvalue | FDRp |
|---|---|---|---|---|---|---|---|---|---|---|
| L_middletemporal_surfavg | 2717 | 0.027 | 0.019 | -0.010 | 0.065 | 0.000 | 0.911 | 1.009 | 0.156 | 0.856 |
| L_parahippocampal_surfavg | 2848 | -0.056 | 0.020 | -0.096 | -0.016 | 0.001 | 9.139 | 1.101 | 0.006 | 0.205 |
| L_paracentral_surfavg | 2858 | 0.019 | 0.028 | -0.036 | 0.075 | 0.009 | 48.056 | 1.925 | 0.494 | 0.856 |
| L_parsopercularis_surfavg | 2836 | 0.013 | 0.019 | -0.024 | 0.050 | 0.000 | 0.000 | 1.000 | 0.486 | 0.856 |
| L_parsorbitalis_surfavg | 2846 | 0.012 | 0.023 | -0.033 | 0.057 | 0.003 | 23.378 | 1.305 | 0.605 | 0.905 |
| L_parstriangularis_surfavg | 2834 | -0.022 | 0.022 | -0.066 | 0.022 | 0.003 | 19.970 | 1.250 | 0.322 | 0.856 |
| L_pericalcarine_surfavg | 2772 | -0.015 | 0.021 | -0.055 | 0.026 | 0.001 | 9.724 | 1.108 | 0.482 | 0.856 |
| L_postcentral_surfavg | 2800 | 0.039 | 0.025 | -0.010 | 0.087 | 0.005 | 32.277 | 1.477 | 0.120 | 0.856 |
| L_posteriorcingulate_surfavg | 2855 | 0.026 | 0.019 | -0.011 | 0.063 | 0.000 | 0.000 | 1.000 | 0.165 | 0.856 |
| L_precentral_surfavg | 2816 | -0.009 | 0.022 | -0.053 | 0.035 | 0.003 | 19.958 | 1.249 | 0.675 | 0.905 |
| L_precuneus_surfavg | 2848 | 0.002 | 0.035 | -0.066 | 0.071 | 0.020 | 67.255 | 3.054 | 0.948 | 0.981 |
| L_rostralanteriorcingulate_surfavg | 2836 | -0.021 | 0.023 | -0.066 | 0.024 | 0.003 | 23.790 | 1.312 | 0.358 | 0.856 |
| L_rostralmiddlefrontal_surfavg | 2847 | 0.021 | 0.019 | -0.016 | 0.058 | 0.000 | 0.000 | 1.000 | 0.260 | 0.856 |
| L_superiorfrontal_surfavg | 2840 | 0.005 | 0.037 | -0.067 | 0.077 | 0.024 | 70.694 | 3.412 | 0.898 | 0.981 |
| L_superiorparietal_surfavg | 2812 | 0.018 | 0.026 | -0.032 | 0.068 | 0.006 | 35.764 | 1.557 | 0.478 | 0.856 |
| L_superiortemporal_surfavg | 2695 | 0.006 | 0.023 | -0.040 | 0.051 | 0.003 | 21.758 | 1.278 | 0.805 | 0.981 |
| L_supramarginal_surfavg | 2737 | 0.048 | 0.032 | -0.014 | 0.110 | 0.014 | 57.169 | 2.335 | 0.129 | 0.856 |
| L_frontalpole_surfavg | 2863 | -0.011 | 0.022 | -0.053 | 0.032 | 0.002 | 17.213 | 1.208 | 0.625 | 0.905 |
| L_temporalpole_surfavg | 2824 | -0.033 | 0.025 | -0.081 | 0.015 | 0.005 | 31.502 | 1.460 | 0.180 | 0.856 |
| L_transversetemporal_surfavg | 2864 | -0.018 | 0.020 | -0.057 | 0.021 | 0.001 | 8.165 | 1.089 | 0.367 | 0.856 |
| L_insula_surfavg | 2790 | -0.013 | 0.019 | -0.050 | 0.024 | 0.000 | 0.000 | 1.000 | 0.476 | 0.856 |
| R_bankssts_surfavg | 2801 | 0.019 | 0.027 | -0.034 | 0.071 | 0.007 | 41.195 | 1.701 | 0.489 | 0.856 |
| R_caudalanteriorcingulate_surfavg | 2857 | -0.018 | 0.022 | -0.061 | 0.026 | 0.002 | 19.412 | 1.241 | 0.420 | 0.856 |
| R_caudalmiddlefrontal_surfavg | 2845 | -0.026 | 0.019 | -0.062 | 0.011 | 0.000 | 0.000 | 1.000 | 0.169 | 0.856 |
| R_cuneus_surfavg | 2754 | -0.015 | 0.019 | -0.052 | 0.022 | 0.000 | 0.000 | 1.000 | 0.419 | 0.856 |
| R_entorhinal_surfavg | 2654 | -0.036 | 0.025 | -0.085 | 0.013 | 0.005 | 29.647 | 1.421 | 0.152 | 0.856 |
| R_fusiform_surfavg | 2838 | 0.003 | 0.022 | -0.041 | 0.046 | 0.002 | 18.627 | 1.229 | 0.900 | 0.981 |
| R_inferiorparietal_surfavg | 2791 | 0.010 | 0.021 | -0.032 | 0.052 | 0.002 | 14.208 | 1.166 | 0.648 | 0.905 |
| R_inferiortemporal_surfavg | 2816 | -0.002 | 0.025 | -0.051 | 0.048 | 0.006 | 35.360 | 1.547 | 0.952 | 0.981 |
| R_isthmuscingulate_surfavg | 2850 | 0.027 | 0.024 | -0.021 | 0.075 | 0.004 | 30.620 | 1.441 | 0.268 | 0.856 |
| R_lateraloccipital_surfavg | 2829 | -0.003 | 0.029 | -0.059 | 0.053 | 0.010 | 49.222 | 1.969 | 0.921 | 0.981 |
| R_lateralorbitofrontal_surfavg | 2814 | -0.009 | 0.023 | -0.055 | 0.036 | 0.003 | 24.657 | 1.327 | 0.687 | 0.905 |
| R_lingual_surfavg | 2804 | -0.008 | 0.019 | -0.045 | 0.029 | 0.000 | 0.000 | 1.000 | 0.678 | 0.905 |
| R_medialorbitofrontal_surfavg | 2808 | -0.062 | 0.032 | -0.124 | 0.000 | 0.014 | 58.977 | 2.438 | 0.051 | 0.856 |

| Region | N | Effect Size (r) | SE | Lower CI | Upper CI | $tau^2$ | $I^2$ | $H^2$ | pvalue | FDRp |
|---|---|---|---|---|---|---|---|---|---|---|
| R_middletemporal_surfavg | 2795 | 0.000 | 0.019 | -0.037 | 0.036 | 0.000 | 0.000 | 1.000 | 0.981 | 0.981 |
| R_parahippocampal_surfavg | 2858 | -0.001 | 0.022 | -0.044 | 0.041 | 0.002 | 17.058 | 1.206 | 0.952 | 0.981 |
| R_paracentral_surfavg | 2855 | -0.005 | 0.024 | -0.052 | 0.043 | 0.004 | 30.618 | 1.441 | 0.842 | 0.981 |
| R_parsopercularis_surfavg | 2820 | 0.017 | 0.019 | -0.020 | 0.053 | 0.000 | 0.000 | 1.000 | 0.376 | 0.856 |
| R_parsorbitalis_surfavg | 2843 | -0.028 | 0.028 | -0.083 | 0.027 | 0.009 | 47.357 | 1.900 | 0.321 | 0.856 |
| R_parstriangularis_surfavg | 2814 | -0.002 | 0.019 | -0.039 | 0.035 | 0.000 | 0.000 | 1.000 | 0.918 | 0.981 |
| R_pericalcarine_surfavg | 2756 | -0.033 | 0.019 | -0.071 | 0.004 | 0.000 | 0.000 | 1.000 | 0.078 | 0.856 |
| R_postcentral_surfavg | 2814 | 0.035 | 0.022 | -0.009 | 0.078 | 0.002 | 19.102 | 1.236 | 0.120 | 0.856 |
| R_posteriorcingulate_surfavg | 2859 | -0.001 | 0.025 | -0.049 | 0.048 | 0.005 | 32.457 | 1.481 | 0.976 | 0.981 |
| R_precentral_surfavg | 2820 | -0.001 | 0.019 | -0.037 | 0.036 | 0.000 | 0.000 | 1.000 | 0.977 | 0.981 |
| R_precuneus_surfavg | 2847 | 0.040 | 0.032 | -0.023 | 0.104 | 0.016 | 61.102 | 2.571 | 0.214 | 0.856 |
| R_rostralanteriorcingulate_surfavg | 2814 | -0.009 | 0.019 | -0.046 | 0.027 | 0.000 | 0.000 | 1.000 | 0.616 | 0.905 |
| R_rostralmiddlefrontal_surfavg | 2828 | 0.023 | 0.021 | -0.018 | 0.064 | 0.001 | 11.952 | 1.136 | 0.278 | 0.856 |
| R_superiorfrontal_surfavg | 2842 | -0.005 | 0.022 | -0.047 | 0.038 | 0.002 | 16.903 | 1.203 | 0.828 | 0.981 |
| R_superiorparietal_surfavg | 2827 | 0.025 | 0.024 | -0.023 | 0.072 | 0.005 | 30.749 | 1.444 | 0.317 | 0.856 |
| R_superiortemporal_surfavg | 2763 | 0.029 | 0.030 | -0.030 | 0.087 | 0.011 | 52.633 | 2.111 | 0.338 | 0.856 |
| R_supramarginal_surfavg | 2756 | -0.009 | 0.029 | -0.066 | 0.048 | 0.010 | 49.241 | 1.970 | 0.749 | 0.963 |
| R_frontalpole_surfavg | 2854 | 0.021 | 0.021 | -0.020 | 0.063 | 0.002 | 14.926 | 1.175 | 0.314 | 0.856 |
| R_temporalpole_surfavg | 2781 | -0.014 | 0.019 | -0.051 | 0.023 | 0.000 | 0.000 | 1.000 | 0.465 | 0.856 |
| R_transversetemporal_surfavg | 2866 | -0.011 | 0.019 | -0.047 | 0.025 | 0.000 | 0.000 | 1.000 | 0.551 | 0.882 |
| R_insula_surfavg | 2758 | -0.003 | 0.021 | -0.043 | 0.038 | 0.001 | 9.129 | 1.100 | 0.892 | 0.981 |
| LSurfArea | 2868 | -0.014 | 0.020 | -0.052 | 0.025 | 0.001 | 5.627 | 1.060 | 0.487 | 0.856 |
| RSurfArea | 2868 | 0.014 | 0.020 | -0.025 | 0.052 | 0.001 | 5.627 | 1.060 | 0.487 | 0.856 |

Table S9: Cortical Surface Area Continuous Model - No Surface Area Covariate

| Region | N | Effect Size (r) | SE | Lower CI | Upper CI | $tau^2$ | $I^2$ | $H^2$ | pvalue | FDRp |
|---|---|---|---|---|---|---|---|---|---|---|
| L_bankssts_surfavg | 2682 | 0.009 | 0.019 | -0.028 | 0.047 | 0.000 | 0.000 | 1.000 | 0.627 | 0.916 |
| L_caudalanteriorcingulate_surfavg | 2812 | -0.005 | 0.022 | -0.048 | 0.039 | 0.002 | 19.198 | 1.238 | 0.834 | 0.936 |
| L_caudalmiddlefrontal_surfavg | 2815 | -0.009 | 0.021 | -0.050 | 0.033 | 0.002 | 12.851 | 1.147 | 0.685 | 0.924 |
| L_cuneus_surfavg | 2717 | -0.018 | 0.020 | -0.058 | 0.021 | 0.001 | 6.306 | 1.067 | 0.364 | 0.916 |
| L_entorhinal_surfavg | 2683 | -0.024 | 0.019 | -0.063 | 0.014 | 0.000 | 1.631 | 1.017 | 0.209 | 0.916 |
| L_fusiform_surfavg | 2798 | -0.036 | 0.019 | -0.072 | 0.001 | 0.000 | 0.000 | 1.000 | 0.059 | 0.916 |
| L_inferiorparietal_surfavg | 2757 | -0.007 | 0.019 | -0.044 | 0.030 | 0.000 | 0.000 | 1.000 | 0.712 | 0.931 |

| Region | N | Effect Size (r) | SE | Lower CI | Upper CI | $tau^2$ | $I^2$ | $H^2$ | pvalue | FDRp |
|---|---|---|---|---|---|---|---|---|---|---|
| L_inferiortemporal_surfavg | 2771 | -0.010 | 0.019 | -0.047 | 0.027 | 0.000 | 0.000 | 1.000 | 0.590 | 0.916 |
| L_isthmuscingulate_surfavg | 2826 | 0.008 | 0.019 | -0.028 | 0.045 | 0.000 | 0.000 | 1.000 | 0.659 | 0.924 |
| L_lateraloccipital_surfavg | 2794 | -0.033 | 0.019 | -0.070 | 0.003 | 0.000 | 0.000 | 1.000 | 0.075 | 0.916 |
| L_lateralorbitofrontal_surfavg | 2801 | -0.005 | 0.019 | -0.042 | 0.032 | 0.000 | 0.000 | 1.000 | 0.783 | 0.936 |
| L_lingual_surfavg | 2778 | 0.002 | 0.025 | -0.048 | 0.052 | 0.005 | 34.053 | 1.516 | 0.932 | 0.949 |
| L_medialorbitofrontal_surfavg | 2790 | -0.027 | 0.032 | -0.089 | 0.035 | 0.014 | 58.706 | 2.422 | 0.398 | 0.916 |
| L_middletemporal_surfavg | 2691 | 0.013 | 0.019 | -0.025 | 0.050 | 0.000 | 0.000 | 1.000 | 0.514 | 0.916 |
| L_parahippocampal_surfavg | 2815 | -0.051 | 0.019 | -0.087 | -0.014 | 0.000 | 0.000 | 1.000 | 0.007 | 0.253 |
| L_paracentral_surfavg | 2825 | 0.021 | 0.028 | -0.035 | 0.076 | 0.009 | 48.043 | 1.925 | 0.465 | 0.916 |
| L_parsopercularis_surfavg | 2803 | 0.010 | 0.019 | -0.026 | 0.047 | 0.000 | 0.000 | 1.000 | 0.580 | 0.916 |
| L_parsorbitalis_surfavg | 2813 | 0.012 | 0.019 | -0.025 | 0.049 | 0.000 | 0.000 | 1.000 | 0.521 | 0.916 |
| L_parstriangularis_surfavg | 2801 | -0.018 | 0.019 | -0.055 | 0.019 | 0.000 | 0.000 | 1.000 | 0.344 | 0.916 |
| L_pericalcarine_surfavg | 2751 | -0.018 | 0.024 | -0.065 | 0.028 | 0.003 | 24.680 | 1.328 | 0.439 | 0.916 |
| L_postcentral_surfavg | 2771 | 0.015 | 0.019 | -0.022 | 0.052 | 0.000 | 0.000 | 1.000 | 0.431 | 0.916 |
| L_posteriorcingulate_surfavg | 2822 | 0.013 | 0.019 | -0.024 | 0.049 | 0.000 | 0.000 | 1.000 | 0.504 | 0.916 |
| L_precentral_surfavg | 2787 | -0.017 | 0.019 | -0.054 | 0.019 | 0.000 | 0.000 | 1.000 | 0.355 | 0.916 |
| L_precuneus_surfavg | 2815 | 0.008 | 0.034 | -0.059 | 0.074 | 0.018 | 64.406 | 2.809 | 0.823 | 0.936 |
| L_rostralanteriorcingulate_surfavg | 2803 | -0.020 | 0.020 | -0.060 | 0.020 | 0.001 | 8.361 | 1.091 | 0.334 | 0.916 |
| L_rostralmiddlefrontal_surfavg | 2814 | 0.009 | 0.019 | -0.028 | 0.045 | 0.000 | 0.000 | 1.000 | 0.648 | 0.924 |
| L_superiorfrontal_surfavg | 2807 | 0.003 | 0.029 | -0.053 | 0.059 | 0.009 | 47.954 | 1.921 | 0.920 | 0.949 |
| L_superiorparietal_surfavg | 2779 | 0.023 | 0.027 | -0.030 | 0.076 | 0.007 | 41.086 | 1.697 | 0.398 | 0.916 |
| L_superiortemporal_surfavg | 2665 | 0.008 | 0.023 | -0.037 | 0.053 | 0.003 | 19.092 | 1.236 | 0.736 | 0.935 |
| L_supramarginal_surfavg | 2705 | 0.033 | 0.026 | -0.017 | 0.083 | 0.005 | 33.790 | 1.510 | 0.194 | 0.916 |
| L_frontalpole_surfavg | 2830 | -0.006 | 0.024 | -0.054 | 0.042 | 0.005 | 31.018 | 1.450 | 0.800 | 0.936 |
| L_temporalpole_surfavg | 2791 | -0.024 | 0.023 | -0.070 | 0.021 | 0.003 | 24.406 | 1.323 | 0.298 | 0.916 |
| L_transversetemporal_surfavg | 2831 | -0.016 | 0.019 | -0.053 | 0.021 | 0.000 | 0.000 | 1.000 | 0.391 | 0.916 |
| L_insula_surfavg | 2757 | -0.021 | 0.024 | -0.068 | 0.025 | 0.004 | 26.053 | 1.352 | 0.368 | 0.916 |
| R_bankssts_surfavg | 2768 | 0.019 | 0.027 | -0.035 | 0.073 | 0.008 | 43.014 | 1.755 | 0.486 | 0.916 |
| R_caudalanteriorcingulate_surfavg | 2824 | -0.017 | 0.019 | -0.054 | 0.019 | 0.000 | 0.089 | 1.001 | 0.353 | 0.916 |
| R_caudalmiddlefrontal_surfavg | 2812 | -0.027 | 0.019 | -0.063 | 0.010 | 0.000 | 0.000 | 1.000 | 0.156 | 0.916 |
| R_cuneus_surfavg | 2731 | -0.019 | 0.023 | -0.065 | 0.027 | 0.003 | 23.123 | 1.301 | 0.413 | 0.916 |
| R_entorhinal_surfavg | 2621 | -0.042 | 0.029 | -0.099 | 0.016 | 0.010 | 47.935 | 1.921 | 0.157 | 0.916 |
| R_fusiform_surfavg | 2805 | -0.005 | 0.019 | -0.042 | 0.032 | 0.000 | 0.000 | 1.000 | 0.783 | 0.936 |
| R_inferiorparietal_surfavg | 2758 | 0.000 | 0.019 | -0.038 | 0.037 | 0.000 | 0.000 | 1.000 | 0.981 | 0.981 |

| Region | N | Effect Size (r) | SE | Lower CI | Upper CI | $tau^2$ | $I^2$ | $H^2$ | pvalue | FDRp |
|---|---|---|---|---|---|---|---|---|---|---|
| R_inferiortemporal_surfavg | 2783 | -0.008 | 0.023 | -0.052 | 0.036 | 0.003 | 20.114 | 1.252 | 0.726 | 0.934 |
| R_isthmuscingulate_surfavg | 2817 | 0.030 | 0.029 | -0.026 | 0.087 | 0.010 | 50.300 | 2.012 | 0.294 | 0.916 |
| R_lateraloccipital_surfavg | 2796 | -0.019 | 0.019 | -0.056 | 0.018 | 0.000 | 0.000 | 1.000 | 0.322 | 0.916 |
| R_lateralorbitofrontal_surfavg | 2781 | -0.010 | 0.019 | -0.047 | 0.027 | 0.000 | 0.000 | 1.000 | 0.587 | 0.916 |
| R_lingual_surfavg | 2781 | -0.016 | 0.019 | -0.053 | 0.021 | 0.000 | 0.000 | 1.000 | 0.401 | 0.916 |
| R_medialorbitofrontal_surfavg | 2775 | -0.039 | 0.019 | -0.076 | -0.002 | 0.000 | 0.000 | 1.000 | 0.038 | 0.916 |
| R_middletemporal_surfavg | 2762 | -0.015 | 0.019 | -0.053 | 0.022 | 0.000 | 0.000 | 1.000 | 0.415 | 0.916 |
| R_parahippocampal_surfavg | 2825 | -0.005 | 0.021 | -0.046 | 0.036 | 0.001 | 10.837 | 1.122 | 0.806 | 0.936 |
| R_paracentral_surfavg | 2822 | -0.006 | 0.027 | -0.059 | 0.047 | 0.008 | 42.734 | 1.746 | 0.830 | 0.936 |
| R_parsopercularis_surfavg | 2787 | 0.012 | 0.019 | -0.025 | 0.049 | 0.000 | 0.000 | 1.000 | 0.531 | 0.916 |
| R_parsorbitalis_surfavg | 2810 | -0.022 | 0.024 | -0.068 | 0.024 | 0.004 | 25.931 | 1.350 | 0.348 | 0.916 |
| R_parstriangularis_surfavg | 2781 | -0.004 | 0.019 | -0.041 | 0.033 | 0.000 | 0.000 | 1.000 | 0.842 | 0.936 |
| R_pericalcarine_surfavg | 2735 | -0.034 | 0.019 | -0.072 | 0.003 | 0.000 | 0.000 | 1.000 | 0.072 | 0.916 |
| R_postcentral_surfavg | 2785 | 0.028 | 0.022 | -0.016 | 0.073 | 0.003 | 19.763 | 1.246 | 0.205 | 0.916 |
| R_posteriorcingulate_surfavg | 2826 | -0.005 | 0.019 | -0.043 | 0.032 | 0.000 | 2.394 | 1.025 | 0.776 | 0.936 |
| R_precentral_surfavg | 2791 | -0.008 | 0.019 | -0.045 | 0.029 | 0.000 | 0.000 | 1.000 | 0.684 | 0.924 |
| R_precuneus_surfavg | 2814 | 0.034 | 0.031 | -0.026 | 0.094 | 0.012 | 55.228 | 2.234 | 0.270 | 0.916 |
| R_rostralanteriorcingulate_surfavg | 2781 | -0.011 | 0.020 | -0.051 | 0.029 | 0.001 | 8.245 | 1.090 | 0.592 | 0.916 |
| R_rostralmiddlefrontal_surfavg | 2795 | 0.001 | 0.019 | -0.036 | 0.038 | 0.000 | 0.000 | 1.000 | 0.967 | 0.976 |
| R_superiorfrontal_surfavg | 2809 | -0.006 | 0.024 | -0.052 | 0.040 | 0.004 | 25.644 | 1.345 | 0.805 | 0.936 |
| R_superiorparietal_surfavg | 2794 | 0.021 | 0.027 | -0.031 | 0.074 | 0.007 | 40.321 | 1.676 | 0.425 | 0.916 |
| R_superiortemporal_surfavg | 2730 | 0.003 | 0.019 | -0.035 | 0.040 | 0.000 | 0.000 | 1.000 | 0.888 | 0.947 |
| R_supramarginal_surfavg | 2723 | -0.005 | 0.025 | -0.054 | 0.044 | 0.005 | 31.945 | 1.469 | 0.850 | 0.936 |
| R_frontalpole_surfavg | 2821 | 0.012 | 0.019 | -0.025 | 0.048 | 0.000 | 0.000 | 1.000 | 0.534 | 0.916 |
| R_temporalpole_surfavg | 2748 | -0.009 | 0.019 | -0.047 | 0.028 | 0.000 | 0.000 | 1.000 | 0.625 | 0.916 |
| R_transversetemporal_surfavg | 2833 | -0.013 | 0.022 | -0.057 | 0.030 | 0.002 | 19.490 | 1.242 | 0.550 | 0.916 |
| R_insula_surfavg | 2725 | -0.010 | 0.019 | -0.047 | 0.028 | 0.000 | 0.000 | 1.000 | 0.607 | 0.916 |
| LSurfArea | 2835 | -0.012 | 0.019 | -0.048 | 0.025 | 0.000 | 0.000 | 1.000 | 0.536 | 0.916 |
| RSurfArea | 2835 | -0.010 | 0.019 | -0.047 | 0.027 | 0.000 | 0.000 | 1.000 | 0.597 | 0.916 |

### Moderator Analyses

Table S10: Schizotypy Questionaire Moderator

| Region | Q | pvalue | df |
|---|---|---|---|
| L_bankssts_thickavg | 0.1210944 | 0.7278506 | 1 |
| L_caudalanteriorcingulate_thickavg | 0.0074802 | 0.9310785 | 1 |
| L_caudalmiddlefrontal_thickavg | 0.1761127 | 0.6747354 | 1 |
| L_cuneus_thickavg | 0.5012191 | 0.4789649 | 1 |
| L_entorhinal_thickavg | 0.0052265 | 0.9423674 | 1 |
| L_fusiform_thickavg | 0.5831693 | 0.4450727 | 1 |
| L_inferiorparietal_thickavg | 0.1734402 | 0.6770723 | 1 |
| L_inferiortemporal_thickavg | 0.0547494 | 0.8149959 | 1 |
| L_isthmuscingulate_thickavg | 0.0493176 | 0.8242549 | 1 |
| L_lateraloccipital_thickavg | 0.0250159 | 0.8743275 | 1 |
| L_lateralorbitofrontal_thickavg | 4.9187359 | 0.0265669 | 1 |
| L_lingual_thickavg | 0.1367613 | 0.7115221 | 1 |
| L_medialorbitofrontal_thickavg | 0.1520148 | 0.6966173 | 1 |
| L_middletemporal_thickavg | 0.1771360 | 0.6738461 | 1 |
| L_parahippocampal_thickavg | 1.7025615 | 0.1919533 | 1 |
| L_paracentral_thickavg | 0.0618448 | 0.8036036 | 1 |
| L_parsopercularis_thickavg | 0.0890043 | 0.7654469 | 1 |
| L_parsorbitalis_thickavg | 0.0007130 | 0.9786968 | 1 |
| L_parstriangularis_thickavg | 0.0397984 | 0.8418753 | 1 |
| L_pericalcarine_thickavg | 0.2736839 | 0.6008712 | 1 |
| L_postcentral_thickavg | 0.3397632 | 0.5599660 | 1 |
| L_posteriorcingulate_thickavg | 0.4695531 | 0.4931928 | 1 |
| L_precentral_thickavg | 0.1011104 | 0.7505012 | 1 |
| L_precuneus_thickavg | 0.7395842 | 0.3897941 | 1 |
| L_rostralanteriorcingulate_thickavg | 0.5078078 | 0.4760894 | 1 |
| L_rostralmiddlefrontal_thickavg | 0.1179889 | 0.7312260 | 1 |
| L_superiorfrontal_thickavg | 0.0942487 | 0.7588437 | 1 |
| L_superiorparietal_thickavg | 0.0020676 | 0.9637321 | 1 |
| L_superiortemporal_thickavg | 1.4838901 | 0.2231669 | 1 |
| L_supramarginal_thickavg | 0.0085677 | 0.9262515 | 1 |
| L_frontalpole_thickavg | 0.1505594 | 0.6980014 | 1 |
| L_temporalpole_thickavg | 1.3129276 | 0.2518653 | 1 |
| L_transversetemporal_thickavg | 0.0342318 | 0.8532146 | 1 |

[a] Uncorrected p-values. All results were non-significant (all pFDR >.05) after FDR correction.

| Region | Q | pvalue | df |
|---|---|---|---|
| L_insula_thickavg | 0.0760752 | 0.7826883 | 1 |
| R_bankssts_thickavg | 0.0178818 | 0.8936217 | 1 |
| R_caudalanteriorcingulate_thickavg | 3.0392794 | 0.0812720 | 1 |
| R_caudalmiddlefrontal_thickavg | 1.4702307 | 0.2253093 | 1 |
| R_cuneus_thickavg | 0.7697907 | 0.3802818 | 1 |
| R_entorhinal_thickavg | 0.0016672 | 0.9674300 | 1 |
| R_fusiform_thickavg | 0.8415015 | 0.3589677 | 1 |
| R_inferiorparietal_thickavg | 0.4163542 | 0.5187618 | 1 |
| R_inferiortemporal_thickavg | 0.0284832 | 0.8659777 | 1 |
| R_isthmuscingulate_thickavg | 2.4462291 | 0.1178076 | 1 |
| R_lateraloccipital_thickavg | 0.0014794 | 0.9693182 | 1 |
| R_lateralorbitofrontal_thickavg | 0.0004730 | 0.9826490 | 1 |
| R_lingual_thickavg | 0.0499329 | 0.8231800 | 1 |
| R_medialorbitofrontal_thickavg | 0.4599144 | 0.4976640 | 1 |
| R_middletemporal_thickavg | 0.3277040 | 0.5670142 | 1 |
| R_parahippocampal_thickavg | 0.0794971 | 0.7779801 | 1 |
| R_paracentral_thickavg | 1.0389130 | 0.3080744 | 1 |
| R_parsopercularis_thickavg | 0.4811284 | 0.4879116 | 1 |
| R_parsorbitalis_thickavg | 0.7505329 | 0.3863076 | 1 |
| R_parstriangularis_thickavg | 0.3795819 | 0.5378271 | 1 |
| R_pericalcarine_thickavg | 0.0985043 | 0.7536319 | 1 |
| R_postcentral_thickavg | 1.1597696 | 0.2815133 | 1 |
| R_posteriorcingulate_thickavg | 0.1642859 | 0.6852409 | 1 |
| R_precentral_thickavg | 0.5599779 | 0.4542691 | 1 |
| R_precuneus_thickavg | 0.4096600 | 0.5221421 | 1 |
| R_rostralanteriorcingulate_thickavg | 0.3231411 | 0.5697261 | 1 |
| R_rostralmiddlefrontal_thickavg | 0.1737071 | 0.6768380 | 1 |
| R_superiorfrontal_thickavg | 0.4339876 | 0.5100387 | 1 |
| R_superiorparietal_thickavg | 1.4224680 | 0.2329976 | 1 |
| R_superiortemporal_thickavg | 0.2801484 | 0.5966039 | 1 |
| R_supramarginal_thickavg | 0.3026792 | 0.5822077 | 1 |
| R_frontalpole_thickavg | 1.8713180 | 0.1713242 | 1 |
| R_temporalpole_thickavg | 0.5870206 | 0.4435736 | 1 |

[a] Uncorrected p-values. All results were non-significant (all pFDR >.05) after FDR correction.

| Region | Q | pvalue | df |
|---|---|---|---|
| R_transversetemporal_thickavg | 0.1103299 | 0.7397689 | 1 |
| R_insula_thickavg | 0.3142672 | 0.5750734 | 1 |
| LThickness | 0.0512076 | 0.8209751 | 1 |
| RThickness | 0.3369752 | 0.5615804 | 1 |
| LFullSurf | 0.5240306 | 0.4691274 | 1 |
| RFullSurf | 1.7808076 | 0.1820502 | 1 |

[a] Uncorrected p-values. All results were non-significant (all pFDR >.05) after FDR correction.

Table S11: Scanner Number Moderator

| Region | Q | pvalue | df |
|---|---|---|---|
| L_bankssts_thickavg | 0.0017196 | 0.9669223 | 1 |
| L_caudalanteriorcingulate_thickavg | 0.2855258 | 0.5931020 | 1 |
| L_caudalmiddlefrontal_thickavg | 0.0159449 | 0.8995158 | 1 |
| L_cuneus_thickavg | 1.7940219 | 0.1804369 | 1 |
| L_entorhinal_thickavg | 0.5064899 | 0.4766623 | 1 |
| L_fusiform_thickavg | 0.0155664 | 0.9007093 | 1 |
| L_inferiorparietal_thickavg | 0.0091129 | 0.9239485 | 1 |
| L_inferiortemporal_thickavg | 1.8208068 | 0.1772173 | 1 |
| L_isthmuscingulate_thickavg | 0.7260419 | 0.3941692 | 1 |
| L_lateraloccipital_thickavg | 0.7061612 | 0.4007212 | 1 |
| L_lateralorbitofrontal_thickavg | 0.6465127 | 0.4213623 | 1 |
| L_lingual_thickavg | 0.0221572 | 0.8816696 | 1 |
| L_medialorbitofrontal_thickavg | 0.0599806 | 0.8065267 | 1 |
| L_middletemporal_thickavg | 2.5910831 | 0.1074668 | 1 |
| L_parahippocampal_thickavg | 1.7577107 | 0.1849103 | 1 |
| L_paracentral_thickavg | 0.9833386 | 0.3213759 | 1 |
| L_parsopercularis_thickavg | 0.0287153 | 0.8654380 | 1 |
| L_parsorbitalis_thickavg | 1.9717844 | 0.1602586 | 1 |
| L_parstriangularis_thickavg | 0.0790648 | 0.7785687 | 1 |
| L_pericalcarine_thickavg | 0.2553202 | 0.6133536 | 1 |
| L_postcentral_thickavg | 0.7877598 | 0.3747784 | 1 |
| L_posteriorcingulate_thickavg | 0.0759918 | 0.7828045 | 1 |
| L_precentral_thickavg | 0.2840249 | 0.5940752 | 1 |

[a] Uncorrected p-values. All results were non-significant (all pFDR >.05) after FDR correction.

| Region | Q | pvalue | df |
|---|---|---|---|
| L_precuneus_thickavg | 1.8770804 | 0.1706664 | 1 |
| L_rostralanteriorcingulate_thickavg | 2.0579533 | 0.1514134 | 1 |
| L_rostralmiddlefrontal_thickavg | 0.1207963 | 0.7281724 | 1 |
| L_superiorfrontal_thickavg | 0.0120182 | 0.9127046 | 1 |
| L_superiorparietal_thickavg | 0.0055395 | 0.9406702 | 1 |
| L_superiortemporal_thickavg | 0.0800926 | 0.7771720 | 1 |
| L_supramarginal_thickavg | 2.2609348 | 0.1326740 | 1 |
| L_frontalpole_thickavg | 0.8267064 | 0.3632266 | 1 |
| L_temporalpole_thickavg | 0.3910813 | 0.5317316 | 1 |
| L_transversetemporal_thickavg | 0.0209110 | 0.8850217 | 1 |
| L_insula_thickavg | 2.1858419 | 0.1392849 | 1 |
| R_bankssts_thickavg | 1.5433915 | 0.2141137 | 1 |
| R_caudalanteriorcingulate_thickavg | 0.2579515 | 0.6115310 | 1 |
| R_caudalmiddlefrontal_thickavg | 1.8352235 | 0.1755118 | 1 |
| R_cuneus_thickavg | 0.8475660 | 0.3572418 | 1 |
| R_entorhinal_thickavg | 0.1257431 | 0.7228872 | 1 |
| R_fusiform_thickavg | 0.5220107 | 0.4699852 | 1 |
| R_inferiorparietal_thickavg | 1.7477582 | 0.1861588 | 1 |
| R_inferiortemporal_thickavg | 0.0096758 | 0.9216418 | 1 |
| R_isthmuscingulate_thickavg | 0.3380831 | 0.5609378 | 1 |
| R_lateraloccipital_thickavg | 2.4199666 | 0.1197975 | 1 |
| R_lateralorbitofrontal_thickavg | 0.7684165 | 0.3807074 | 1 |
| R_lingual_thickavg | 1.3826518 | 0.2396500 | 1 |
| R_medialorbitofrontal_thickavg | 0.0545302 | 0.8153600 | 1 |
| R_middletemporal_thickavg | 0.3064360 | 0.5798755 | 1 |
| R_parahippocampal_thickavg | 10.6294360 | 0.0011130 | 1 |
| R_paracentral_thickavg | 0.6496865 | 0.4202248 | 1 |
| R_parsopercularis_thickavg | 2.7693669 | 0.0960842 | 1 |
| R_parsorbitalis_thickavg | 0.4203147 | 0.5167800 | 1 |
| R_parstriangularis_thickavg | 0.3428816 | 0.5581706 | 1 |
| R_pericalcarine_thickavg | 0.6571204 | 0.4175784 | 1 |
| R_postcentral_thickavg | 2.0449024 | 0.1527168 | 1 |
| R_posteriorcingulate_thickavg | 0.2225212 | 0.6371256 | 1 |

[a] Uncorrected p-values. All results were non-significant (all pFDR >.05) after FDR correction.

| Region | Q | pvalue | df |
|---|---|---|---|
| R_precentral_thickavg | 1.6571356 | 0.1979905 | 1 |
| R_precuneus_thickavg | 0.1327091 | 0.7156395 | 1 |
| R_rostralanteriorcingulate_thickavg | 0.0966012 | 0.7559470 | 1 |
| R_rostralmiddlefrontal_thickavg | 0.1302150 | 0.7182092 | 1 |
| R_superiorfrontal_thickavg | 0.4413176 | 0.5064872 | 1 |
| R_superiorparietal_thickavg | 0.1617291 | 0.6875696 | 1 |
| R_superiortemporal_thickavg | 1.6381920 | 0.2005737 | 1 |
| R_supramarginal_thickavg | 0.0491681 | 0.8245172 | 1 |
| R_frontalpole_thickavg | 0.0188587 | 0.8907723 | 1 |
| R_temporalpole_thickavg | 0.7669973 | 0.3811476 | 1 |
| R_transversetemporal_thickavg | 1.2202363 | 0.2693143 | 1 |
| R_insula_thickavg | 0.2308803 | 0.6308719 | 1 |
| LThickness | 0.4583218 | 0.4984093 | 1 |
| RThickness | 0.5620012 | 0.4534550 | 1 |
| LFullSurf | 0.8720794 | 0.3503803 | 1 |
| RFullSurf | 1.0719482 | 0.3005058 | 1 |

[a] Uncorrected p-values. All results were non-significant (all pFDR >.05) after FDR correction.

Table S12: FreeSurfer Version Moderator

| Region | Q | pvalue | df |
|---|---|---|---|
| L_bankssts_thickavg | 1.0593816 | 0.3033563 | 1 |
| L_caudalanteriorcingulate_thickavg | 1.1035228 | 0.2934943 | 1 |
| L_caudalmiddlefrontal_thickavg | 0.0914446 | 0.7623486 | 1 |
| L_cuneus_thickavg | 0.2293270 | 0.6320233 | 1 |
| L_entorhinal_thickavg | 0.8078318 | 0.3687621 | 1 |
| L_fusiform_thickavg | 0.0827799 | 0.7735650 | 1 |
| L_inferiorparietal_thickavg | 1.9919730 | 0.1581347 | 1 |
| L_inferiortemporal_thickavg | 0.3024618 | 0.5823432 | 1 |
| L_isthmuscingulate_thickavg | 0.4184781 | 0.5176974 | 1 |
| L_lateraloccipital_thickavg | 2.1544643 | 0.1421560 | 1 |
| L_lateralorbitofrontal_thickavg | 6.5508866 | 0.0104832 | 1 |
| L_lingual_thickavg | 1.8698613 | 0.1714910 | 1 |
| L_medialorbitofrontal_thickavg | 0.5262842 | 0.4681733 | 1 |

[a] Uncorrected p-values. All results were non-significant (all pFDR >.05) after FDR correction.

| Region | Q | pvalue | df |
|---|---|---|---|
| L_middletemporal_thickavg | 0.0331324 | 0.8555646 | 1 |
| L_parahippocampal_thickavg | 1.1420588 | 0.2852177 | 1 |
| L_paracentral_thickavg | 0.7724375 | 0.3794640 | 1 |
| L_parsopercularis_thickavg | 4.5958183 | 0.0320500 | 1 |
| L_parsorbitalis_thickavg | 2.5504018 | 0.1102662 | 1 |
| L_parstriangularis_thickavg | 0.1376242 | 0.7106544 | 1 |
| L_pericalcarine_thickavg | 0.6842403 | 0.4081304 | 1 |
| L_postcentral_thickavg | 0.7045372 | 0.4012633 | 1 |
| L_posteriorcingulate_thickavg | 0.1649287 | 0.6846588 | 1 |
| L_precentral_thickavg | 0.1470970 | 0.7013252 | 1 |
| L_precuneus_thickavg | 2.1313740 | 0.1443113 | 1 |
| L_rostralanteriorcingulate_thickavg | 1.6656537 | 0.1968417 | 1 |
| L_rostralmiddlefrontal_thickavg | 2.9936851 | 0.0835897 | 1 |
| L_superiorfrontal_thickavg | 2.8715194 | 0.0901597 | 1 |
| L_superiorparietal_thickavg | 0.0101249 | 0.9198501 | 1 |
| L_superiortemporal_thickavg | 1.3369722 | 0.2475686 | 1 |
| L_supramarginal_thickavg | 0.2510022 | 0.6163703 | 1 |
| L_frontalpole_thickavg | 0.0199596 | 0.8876499 | 1 |
| L_temporalpole_thickavg | 0.1277281 | 0.7207994 | 1 |
| L_transversetemporal_thickavg | 0.0032659 | 0.9544272 | 1 |
| L_insula_thickavg | 0.6720310 | 0.4123444 | 1 |
| R_bankssts_thickavg | 0.0441198 | 0.8336309 | 1 |
| R_caudalanteriorcingulate_thickavg | 1.8305626 | 0.1760611 | 1 |
| R_caudalmiddlefrontal_thickavg | 3.6185723 | 0.0571379 | 1 |
| R_cuneus_thickavg | 1.8104356 | 0.1784560 | 1 |
| R_entorhinal_thickavg | 0.3682273 | 0.5439721 | 1 |
| R_fusiform_thickavg | 0.2009810 | 0.6539301 | 1 |
| R_inferiorparietal_thickavg | 0.0250086 | 0.8743457 | 1 |
| R_inferiortemporal_thickavg | 0.2230228 | 0.6367463 | 1 |
| R_isthmuscingulate_thickavg | 0.1532732 | 0.6954268 | 1 |
| R_lateraloccipital_thickavg | 0.8605208 | 0.3535930 | 1 |
| R_lateralorbitofrontal_thickavg | 0.1433781 | 0.7049456 | 1 |
| R_lingual_thickavg | 0.4485599 | 0.5030196 | 1 |

[a] Uncorrected p-values. All results were non-significant (all pFDR >.05) after FDR correction.

| Region | Q | pvalue | df |
|---|---|---|---|
| R_medialorbitofrontal_thickavg | 0.3770957 | 0.5391617 | 1 |
| R_middletemporal_thickavg | 0.1177503 | 0.7314874 | 1 |
| R_parahippocampal_thickavg | 0.7731686 | 0.3792386 | 1 |
| R_paracentral_thickavg | 0.1018900 | 0.7495732 | 1 |
| R_parsopercularis_thickavg | 1.9817210 | 0.1592092 | 1 |
| R_parsorbitalis_thickavg | 0.9123129 | 0.3395015 | 1 |
| R_parstriangularis_thickavg | 0.4662616 | 0.4947121 | 1 |
| R_pericalcarine_thickavg | 0.0144068 | 0.9044608 | 1 |
| R_postcentral_thickavg | 0.7443519 | 0.3882704 | 1 |
| R_posteriorcingulate_thickavg | 0.2155739 | 0.6424332 | 1 |
| R_precentral_thickavg | 0.5040212 | 0.4777386 | 1 |
| R_precuneus_thickavg | 0.0003396 | 0.9852978 | 1 |
| R_rostralanteriorcingulate_thickavg | 0.6664227 | 0.4143016 | 1 |
| R_rostralmiddlefrontal_thickavg | 0.1412705 | 0.7070212 | 1 |
| R_superiorfrontal_thickavg | 1.7331530 | 0.1880087 | 1 |
| R_superiorparietal_thickavg | 0.0006000 | 0.9804571 | 1 |
| R_superiortemporal_thickavg | 0.8620471 | 0.3531665 | 1 |
| R_supramarginal_thickavg | 0.8955106 | 0.3439883 | 1 |
| R_frontalpole_thickavg | 0.2856020 | 0.5930527 | 1 |
| R_temporalpole_thickavg | 0.0817343 | 0.7749608 | 1 |
| R_transversetemporal_thickavg | 1.4248812 | 0.2326017 | 1 |
| R_insula_thickavg | 0.1640300 | 0.6854730 | 1 |
| LThickness | 0.3129084 | 0.5759009 | 1 |
| RThickness | 1.9376487 | 0.1639241 | 1 |
| LFullSurf | 4.7117448 | 0.0299572 | 1 |
| RFullSurf | 9.9401612 | 0.0016171 | 1 |

[a] Uncorrected p-values. All results were non-significant (all pFDR >.05) after FDR correction.

### Subcortical Models

##### Note - Measures of heterogeneity

$Tau^2$: Represents the absolute value of the true variance of the effect sizes, between-study variance

The $I^2$ statistic: Represents the proportion (in percentage) of total variation in the estimates that can be attributed to heterogeneity among the effects rather than sampling variability. The $I^2$ may be interpreted according to the following ranges $I^2$ = 0-40%: low heterogeneity, $I^2$ = 30-60%: moderate heterogeneity, $I^2$ = 50-90%: substantial heterogeneity, $I^2$ = 75-100%: considerable heterogeneity

The $H^2$ statistic: Represents the ratio of the total amount of variability in the effect size estimates to the amount of sampling variability

Table S13: Subcortical Correlation Model

| Region | N | Effect Size (r) | SE | Lower CI | Upper CI | $tau^2$ | $I^2$ | $H^2$ | pvalue | FDRp |
|---|---|---|---|---|---|---|---|---|---|---|
| LLatVent | 2990 | 0.002 | 0.023 | -0.043 | 0.046 | 0.003 | 24.698 | 1.328 | 0.939 | 0.985 |
| RLatVent | 2990 | 0.001 | 0.019 | -0.036 | 0.039 | 0.001 | 6.043 | 1.064 | 0.944 | 0.985 |
| Lthal | 2966 | 0.007 | 0.030 | -0.051 | 0.066 | 0.012 | 55.391 | 2.242 | 0.806 | 0.967 |
| Lcaud | 2981 | 0.017 | 0.024 | -0.029 | 0.064 | 0.004 | 29.879 | 1.426 | 0.460 | 0.923 |
| Lput | 2934 | -0.012 | 0.018 | -0.048 | 0.024 | 0.000 | 0.000 | 1.000 | 0.522 | 0.923 |
| Lpal | 2762 | -0.023 | 0.019 | -0.060 | 0.014 | 0.000 | 0.000 | 1.000 | 0.226 | 0.923 |
| Lhippo | 2970 | 0.014 | 0.018 | -0.022 | 0.050 | 0.000 | 0.000 | 1.000 | 0.443 | 0.923 |
| Lamyg | 2972 | 0.010 | 0.018 | -0.026 | 0.045 | 0.000 | 0.000 | 1.000 | 0.601 | 0.923 |
| Laccumb | 2965 | 0.012 | 0.020 | -0.027 | 0.052 | 0.001 | 10.789 | 1.121 | 0.547 | 0.923 |
| Rthal | 2986 | 0.014 | 0.031 | -0.047 | 0.075 | 0.014 | 59.948 | 2.497 | 0.654 | 0.923 |
| Rcaud | 2982 | 0.013 | 0.027 | -0.039 | 0.065 | 0.007 | 43.636 | 1.774 | 0.625 | 0.923 |
| Rput | 2965 | 0.000 | 0.018 | -0.036 | 0.036 | 0.000 | 0.000 | 1.000 | 0.985 | 0.985 |
| Rpal | 2970 | -0.032 | 0.018 | -0.067 | 0.004 | 0.000 | 0.000 | 1.000 | 0.084 | 0.923 |
| Rhippo | 2978 | 0.022 | 0.018 | -0.014 | 0.058 | 0.000 | 0.000 | 1.000 | 0.229 | 0.923 |
| Ramyg | 2960 | 0.019 | 0.024 | -0.029 | 0.067 | 0.005 | 33.861 | 1.512 | 0.444 | 0.923 |
| Raccumb | 2963 | 0.020 | 0.023 | -0.025 | 0.064 | 0.003 | 24.514 | 1.325 | 0.388 | 0.923 |

### Smoking Status - Cortical Thickness

Table S14: Effect of Schizotypy in Subgroup with Smoking Data

| Region | N | Effect Size (r) | SE | Lower CI | Upper CI | $tau^2$ | $I^2$ | $H^2$ | pvalue | FDRp |
|---|---|---|---|---|---|---|---|---|---|---|
| L_bankssts_thickavg | 1212 | -0.034 | 0.029 | -0.090 | 0.022 | 0.000 | 0.000 | 1.000 | 0.239 | 0.958 |
| L_caudalanteriorcingulate_thickavg | 1300 | 0.006 | 0.046 | -0.084 | 0.096 | 0.009 | 56.059 | 2.276 | 0.902 | 0.960 |
| L_caudalmiddlefrontal_thickavg | 1307 | -0.013 | 0.075 | -0.160 | 0.135 | 0.036 | 84.669 | 6.523 | 0.864 | 0.960 |
| L_cuneus_thickavg | 1276 | -0.010 | 0.049 | -0.107 | 0.087 | 0.011 | 61.171 | 2.575 | 0.840 | 0.960 |
| L_entorhinal_thickavg | 1233 | -0.017 | 0.033 | -0.082 | 0.049 | 0.002 | 19.491 | 1.242 | 0.614 | 0.958 |
| L_fusiform_thickavg | 1311 | -0.068 | 0.049 | -0.163 | 0.028 | 0.011 | 61.796 | 2.618 | 0.166 | 0.958 |
| L_inferiorparietal_thickavg | 1308 | -0.034 | 0.043 | -0.118 | 0.051 | 0.007 | 50.303 | 2.012 | 0.433 | 0.958 |
| L_inferiortemporal_thickavg | 1293 | -0.045 | 0.028 | -0.099 | 0.010 | 0.000 | 0.000 | 1.000 | 0.109 | 0.958 |
| L_isthmuscingulate_thickavg | 1312 | 0.004 | 0.041 | -0.075 | 0.084 | 0.005 | 44.816 | 1.812 | 0.915 | 0.964 |

[a] Sites with smoking data include BONNRISC, BONNSPQ, FOR2107-MR, FOR2017-MS, MNC, NYC, ZUR1, ZUR2

| Region | N | Effect Size (r) | SE | Lower CI | Upper CI | $tau^2$ | $I^2$ | $H^2$ | pvalue | FDRp |
|---|---|---|---|---|---|---|---|---|---|---|
| L_lateraloccipital_thickavg | 1313 | -0.011 | 0.046 | -0.102 | 0.080 | 0.009 | 57.545 | 2.355 | 0.819 | 0.960 |
| L_lateralorbitofrontal_thickavg | 1315 | -0.014 | 0.039 | -0.089 | 0.062 | 0.004 | 39.168 | 1.644 | 0.724 | 0.958 |
| L_lingual_thickavg | 1306 | -0.011 | 0.032 | -0.073 | 0.051 | 0.001 | 16.309 | 1.195 | 0.728 | 0.958 |
| L_medialorbitofrontal_thickavg | 1303 | 0.037 | 0.029 | -0.020 | 0.093 | 0.000 | 5.830 | 1.062 | 0.203 | 0.958 |
| L_middletemporal_thickavg | 1250 | -0.012 | 0.028 | -0.067 | 0.043 | 0.000 | 0.000 | 1.000 | 0.668 | 0.958 |
| L_parahippocampal_thickavg | 1313 | -0.020 | 0.041 | -0.101 | 0.060 | 0.006 | 45.539 | 1.836 | 0.616 | 0.958 |
| L_paracentral_thickavg | 1312 | -0.049 | 0.063 | -0.171 | 0.074 | 0.022 | 77.198 | 4.386 | 0.435 | 0.958 |
| L_parsopercularis_thickavg | 1304 | -0.053 | 0.049 | -0.149 | 0.043 | 0.011 | 61.722 | 2.612 | 0.280 | 0.958 |
| L_parsorbitalis_thickavg | 1309 | -0.013 | 0.028 | -0.067 | 0.042 | 0.000 | 0.000 | 1.000 | 0.649 | 0.958 |
| L_parstriangularis_thickavg | 1307 | -0.048 | 0.076 | -0.196 | 0.101 | 0.037 | 84.997 | 6.665 | 0.531 | 0.958 |
| L_pericalcarine_thickavg | 1291 | 0.006 | 0.041 | -0.073 | 0.086 | 0.005 | 44.174 | 1.791 | 0.874 | 0.960 |
| L_postcentral_thickavg | 1294 | -0.064 | 0.071 | -0.203 | 0.075 | 0.031 | 82.562 | 5.735 | 0.367 | 0.958 |
| L_posteriorcingulate_thickavg | 1311 | 0.000 | 0.028 | -0.054 | 0.054 | 0.000 | 0.000 | 1.000 | 0.997 | 0.997 |
| L_precentral_thickavg | 1298 | -0.028 | 0.044 | -0.115 | 0.058 | 0.007 | 52.802 | 2.119 | 0.521 | 0.958 |
| L_precuneus_thickavg | 1315 | -0.035 | 0.029 | -0.093 | 0.022 | 0.001 | 7.797 | 1.085 | 0.230 | 0.958 |
| L_rostralanteriorcingulate_thickavg | 1298 | 0.023 | 0.028 | -0.031 | 0.077 | 0.000 | 0.000 | 1.000 | 0.403 | 0.958 |
| L_rostralmiddlefrontal_thickavg | 1309 | 0.001 | 0.028 | -0.053 | 0.055 | 0.000 | 0.000 | 1.000 | 0.975 | 0.985 |
| L_superiorfrontal_thickavg | 1300 | -0.031 | 0.046 | -0.121 | 0.059 | 0.009 | 56.317 | 2.289 | 0.504 | 0.958 |
| L_superiorparietal_thickavg | 1306 | -0.066 | 0.053 | -0.170 | 0.038 | 0.014 | 67.730 | 3.099 | 0.211 | 0.958 |
| L_superiortemporal_thickavg | 1214 | -0.054 | 0.043 | -0.138 | 0.030 | 0.006 | 47.254 | 1.896 | 0.209 | 0.958 |
| L_supramarginal_thickavg | 1268 | -0.041 | 0.042 | -0.124 | 0.042 | 0.006 | 47.590 | 1.908 | 0.330 | 0.958 |
| L_frontalpole_thickavg | 1315 | 0.037 | 0.057 | -0.074 | 0.148 | 0.017 | 71.940 | 3.564 | 0.510 | 0.958 |
| L_temporalpole_thickavg | 1312 | 0.014 | 0.046 | -0.077 | 0.104 | 0.009 | 56.948 | 2.323 | 0.767 | 0.958 |
| L_transversetemporal_thickavg | 1315 | 0.025 | 0.065 | -0.103 | 0.153 | 0.025 | 79.204 | 4.809 | 0.701 | 0.958 |
| L_insula_thickavg | 1277 | 0.001 | 0.028 | -0.054 | 0.056 | 0.000 | 0.000 | 1.000 | 0.969 | 0.985 |
| R_bankssts_thickavg | 1275 | -0.045 | 0.028 | -0.100 | 0.009 | 0.000 | 0.000 | 1.000 | 0.105 | 0.958 |
| R_caudalanteriorcingulate_thickavg | 1310 | 0.040 | 0.062 | -0.082 | 0.162 | 0.022 | 76.811 | 4.312 | 0.520 | 0.958 |
| R_caudalmiddlefrontal_thickavg | 1312 | -0.010 | 0.028 | -0.064 | 0.044 | 0.000 | 0.000 | 1.000 | 0.713 | 0.958 |
| R_cuneus_thickavg | 1283 | -0.020 | 0.055 | -0.128 | 0.088 | 0.015 | 69.577 | 3.287 | 0.717 | 0.958 |
| R_entorhinal_thickavg | 1204 | -0.019 | 0.049 | -0.116 | 0.078 | 0.011 | 59.891 | 2.493 | 0.703 | 0.958 |
| R_fusiform_thickavg | 1314 | -0.051 | 0.033 | -0.115 | 0.013 | 0.002 | 20.514 | 1.258 | 0.119 | 0.958 |
| R_inferiorparietal_thickavg | 1304 | -0.005 | 0.062 | -0.127 | 0.116 | 0.021 | 76.427 | 4.242 | 0.929 | 0.968 |
| R_inferiortemporal_thickavg | 1307 | -0.034 | 0.041 | -0.115 | 0.047 | 0.006 | 46.304 | 1.862 | 0.415 | 0.958 |

[a] Sites with smoking data include BONNRISC, BONNSPQ, FOR2107-MR, FOR2017-MS, MNC, NYC, ZUR1, ZUR2

| Region | N | Effect Size (r) | SE | Lower CI | Upper CI | $tau^2$ | $I^2$ | $H^2$ | pvalue | FDRp |
|---|---|---|---|---|---|---|---|---|---|---|
| R_isthmuscingulate_thickavg | 1309 | -0.046 | 0.028 | -0.100 | 0.008 | 0.000 | 0.000 | 1.000 | 0.097 | 0.958 |
| R_lateraloccipital_thickavg | 1311 | -0.011 | 0.053 | -0.114 | 0.092 | 0.013 | 67.023 | 3.032 | 0.834 | 0.960 |
| R_lateralorbitofrontal_thickavg | 1312 | -0.039 | 0.055 | -0.148 | 0.069 | 0.016 | 70.208 | 3.357 | 0.476 | 0.958 |
| R_lingual_thickavg | 1303 | -0.021 | 0.050 | -0.119 | 0.077 | 0.011 | 63.143 | 2.713 | 0.677 | 0.958 |
| R_medialorbitofrontal_thickavg | 1303 | 0.061 | 0.027 | 0.007 | 0.115 | 0.000 | 0.000 | 1.000 | 0.027 | 0.958 |
| R_middletemporal_thickavg | 1303 | -0.029 | 0.028 | -0.083 | 0.025 | 0.000 | 0.000 | 1.000 | 0.293 | 0.958 |
| R_parahippocampal_thickavg | 1317 | -0.039 | 0.059 | -0.154 | 0.077 | 0.019 | 74.005 | 3.847 | 0.511 | 0.958 |
| R_paracentral_thickavg | 1309 | -0.012 | 0.063 | -0.135 | 0.112 | 0.022 | 77.599 | 4.464 | 0.854 | 0.960 |
| R_parsopercularis_thickavg | 1306 | 0.005 | 0.041 | -0.076 | 0.087 | 0.006 | 46.449 | 1.867 | 0.896 | 0.960 |
| R_parsorbitalis_thickavg | 1313 | -0.027 | 0.028 | -0.081 | 0.027 | 0.000 | 0.000 | 1.000 | 0.323 | 0.958 |
| R_parstriangularis_thickavg | 1302 | -0.014 | 0.040 | -0.094 | 0.065 | 0.005 | 43.765 | 1.778 | 0.721 | 0.958 |
| R_pericalcarine_thickavg | 1279 | 0.009 | 0.047 | -0.083 | 0.101 | 0.009 | 57.406 | 2.348 | 0.846 | 0.960 |
| R_postcentral_thickavg | 1291 | -0.062 | 0.039 | -0.138 | 0.014 | 0.004 | 40.122 | 1.670 | 0.112 | 0.958 |
| R_posteriorcingulate_thickavg | 1313 | 0.017 | 0.028 | -0.037 | 0.071 | 0.000 | 0.000 | 1.000 | 0.533 | 0.958 |
| R_precentral_thickavg | 1297 | -0.047 | 0.054 | -0.152 | 0.058 | 0.014 | 68.000 | 3.125 | 0.382 | 0.958 |
| R_precuneus_thickavg | 1312 | -0.027 | 0.055 | -0.136 | 0.081 | 0.015 | 70.344 | 3.372 | 0.621 | 0.958 |
| R_rostralanteriorcingulate_thickavg | 1289 | 0.019 | 0.041 | -0.062 | 0.100 | 0.006 | 46.023 | 1.853 | 0.641 | 0.958 |
| R_rostralmiddlefrontal_thickavg | 1309 | -0.001 | 0.028 | -0.055 | 0.053 | 0.000 | 0.000 | 1.000 | 0.965 | 0.985 |
| R_superiorfrontal_thickavg | 1306 | -0.015 | 0.044 | -0.102 | 0.072 | 0.007 | 52.905 | 2.123 | 0.734 | 0.958 |
| R_superiorparietal_thickavg | 1309 | -0.055 | 0.052 | -0.157 | 0.048 | 0.013 | 66.805 | 3.013 | 0.295 | 0.958 |
| R_superiortemporal_thickavg | 1258 | -0.047 | 0.059 | -0.163 | 0.070 | 0.019 | 73.744 | 3.809 | 0.433 | 0.958 |
| R_supramarginal_thickavg | 1278 | -0.029 | 0.028 | -0.084 | 0.025 | 0.000 | 0.000 | 1.000 | 0.293 | 0.958 |
| R_frontalpole_thickavg | 1308 | 0.018 | 0.028 | -0.036 | 0.072 | 0.000 | 0.000 | 1.000 | 0.520 | 0.958 |
| R_temporalpole_thickavg | 1306 | -0.007 | 0.045 | -0.096 | 0.082 | 0.008 | 55.114 | 2.228 | 0.874 | 0.960 |
| R_transversetemporal_thickavg | 1317 | -0.042 | 0.044 | -0.128 | 0.044 | 0.007 | 52.429 | 2.102 | 0.336 | 0.958 |
| R_insula_thickavg | 1252 | -0.023 | 0.065 | -0.152 | 0.105 | 0.025 | 78.400 | 4.630 | 0.721 | 0.958 |
| LThickness | 1318 | -0.054 | 0.060 | -0.170 | 0.063 | 0.019 | 74.949 | 3.992 | 0.368 | 0.958 |
| RThickness | 1318 | -0.053 | 0.057 | -0.165 | 0.059 | 0.017 | 72.805 | 3.677 | 0.356 | 0.958 |
| LFullSurf | 1318 | 0.017 | 0.028 | -0.037 | 0.071 | 0.000 | 0.000 | 1.000 | 0.545 | 0.958 |
| RFullSurf | 1318 | 0.013 | 0.028 | -0.041 | 0.067 | 0.000 | 0.000 | 1.000 | 0.636 | 0.958 |

[a] Sites with smoking data include BONNRISC, BONNSPQ, FOR2107-MR, FOR2017-MS, MNC, NYC, ZUR1, ZUR2

Table S15: Effect of Schizopty Controlling for Smoking Status

| Region | N | Effect Size (r) | SE | Lower CI | Upper CI | $tau^2$ | $I^2$ | $H^2$ | pvalue | FDRp |
|---|---|---|---|---|---|---|---|---|---|---|
| L_bankssts_thickavg | 1201 | -0.050 | 0.029 | -0.106 | 0.006 | 0.000 | 0.000 | 1.000 | 0.082 | 0.921 |
| L_caudalanteriorcingulate_thickavg | 1288 | 0.017 | 0.038 | -0.058 | 0.093 | 0.004 | 37.894 | 1.610 | 0.651 | 0.965 |
| L_caudalmiddlefrontal_thickavg | 1295 | -0.004 | 0.072 | -0.145 | 0.137 | 0.032 | 82.782 | 5.808 | 0.958 | 0.977 |
| L_cuneus_thickavg | 1266 | -0.010 | 0.049 | -0.106 | 0.086 | 0.010 | 60.220 | 2.514 | 0.841 | 0.965 |
| L_entorhinal_thickavg | 1221 | -0.023 | 0.035 | -0.092 | 0.045 | 0.002 | 24.152 | 1.318 | 0.504 | 0.965 |
| L_fusiform_thickavg | 1299 | -0.062 | 0.043 | -0.146 | 0.022 | 0.007 | 49.546 | 1.982 | 0.145 | 0.965 |
| L_inferiorparietal_thickavg | 1296 | -0.020 | 0.028 | -0.075 | 0.034 | 0.000 | 0.000 | 1.000 | 0.462 | 0.965 |
| L_inferiortemporal_thickavg | 1281 | -0.049 | 0.028 | -0.103 | 0.006 | 0.000 | 0.000 | 1.000 | 0.082 | 0.921 |
| L_isthmuscingulate_thickavg | 1300 | -0.020 | 0.037 | -0.092 | 0.052 | 0.003 | 32.997 | 1.492 | 0.578 | 0.965 |
| L_lateraloccipital_thickavg | 1301 | -0.021 | 0.048 | -0.116 | 0.074 | 0.010 | 60.491 | 2.531 | 0.661 | 0.965 |
| L_lateralorbitofrontal_thickavg | 1303 | -0.004 | 0.041 | -0.084 | 0.076 | 0.005 | 44.674 | 1.807 | 0.925 | 0.965 |
| L_lingual_thickavg | 1295 | -0.014 | 0.035 | -0.083 | 0.056 | 0.003 | 28.634 | 1.401 | 0.701 | 0.965 |
| L_medialorbitofrontal_thickavg | 1291 | 0.046 | 0.028 | -0.008 | 0.100 | 0.000 | 0.000 | 1.000 | 0.095 | 0.921 |
| L_middletemporal_thickavg | 1238 | -0.017 | 0.028 | -0.072 | 0.039 | 0.000 | 0.033 | 1.000 | 0.549 | 0.965 |
| L_parahippocampal_thickavg | 1302 | -0.009 | 0.028 | -0.063 | 0.046 | 0.000 | 0.000 | 1.000 | 0.754 | 0.965 |
| L_paracentral_thickavg | 1300 | -0.042 | 0.061 | -0.162 | 0.078 | 0.020 | 75.676 | 4.111 | 0.494 | 0.965 |
| L_parsopercularis_thickavg | 1292 | -0.048 | 0.034 | -0.116 | 0.019 | 0.002 | 25.734 | 1.347 | 0.162 | 0.965 |
| L_parsorbitalis_thickavg | 1297 | -0.007 | 0.028 | -0.062 | 0.047 | 0.000 | 0.000 | 1.000 | 0.792 | 0.965 |
| L_parstriangularis_thickavg | 1295 | -0.038 | 0.072 | -0.179 | 0.104 | 0.032 | 83.008 | 5.885 | 0.603 | 0.965 |
| L_pericalcarine_thickavg | 1279 | 0.005 | 0.043 | -0.080 | 0.090 | 0.007 | 49.744 | 1.990 | 0.913 | 0.965 |
| L_postcentral_thickavg | 1283 | -0.054 | 0.066 | -0.184 | 0.076 | 0.025 | 79.353 | 4.843 | 0.414 | 0.965 |
| L_posteriorcingulate_thickavg | 1299 | 0.003 | 0.028 | -0.052 | 0.057 | 0.000 | 0.000 | 1.000 | 0.924 | 0.965 |
| L_precentral_thickavg | 1286 | -0.015 | 0.036 | -0.085 | 0.055 | 0.003 | 29.552 | 1.419 | 0.673 | 0.965 |
| L_precuneus_thickavg | 1303 | -0.031 | 0.028 | -0.085 | 0.023 | 0.000 | 0.000 | 1.000 | 0.259 | 0.965 |
| L_rostralanteriorcingulate_thickavg | 1286 | 0.025 | 0.028 | -0.030 | 0.079 | 0.000 | 0.000 | 1.000 | 0.376 | 0.965 |
| L_rostralmiddlefrontal_thickavg | 1297 | 0.005 | 0.028 | -0.050 | 0.059 | 0.000 | 0.000 | 1.000 | 0.864 | 0.965 |
| L_superiorfrontal_thickavg | 1288 | -0.014 | 0.039 | -0.090 | 0.062 | 0.004 | 38.667 | 1.630 | 0.720 | 0.965 |
| L_superiorparietal_thickavg | 1295 | -0.034 | 0.036 | -0.104 | 0.036 | 0.003 | 30.189 | 1.432 | 0.344 | 0.965 |
| L_superiortemporal_thickavg | 1204 | -0.048 | 0.033 | -0.113 | 0.017 | 0.002 | 17.319 | 1.209 | 0.148 | 0.965 |
| L_supramarginal_thickavg | 1257 | -0.032 | 0.028 | -0.087 | 0.024 | 0.000 | 0.000 | 1.000 | 0.263 | 0.965 |
| L_frontalpole_thickavg | 1303 | 0.042 | 0.055 | -0.066 | 0.150 | 0.015 | 69.699 | 3.300 | 0.442 | 0.965 |
| L_temporalpole_thickavg | 1300 | 0.006 | 0.046 | -0.084 | 0.096 | 0.008 | 55.975 | 2.271 | 0.897 | 0.965 |
| L_transversetemporal_thickavg | 1303 | 0.021 | 0.066 | -0.109 | 0.150 | 0.025 | 79.424 | 4.860 | 0.756 | 0.965 |

[a] Sites with smoking data include BONNRISC, BONNSPQ, FOR2107-MR, FOR2017-MS, MNC, NYC, ZUR1, ZUR2

| Region | N | Effect Size (r) | SE | Lower CI | Upper CI | $tau^2$ | $I^2$ | $H^2$ | pvalue | FDRp |
|---|---|---|---|---|---|---|---|---|---|---|
| L_insula_thickavg | 1265 | 0.014 | 0.028 | -0.041 | 0.069 | 0.000 | 0.000 | 1.000 | 0.624 | 0.965 |
| R_bankssts_thickavg | 1263 | -0.046 | 0.028 | -0.101 | 0.009 | 0.000 | 0.000 | 1.000 | 0.099 | 0.921 |
| R_caudalanteriorcingulate_thickavg | 1298 | 0.064 | 0.052 | -0.038 | 0.165 | 0.013 | 65.301 | 2.882 | 0.219 | 0.965 |
| R_caudalmiddlefrontal_thickavg | 1300 | -0.001 | 0.028 | -0.055 | 0.053 | 0.000 | 0.000 | 1.000 | 0.970 | 0.977 |
| R_cuneus_thickavg | 1272 | -0.037 | 0.059 | -0.152 | 0.078 | 0.018 | 72.731 | 3.667 | 0.532 | 0.965 |
| R_entorhinal_thickavg | 1194 | -0.018 | 0.051 | -0.117 | 0.081 | 0.011 | 61.223 | 2.579 | 0.724 | 0.965 |
| R_fusiform_thickavg | 1302 | -0.048 | 0.028 | -0.102 | 0.006 | 0.000 | 0.000 | 1.000 | 0.080 | 0.921 |
| R_inferiorparietal_thickavg | 1292 | 0.005 | 0.052 | -0.096 | 0.106 | 0.012 | 64.730 | 2.835 | 0.926 | 0.965 |
| R_inferiortemporal_thickavg | 1295 | -0.030 | 0.043 | -0.113 | 0.054 | 0.006 | 48.554 | 1.944 | 0.482 | 0.965 |
| R_isthmuscingulate_thickavg | 1297 | -0.051 | 0.028 | -0.105 | 0.003 | 0.000 | 0.000 | 1.000 | 0.067 | 0.921 |
| R_lateraloccipital_thickavg | 1299 | -0.018 | 0.050 | -0.117 | 0.080 | 0.011 | 62.851 | 2.692 | 0.717 | 0.965 |
| R_lateralorbitofrontal_thickavg | 1300 | -0.025 | 0.052 | -0.127 | 0.077 | 0.013 | 65.511 | 2.899 | 0.634 | 0.965 |
| R_lingual_thickavg | 1292 | -0.021 | 0.052 | -0.123 | 0.081 | 0.013 | 65.559 | 2.904 | 0.683 | 0.965 |
| R_medialorbitofrontal_thickavg | 1291 | 0.076 | 0.028 | 0.022 | 0.130 | 0.000 | 0.000 | 1.000 | 0.006 | 0.408 |
| R_middletemporal_thickavg | 1291 | -0.024 | 0.028 | -0.078 | 0.030 | 0.000 | 0.000 | 1.000 | 0.389 | 0.965 |
| R_parahippocampal_thickavg | 1305 | -0.012 | 0.050 | -0.110 | 0.085 | 0.011 | 62.527 | 2.669 | 0.804 | 0.965 |
| R_paracentral_thickavg | 1297 | 0.002 | 0.054 | -0.104 | 0.109 | 0.014 | 68.688 | 3.194 | 0.969 | 0.977 |
| R_parsopercularis_thickavg | 1294 | 0.013 | 0.036 | -0.057 | 0.084 | 0.003 | 30.281 | 1.434 | 0.712 | 0.965 |
| R_parsorbitalis_thickavg | 1301 | -0.024 | 0.028 | -0.079 | 0.030 | 0.000 | 0.000 | 1.000 | 0.377 | 0.965 |
| R_parstriangularis_thickavg | 1290 | -0.022 | 0.044 | -0.108 | 0.063 | 0.007 | 50.870 | 2.035 | 0.609 | 0.965 |
| R_pericalcarine_thickavg | 1267 | 0.008 | 0.051 | -0.091 | 0.107 | 0.012 | 62.691 | 2.680 | 0.872 | 0.965 |
| R_postcentral_thickavg | 1279 | -0.057 | 0.038 | -0.131 | 0.016 | 0.004 | 35.290 | 1.545 | 0.127 | 0.965 |
| R_posteriorcingulate_thickavg | 1301 | 0.027 | 0.028 | -0.027 | 0.081 | 0.000 | 0.000 | 1.000 | 0.324 | 0.965 |
| R_precentral_thickavg | 1285 | -0.036 | 0.041 | -0.116 | 0.044 | 0.005 | 43.598 | 1.773 | 0.376 | 0.965 |
| R_precuneus_thickavg | 1300 | -0.027 | 0.052 | -0.129 | 0.074 | 0.013 | 65.343 | 2.885 | 0.599 | 0.965 |
| R_rostralanteriorcingulate_thickavg | 1277 | 0.028 | 0.042 | -0.054 | 0.110 | 0.006 | 46.562 | 1.871 | 0.505 | 0.965 |
| R_rostralmiddlefrontal_thickavg | 1297 | 0.004 | 0.028 | -0.050 | 0.058 | 0.000 | 0.000 | 1.000 | 0.878 | 0.965 |
| R_superiorfrontal_thickavg | 1294 | -0.014 | 0.049 | -0.111 | 0.083 | 0.011 | 61.596 | 2.604 | 0.773 | 0.965 |
| R_superiorparietal_thickavg | 1297 | -0.040 | 0.039 | -0.116 | 0.035 | 0.004 | 38.682 | 1.631 | 0.298 | 0.965 |
| R_superiortemporal_thickavg | 1247 | -0.024 | 0.048 | -0.118 | 0.071 | 0.010 | 58.647 | 2.418 | 0.621 | 0.965 |
| R_supramarginal_thickavg | 1266 | -0.022 | 0.028 | -0.077 | 0.033 | 0.000 | 0.000 | 1.000 | 0.440 | 0.965 |
| R_frontalpole_thickavg | 1296 | 0.017 | 0.028 | -0.037 | 0.072 | 0.000 | 0.000 | 1.000 | 0.538 | 0.965 |
| R_temporalpole_thickavg | 1294 | -0.008 | 0.045 | -0.097 | 0.081 | 0.008 | 54.394 | 2.193 | 0.864 | 0.965 |

[a] Sites with smoking data include BONNRISC, BONNSPQ, FOR2107-MR, FOR2017-MS, MNC, NYC, ZUR1, ZUR2

| Region | N | Effect Size (r) | SE | Lower CI | Upper CI | $tau^2$ | $I^2$ | $H^2$ | pvalue | FDRp |
|---|---|---|---|---|---|---|---|---|---|---|
| R_transversetemporal_thickavg | 1305 | -0.028 | 0.036 | -0.098 | 0.042 | 0.003 | 29.754 | 1.424 | 0.435 | 0.965 |
| R_insula_thickavg | 1240 | -0.011 | 0.066 | -0.140 | 0.118 | 0.025 | 78.244 | 4.596 | 0.870 | 0.965 |
| LThickness | 1306 | -0.042 | 0.051 | -0.142 | 0.057 | 0.012 | 64.337 | 2.804 | 0.404 | 0.965 |
| RThickness | 1306 | -0.040 | 0.050 | -0.137 | 0.057 | 0.011 | 62.465 | 2.664 | 0.419 | 0.965 |
| LFullSurf | 1306 | 0.018 | 0.028 | -0.036 | 0.073 | 0.000 | 0.000 | 1.000 | 0.504 | 0.965 |
| RFullSurf | 1306 | 0.013 | 0.028 | -0.041 | 0.067 | 0.000 | 0.000 | 1.000 | 0.639 | 0.965 |

[a] Sites with smoking data include BONNRISC, BONNSPQ, FOR2107-MR, FOR2017-MS, MNC, NYC, ZUR1, ZUR2

### Smoking Status - Subcortical Volume

Table S16: Effect of Schizotypy in Subgroup with Smoking Data

| Region | N | Effect Size (r) | SE | Lower CI | Upper CI | $tau^2$ | $I^2$ | $H^2$ | pvalue | FDRp |
|---|---|---|---|---|---|---|---|---|---|---|
| LLatVent | 1334 | 0.023 | 0.039 | -0.053 | 0.100 | 0.005 | 42.665 | 1.744 | 0.550 | 0.877 |
| RLatVent | 1334 | 0.006 | 0.042 | -0.076 | 0.089 | 0.006 | 49.716 | 1.989 | 0.885 | 0.996 |
| Lthal | 1325 | 0.004 | 0.066 | -0.126 | 0.134 | 0.026 | 80.206 | 5.052 | 0.954 | 0.996 |
| Lcaud | 1326 | 0.035 | 0.044 | -0.051 | 0.122 | 0.007 | 53.415 | 2.147 | 0.421 | 0.877 |
| Lput | 1292 | 0.003 | 0.031 | -0.059 | 0.064 | 0.001 | 15.079 | 1.178 | 0.934 | 0.996 |
| Lpal | 1223 | -0.035 | 0.029 | -0.092 | 0.021 | 0.000 | 0.000 | 1.000 | 0.215 | 0.877 |
| Lhippo | 1326 | 0.016 | 0.041 | -0.063 | 0.095 | 0.005 | 45.463 | 1.834 | 0.692 | 0.877 |
| Lamyg | 1320 | 0.025 | 0.027 | -0.029 | 0.079 | 0.000 | 0.000 | 1.000 | 0.364 | 0.877 |
| Laccumb | 1318 | 0.017 | 0.042 | -0.066 | 0.099 | 0.006 | 48.804 | 1.953 | 0.694 | 0.877 |
| Rthal | 1334 | 0.050 | 0.069 | -0.084 | 0.185 | 0.029 | 81.830 | 5.504 | 0.464 | 0.877 |
| Rcaud | 1330 | 0.027 | 0.052 | -0.075 | 0.129 | 0.013 | 66.927 | 3.024 | 0.607 | 0.877 |
| Rput | 1316 | -0.008 | 0.033 | -0.072 | 0.056 | 0.002 | 20.288 | 1.255 | 0.802 | 0.962 |
| Rpal | 1315 | -0.035 | 0.043 | -0.119 | 0.048 | 0.007 | 50.506 | 2.020 | 0.408 | 0.877 |
| Rhippo | 1329 | 0.023 | 0.030 | -0.036 | 0.082 | 0.001 | 12.474 | 1.143 | 0.445 | 0.877 |
| Ramyg | 1315 | 0.031 | 0.027 | -0.023 | 0.085 | 0.000 | 0.000 | 1.000 | 0.256 | 0.877 |
| Raccumb | 1319 | 0.024 | 0.052 | -0.078 | 0.126 | 0.013 | 66.713 | 3.004 | 0.649 | 0.877 |

[a] Sites with smoking data include BONNRISC, BONNSPQ, FOR2107-MR, FOR2017-MS, MNC, NYC, ZUR1, ZUR2

Table S17: Effect of Schizopty Controlling for Smoking Status

| Region | N | Effect Size (r) | SE | Lower CI | Upper CI | $tau^2$ | $I^2$ | $H^2$ | pvalue | FDRp |
|---|---|---|---|---|---|---|---|---|---|---|
| LLatVent | 1137 | 0.004 | 0.037 | -0.069 | 0.077 | 0.002 | 25.333 | 1.339 | 0.914 | 0.962 |
| RLatVent | 1137 | 0.014 | 0.056 | -0.095 | 0.123 | 0.012 | 64.227 | 2.795 | 0.799 | 0.962 |
| Lthal | 1137 | 0.004 | 0.082 | -0.157 | 0.165 | 0.037 | 84.505 | 6.454 | 0.962 | 0.962 |
| Lcaud | 1129 | 0.028 | 0.048 | -0.065 | 0.122 | 0.007 | 50.868 | 2.035 | 0.554 | 0.962 |
| Lput | 1098 | -0.019 | 0.030 | -0.078 | 0.040 | 0.000 | 0.000 | 1.000 | 0.530 | 0.962 |
| Lpal | 1034 | -0.030 | 0.031 | -0.091 | 0.031 | 0.000 | 0.000 | 1.000 | 0.333 | 0.962 |
| Lhippo | 1132 | -0.008 | 0.030 | -0.066 | 0.051 | 0.000 | 0.000 | 1.000 | 0.794 | 0.962 |
| Lamyg | 1127 | -0.003 | 0.030 | -0.061 | 0.056 | 0.000 | 0.000 | 1.000 | 0.926 | 0.962 |
| Laccumb | 1123 | 0.009 | 0.050 | -0.090 | 0.107 | 0.009 | 55.603 | 2.252 | 0.859 | 0.962 |
| Rthal | 1137 | 0.059 | 0.088 | -0.114 | 0.233 | 0.045 | 86.955 | 7.666 | 0.501 | 0.962 |
| Rcaud | 1133 | 0.015 | 0.057 | -0.097 | 0.127 | 0.013 | 65.786 | 2.923 | 0.797 | 0.962 |
| Rput | 1120 | -0.036 | 0.030 | -0.095 | 0.023 | 0.000 | 0.000 | 1.000 | 0.228 | 0.962 |
| Rpal | 1119 | -0.036 | 0.037 | -0.107 | 0.036 | 0.002 | 22.917 | 1.297 | 0.329 | 0.962 |
| Rhippo | 1136 | -0.031 | 0.030 | -0.089 | 0.027 | 0.000 | 0.000 | 1.000 | 0.295 | 0.962 |
| Ramyg | 1122 | 0.014 | 0.030 | -0.044 | 0.072 | 0.000 | 0.000 | 1.000 | 0.641 | 0.962 |
| Raccumb | 1124 | 0.014 | 0.050 | -0.084 | 0.112 | 0.009 | 55.242 | 2.234 | 0.781 | 0.962 |

[a] Sites with smoking data include BONNRISC, BONNSPQ, FOR2107-MR, FOR2017-MS, MNC, NYC, ZUR1, ZUR2

### Measures of Schizotypy

Table S18: Schizotypy Scales

| Questionnaires | Total number of items | Factors/Subscales |
|---|---|---|
| SPQ | 74 | Cognitive/Perceptual; Interpersonal; Disorganized |
| SPQ-B | 22 | Cognitive/Perceptual; Interpersonal; Disorganized |
| CAPE | 42 | Depression, Negative, Positive Symptoms |
| OLIFE | 104 | Unusual Experiences; Cognitive Disorganization; Introvertive Anhedonia; Impulsive Nonconformity |
| RISC | 26 | Positive and Cognitive content of schizotypy |
| CHAP | 196 | Perceptual Aberration; Magical Ideation; Physical Anhedonia; Social Anhedonia; Impulsive Nonconformity |

[a] Note. CHAP=Chapman scales; CAPE=Community Assessment of Psychotic Experiences;
Schizotypal Personality Questionnaire=SPQ; OLIFE=Oxford-Liverpool Inventory of Feelings and Experiences;
RISC=Rust Inventory of Schizotypal Cognitions.

### Funding and Acknowledgments

MK acknowledges support from the National Bank Fellowship (McGill University) and the Swiss National Foundation (P2SKP3_178175). GM was supported by a Sir Henry Dale Fellowship jointly funded by the Wellcome Trust and the Royal Society (grant number 202397/Z/16/Z). MG was supported by NHMRC as an R.D. Wright Biomedical Career Development Fellow (#1061875). CP was supported by NHMRC Senior Principal Research Fellowships (#628386 & #1105825) and a NHMRC Program Grant (ID: 1150083). AF was supported by the Sylvia and Charles Viertel Charitable Foundation; and the National Health and Medical Research Council (ID: 1050504). MAB is supported by a Senior Research Fellowship from the Australian National Health and Medical Research Council (NHMRC). AK was supported by the German Research Foundation (KR 3822/5-1, KR 3822/7-2). YW was supported by the National Natural Science Foundation of China (31871114), CAS Key Laboratory of Mental Health, Institute of Psychology. ET was supported by the German Research Foundation (DFG 31/2-1). TK was funded by the German Research Foundation (DFG FOR2107 KI588/14-1 and KI588/14-2). IL was funded by RBRF (20-013-00748). SL acknowledges funding from Fonds de la Recherche du Québec – Santé (FRQ-S) and the Canadian Institutes of Health Research (CIHR). PK was funded by a University of Roehampton Vice Chancellor Scholarship. SK was funded by the Swiss National Science Foundation. IN was funded by the DFG: FOR2107, FSU Jena: Junior Scientist Programme, UKGM: FoFoe. MD was funded by the Schweizerischer Nationalfonds zur Förderung der Wissenschaftlichen Forschung (Grant/Award Number: 100019_159440). PD was supported by the National Institute of Mental Health (NIMH R00 MH086756; NIMH P50 MH080173). WR was funded by Collegium Helveticum, Transdisciplinary Research Institute of ETH Zurich and University Zurich. MJG was supported by National Health and Medical Research Council (NHMRC) Project Grants 630471 and 1081603. AD was supported by the Canadian Institutes of Health Research. JT was supported by the NIMH (5R01MH116147 (PI P. Thompson). BB was funded by the Canada Research Chairs Program.

FOR2107: This work was funded by the German Research Foundation (DFG, grant FOR2107 DA1151/5-1 and DA1151/5-2 to UD; SFB-TRR58, Projects C09 and Z02 to UD) and the Interdisciplinary Center for Clinical Research (IZKF) of the medical faculty of Münster (grant Dan3/012/17 to UD).

DECOP: This study was funded by the Netherland Organization for Scientific Research (NWO) [Veni #451-13-035].

ASRB: The Australian Schizophrenia Research Bank (ASRB) was supported by the National Health and Medical Research Council of Australia (NHMRC) (Enabling Grant, ID 386500), the Pratt Foundation, Ramsay Health Care, the Viertel Charitable Foundation and the Schizophrenia Research Institute. Chief Investigators for ASRB were Carr, V., Schall, U., Scott, R., Jablensky, A., Mowry, B., Michie, P., Catts, S., Henskens, F., Pantelis, C. We thank Loughland, C., the ASRB Manager, and acknowledge the help of Jason Bridge for ASRB database queries.

IGP: This study was funded by Project Grants from the Australian National Health and Medical Research Council of Australia (NHMRC; APP630471 and APP1081603), the Macquarie University's Australian Research Council Centre of Excellence in Cognition and its Disorders (CE110001021).

London1a: This work was supported by a Brain & Behavior Research Foundation NARSAD Young Investigator Grant to GM (#21200, Lieber Investigator).

London1b: Funding for this study was provided by European Science Foundation EURYI grant (NWO 044035001 to AA).

AUCK: This study was funded by the Faculty Research Development Fund (No. 3702215) and the MRI Pilot Study Fund (Skyra 25- 001-A) from the University of Auckland.

CAM: This work was supported by the Wellcome Trust (to PM and WS) and the Niels Stensen Foundation (to KMJD).
